# No one-size-fits-all approach: Retrospective analysis of efficacy and safety of serum concentrations of continuously administered vancomycin in critically ill adults reveals different target serum concentrations depending on disease severity

**DOI:** 10.1101/2024.12.26.24319482

**Authors:** Katrin Viertel, Carmen van Meegen, Swetlana Herbrandt, Thorsten Annecke, Frauke Mattner

**Affiliations:** Central Pharmacy, Cologne Merheim Medical Centre, University Hospital of Witten/Herdecke, Ostmerheimer Str. 200, 51109 Cologne, Germany; Institute of Hygiene, Cologne Merheim Medical Centre, University Hospital of Witten/Herdecke, Ostmerheimer Str. 200, 51109 Cologne, Germany; Division of Hygiene and Environmental Medicine, Department of Human Medicine, Faculty of Health, Witten/Herdecke University, Witten, Alfred-Herrhausen-Straße 50, 58455 Witten, Germany; Center for Higher Education, Statistical Consulting and Analysis, TU Dortmund University, Vogelpothsweg 78, 44221, Dortmund, Germany; Department of Anaesthesiology and Intensive Care Medicine, Cologne Merheim Medical Centre, University Hospital of Witten/Herdecke, Ostmerheimer Str. 200, 51109 Cologne, Germany

**Keywords:** vancomycin, drug level, continuous infusion, intensive care unit (ICU), infection, acute kidney injury (AKI), mortality

## Abstract

**Background:** Vancomycin is frequently monitored, but target levels for continuous infusion of vancomycin (CIV) are based on expert opinion. Rarely have vancomycin concentrations been correlated with therapeutic efficacy or safety of CIV.

**Objectives:** Associations between vancomycin steady-state serum concentrations and treatment failure or toxicity with CIV were examined.

**Methods:** A retrospective, single-centre cohort study was conducted of consecutive critically ill surgical patients receiving CIV between 2010-2022. After detecting associations between vancomycin levels, renal function and health status, four subgroups were defined based on estimated glomerular filtration rate (</≥90mL/min/1.73m²) and Simplified Acute Physiology Score (SAPS) II (≤/>36). Failure and toxicity of vancomycin serum concentrations were assessed using primary (mortality, acute kidney injury (AKI)) and secondary (clinical and microbiological failure) endpoints. Predictors of outcome parameters were identified using logistic and Cox regression. Concentrations were compared by bivariate comparisons, post-hoc tests following analysis of variance for the regression models and desirability of outcome ranking. Concentration cut-offs were determined by receiver-operating characteristic and classification and regression tree analyses.

**Results:** 922 patients were included. Higher vancomycin concentrations (first 72h average; specifically >25mg/L) were associated with higher mortality, AKI and clinical failure, but less microbiological failure. For SAPS>36, concentrations <20mg/L (i.e. 15-20mg/L or <17mg/L) correlated with the best treatment outcome, for SAPS≤36 concentrations >19mg/L (i.e. 20-25mg/L or 19-28mg/L).

**Conclusion:** Retrospective analyses of vancomycin serum concentrations during CIV suggest that ICU patients’ disease severity should be considered when selecting a target concentration. The target concentration might be sought inversely related to SAPS, which should be confirmed in future prospective controlled trials.

## 1 Introduction

Infections occur frequently in intensive care units (ICUs) with more than 50% of patients affected [1]. Up to 22% of patients contracted the infection within the ICU [1]. One third (22-74%) of infections were caused by Gram-positive pathogens [2]. Vancomycin is an antibiotic that is regularly used empirically for sepsis and for the treatment of infections with Gram-positive bacteria (e.g. *methicillin-resistant Staphylococcus aureus* (MRSA), coaculase-negative staphylococci (CoNS), *Enterococcus faecium*) in ICUs [3]. The risk of under- or overdosing vancomycin is considerable due to the narrow therapeutic range and inter- and intra-individual pharmacokinetic variability [4], which is particularly high in critically ill patients [5]. Underdosing may lead to treatment failure (i.e. infection-related mortality, persistence, progression or recurrence of signs and symptoms of infection or bacteria due to resistance to vancomycin) [6]. In particular, overdosage may cause acute kidney injury (AKI) [7], which may worsen the patient’s outcome in intensive care [8]. To improve efficacy (i.e. reduce mortality, eliminate or improve signs and symptoms of infection, eradicate bacteria) and reduce toxicity, vancomycin serum levels are controlled by therapeutic drug monitoring (TDM) [4]. Continuous infusion of vancomycin (CIV) had some advantages over intermittent infusion of vancomycin (IIV) in this context [9]. However, unlike IIV, no target vancomycin serum levels have yet been validated for CIV [4]. While the revised consensus guideline on vancomycin for MRSA infections suggested a target concentration of 20-25 mg/L for CIV [4], other authors aimed for different target values [10]. Higher mortality and lower clinical cure have been reported at concentrations <15 mg/L [11, 12]. With increasing concentrations, an increasing rate of AKI was observed [12–14], for which we calculated a cut-off value of 25 mg/L [10]. Average concentrations and corresponding frequencies of treatment success, failure and nephrotoxicity in specific cohorts have been described [11, 12, 14, 15]. Comparative analyses that determined the most appropriate vancomycin steady-state serum concentration (ranges) in terms of treatment outcomes are still rare. Hence, a retrospective analysis (i.e. bivariate comparisons, regression models, desirability of outcome ranking (DOOR), cut-offs by receiver-operating characteristic (ROC) and classification and regression tree (CART) methods) of a routine vancomycin TDM programme in surgical ICU patients was conducted to identify vancomycin serum concentrations and patient parameters associated with the lowest rates of treatment failure (e.g. mortality, clinical and microbiological failure) and toxicity (e.g. AKI) during CIV. Due to the correlation we found between vancomycin serum concentration, renal function and physiological score, this study was designed as a four-arm subgroup analysis.

## 2 Materials and Methods

### 2.1 Study design

In a retrospective, single-centre cohort study, all consecutive inpatients admitted to the interdisciplinary surgical ICU at the Cologne-Merheim Hospital in Germany between 1 May 2010 and 5 August 2022 were included.

#### 2.1.1 Subgroups

We originally intended to analyse vancomycin serum concentrations only for the entire cohort. However, we realised that estimated glomerular filtration rate (eGFR; calculated using the Chronic Kidney Disease Epidemiology Collaboration (CKD-EPI) 2021 formula [16]) at the start of CIV and Simplified Acute Physiology Score (SAPS) II [17] at the start of CIV showed opposite concentration-dependent behaviour (supplementary material **Figure S 1**) and that eGFR, SAPS and mean vancomycin serum concentration during the first three days of CIV (C72mean) were correlated (supplementary material **Table S 1**). As a result, it was unlikely that an optimal target range for all patients could have been identified across the cohort. To reduce the mutual influence of the correlated parameters, we structured our evaluation (from the outset) as a subgroup analysis with eGFR and SAPS classes. eGFR was divided into good (eGFR ≥90 mL/min/1.73 m^2^: 60%) and poor (eGFR <90 mL/min/1.73 m^2^: 40%) (supplementary material **Table S 3a**) using KDIGO (Kidney Disease: Improving Global Outcomes) definitions [18]. There were too few patients with severely impaired renal function (17% <60 mL/min/1.73 m^2^, 3% <30 mL/min/1.73 m^2^) to select a lower threshold. A median split was used for SAPS (median=36) (supplementary material **Table S 3b**), supported by similar cut-offs calculated with ROC or CART (∼40). Four eGFR/SAPS subgroups were defined: subgroup A with eGFR <90 mL/min/1.73 m^2^ and SAPS >36, subgroup B with eGFR ≥90 mL/min/1.73 m^2^ and SAPS >36, subgroup C with eGFR <90 mL/min/1.73 m^2^ and SAPS ≤36, subgroup D with eGFR ≥90 mL/min/1.73 m^2^ and SAPS ≤36.

### 2.2 Ethics

The study was conducted in accordance with the Declaration of Helsinki (2013 version) and was approved by the Ethics Committee of the University of Witten-Herdecke, Germany, with a waiver of the need for patients’ informed consent (Chair: Prof. Dr. med. P.W. Gaidzik, No. S-55/2022).

### 2.3 Patient eligibility

Eligible patients were identified using our electronic patient data management and documentation systems (detailed description in the supplementary material). Patients were included if they: (a) were 18 years of age and older, (b) were treated with CIV for 24 hours or longer, (c) did not receive a change in treatment modality from IIV to CIV or vice versa, (d) underwent daily drug monitoring during the first days of therapy, (e) had a total body weight (TBW), serum creatinine (SCr), urine output and SAPS II recorded in the data file during treatment, (f) were admitted to the ICU in connection with surgery. Patients with less than 48 hours of CIV, renal replacement therapy (RRT) at the start of CIV and pregnancy were excluded (**Figure 1**).

**Figure 1.**
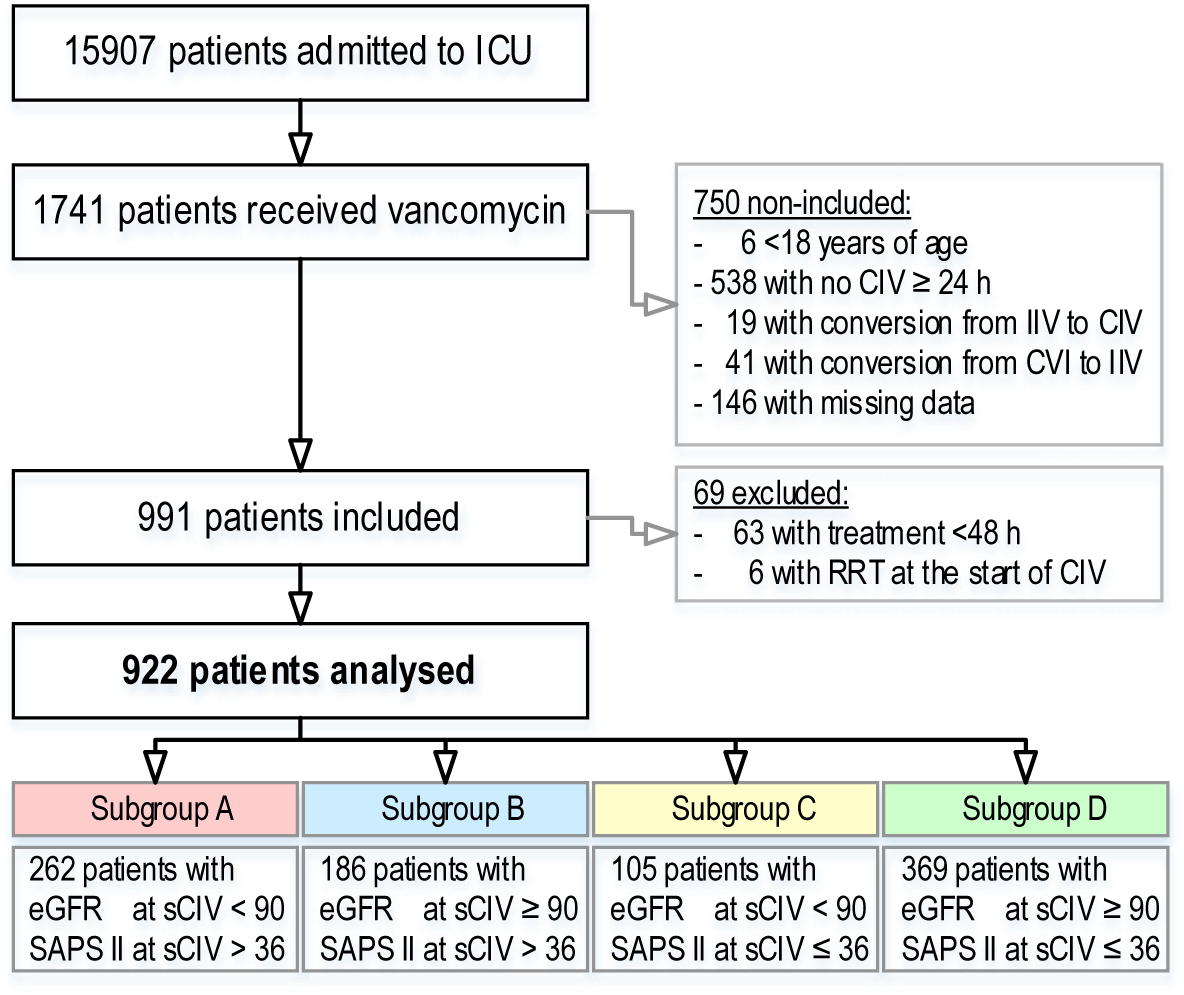
Flowchart illustrating the inclusion and exclusion of patients. Abbreviations: CIV, continuous infusion of vancomycin; eGFR, estimated glomerular filtration rate [mL/min/1.73 m^2^]; ICU, intensive care unit; IIV: intermittent infusion of vancomycin; RRT, renal replacement therapy; SAPS, Simplified Acute Physiology Score; sCIV, start of continuous infusion of vancomycin.

### 2.4 Objectives and assessment criteria

The failure and toxicity of steady-state serum concentrations during CIV were evaluated using primary and secondary endpoints.

#### 2.4.1 Primary outcomes

##### 2.4.1.1 Failure

The primary failure endpoint was all-cause mortality in the ICU. In addition, all-cause in-hospital mortality and in-hospital mortality at day 30 after CIV initiation were evaluated.

##### 2.4.1.2 Toxicity

The primary toxicity endpoint was the incidence of nephrotoxicity, defined as new-onset of acute kidney injury (AKI) according to Acute Kidney Injury Network (AKIN) or KDIGO [19, 20] criteria (supplementary material **Table S 5**), between the start of treatment until three days after the end of CIV, distinguishing between early AKI (within the first 48 hours after the start of CIV) and late AKI (48 hours after the start to 72 hours after the end of CIV) [13]. The diagnostic criteria for AKI were an increase in SCr of at least 50% within seven days, an increase in SCr of ≥0.3 mg/dL within 48 hours, urine output <0.5 mL/kg TBW/h for 6-12 hours, or initiation of dialysis.

#### 2.4.2 Secondary outcomes

##### 2.4.2.1 Failure

Secondary failure endpoints were evaluated in terms of clinical and microbiological treatment failure (detailed description in the supplementary material).

##### 2.4.2.2 Toxicity

Secondary toxicity endpoints included time to onset of AKI, frequency of different AKI stages, reversibility of AKI at hospital discharge and change in renal function between the start of CIV and day 28.

### 2.5 Data collection

Data were collected on demographics, medical history, ICU stay, medication, infection, infection laboratory, microbiology, renal function and vancomycin therapy (**Table 1**, **Table 2** and supplementary material **Table S 4**).

**Table 1.**
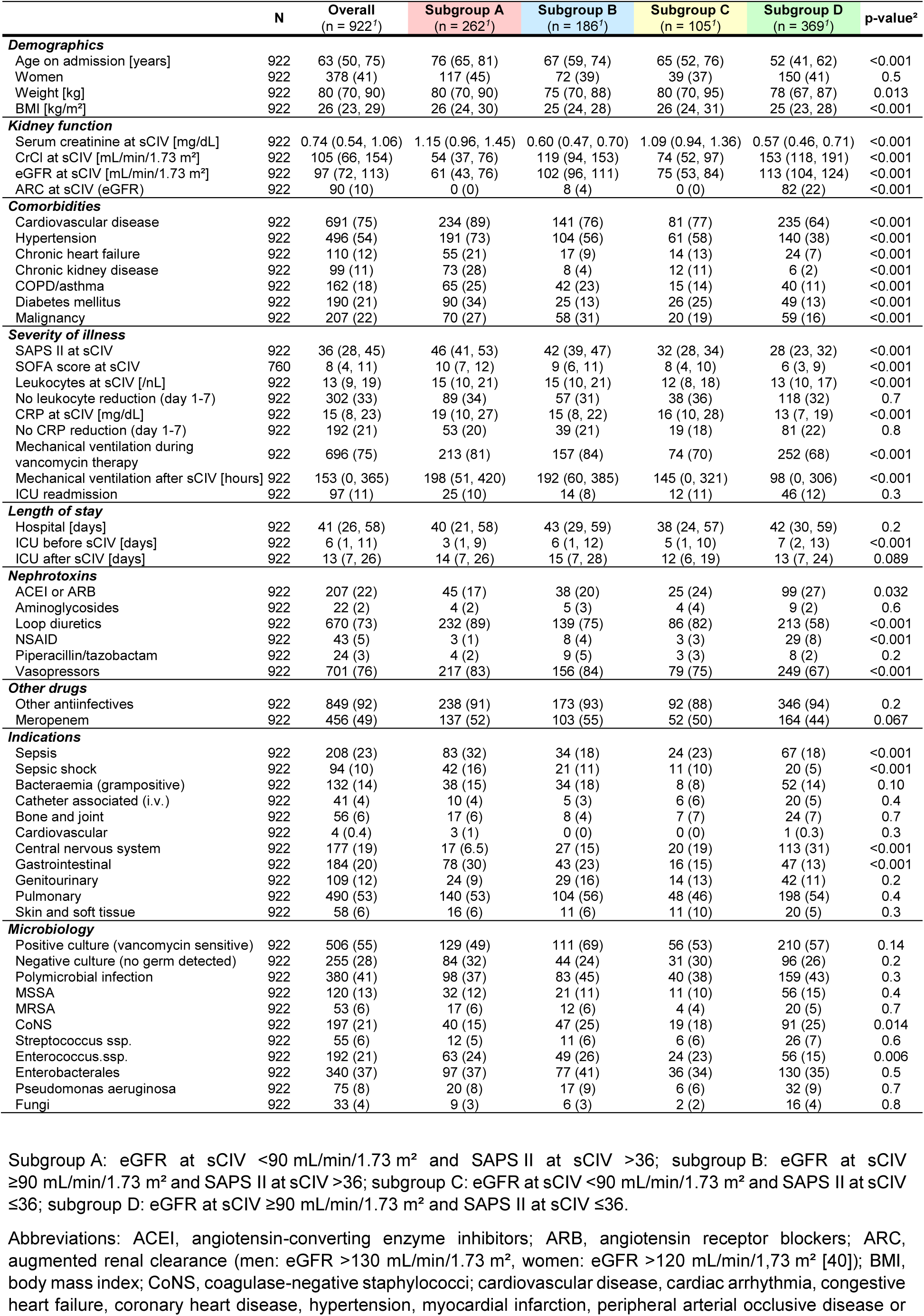
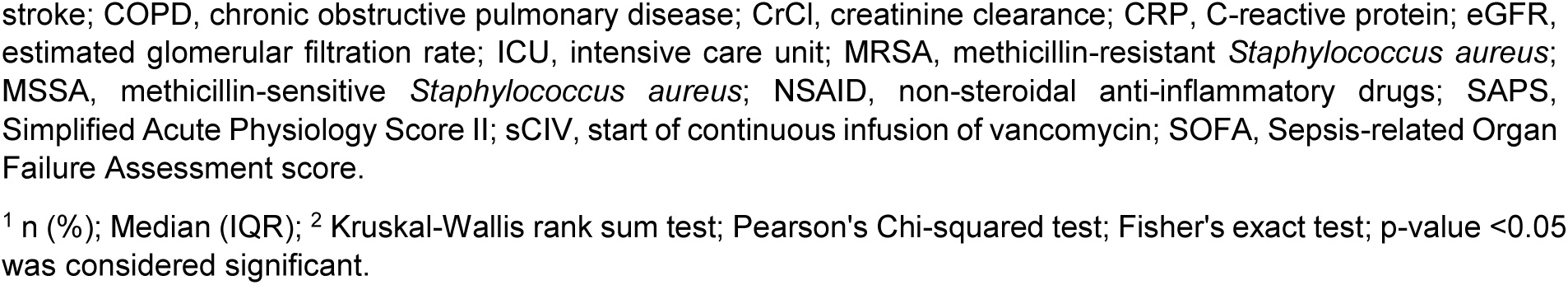
Characteristics of the study population, with regard to the kidney function and SAPS II at the start of continuous infusion of vancomycin (n=922).

**Table 2.** Vancomycin therapy.

|  | N | Overall<br>(n = 922<sup>1</sup>) | Subgroup A<br>(n = 262<sup>1</sup>) | Subgroup B<br>(n = 186<sup>1</sup>) | Subgroup C<br>(n = 105<sup>1</sup>) | Subgroup D<br>(n = 369<sup>1</sup>) | p-value<sup>2</sup> |
| --- | --- | --- | --- | --- | --- | --- | --- |
| Empirical therapy | 922 | 609 (66) | 187 (71) | 121 (65) | 71 (68) | 230 (62) | 0.12 |
| Loading dose atp | 922 | 53 (7) | 14 (5) | 7 (4) | 5 (5) | 27 (7) | 0.3 |
| Initial maintenance dose atp | 922 | 298 (32) | 77 (29) | 62 (33) | 30 (29) | 129 (35) | 0.4 |
| Loading dose [mg/kg TBW] | 922 | 12.5 (10.7, 15.4) | 12.5 (10.0, 14.6) | 13.3 (11.1, 15.4) | 12.5 (10.0, 15.4) | 12.5 (11.1, 15.9) | 0.2 |
| Dose day 1 [mg/kg TBW] | 922 | 38.6 (32.8, 46.4) | 37.7 (30.0, 43.1) | 40.2 (33.5, 47.3) | 37.7 (31.1, 43.7) | 40.2 (33.5, 49.4) | <0.001 |
| Dose day 1-3 [mg/kg TBW/day] | 922 | 31.3 (26.1, 37.2) | 27.9 (23.3, 32.2) | 32.9 (28.4, 40.2) | 27.8 (23.8, 33.8) | 33.9 (28.3, 39.8) | <0.001 |
| Duration of therapy [days] | 922 | 5.4 (3.7, 7.3) | 5.3 (3.2, 7.1) | 5.2 (3.8, 7.1) | 4.8 (3.0, 6.9) | 5.8 (4.0, 7.6) | 0.016 |
| CL<sub>vancomycin</sub> day 1-3 [L/h] | 922 | 4.9 (3.5, 6.6) | 3.4 (2.6, 4.4) | 5.4 (4.2, 6.5) | 4.3 (3.1, 5.8) | 6.2 (4.7, 7.9) | <0.001 |
| C24 [mg/L] | 922 | 18.1 (14.2, 23.3) | 23.0 (18.1, 28.1) | 17.1 (14.3, 21.1) | 21.0 (16.3, 27.8) | 15.4 (12.3, 19.4) | <0.001 |
| C72mean [mg/L] | 922 | 20.8 (16.6, 26.7) | 27.3 (21.9, 31.9) | 19.7 (16.7, 24.3) | 23.9 (18.4, 32.1) | 17.8 (14.3, 21.2) | <0.001 |
| Cmean [mg/L] | 922 | 22.0 (17.8, 27.4) | 27.5 (23.3, 32.4) | 21.5 (18.3, 25.2) | 24.3 (19.6, 31.3) | 19.0 (15.8, 22.2) | <0.001 |

Subgroup A: eGFR at sCIV <90 mL/min/1.73 m<sup>2</sup> and SAPS II at sCIV >36; subgroup B: eGFR at sCIV ≥90 mL/min/1.73 m<sup>2</sup> and SAPS II at sCIV >36; subgroup C: eGFR at sCIV <90 mL/min/1.73 m<sup>2</sup> and SAPS II at sCIV ≤36; subgroup D: eGFR at sCIV ≥90 mL/min/1.73 m<sup>2</sup> and SAPS II at sCIV ≤36.
Abbreviations: atp, according to protocol; C24, vancomycin serum concentration after day 1 of continuous infusion of vancomycin; C72mean, mean vancomycin serum concentration during the first three days of continuous infusion of vancomycin; CL, clearance; Cmean, mean vancomycin serum concentration over the entire period of continuous infusion of vancomycin; eGFR, estimated glomerular filtration rate; SAPS, Simplified Acute Physiology Score II; sCIV, start of continuous infusion of vancomycin; TBW, total body weight.
<sup>1</sup> n (%); Median (IQR); <sup>2</sup> Kruskal-Wallis rank sum test; Pearson's Chi-squared test; p-value <0.05 was considered significant.

#### 2.5.1 Vancomycin therapy/administration

Vancomycin was reconstituted according to the instructions of the prescribing information and applied in a perfusor at a concentration of 40 mg/mL (2 g/50 mL). The standard protocol was to administer vancomycin on a weight basis. TBW was used; if TBW was not available in the medical record, it was estimated by the attending physician. The initial dose was 30 mg/kg TBW, followed by a standard daily dose of 30 mg/kg TBW (supplementary material **Table S 6**). In addition, the dose was adjusted on a sliding scale according to renal function (supplementary material **Table S 7**). Renal function was estimated using the CKD-EPI formula [21].

#### 2.5.2 Vancomycin therapeutic drug monitoring

Blood samples were taken daily along with routine laboratory tests. Vancomycin serum concentrations were measured using an immunoassay technique (CMIA). Treatment was adjusted to achieve a plateau concentration of 20-30 mg/L. If the concentration was insufficient (<20 mg/L), a bolus of 500 mg was given and the daily dose was increased by 480 mg (+0.5 mL/h). If the concentration was excessive, CIV was a) interrupted for 4 hours and the daily dose was decreased by 480 mg (-0.5 mL/h) if the vancomycin concentration was 30-40 mg/L, or b) paused for 12 hours and the daily dose was decreased by 960 mg (-1.0 mL/h) if the vancomycin concentration was >40 mg/L (supplementary material **Table S 8**).

### 2.6 Statistical analysis

Collected data were summarised using Microsoft Excel (version 16.0.5317.1000; Microsoft Corporation, Redmond, WA USA)). Statistical analyses were performed using the statistical software R (version 4.3.2, R Foundation for Statistical Computing, Vienna, Austria) and RStudio (version 2023.06.0+421 “Mountain Hydrangea”, Posit Software, PCB). Descriptive statistics were calculated for each study variable. Histograms and normal-quantile plots were used to check whether the univariate distribution of continuous variables was normal. Categorical variables were expressed as frequencies and proportions, and quantitative variables as mean±standard deviation (SD) if normally distributed, or median and interquartile range (IQR) if not. Characteristics between the subgroups were compared using Fisher’s exact test, Pearson’s Chi-squared test or Kruskal-Wallis rank sum test, as appropriate (**Table 1**, **Table 2**, **Table 3**). P-values <0.05 were considered statistically significant. Three classes were defined for C72mean based on the currently recommended concentration range [4]: low (<20 mg/L), optimal (20-25 mg/L) and high (>25 mg/L) (supplementary material **Table S 3c**). Bivariate comparisons between these classes and outcome parameters were performed to investigate their relationship. Logistic regression was used to model the odds of mortality, developing renal insufficiency and clinical or microbiological treatment failure as a function of various influencing variables (supplementary material **Table S 9**). Variables for which an effect was known or suspected were used as predictors. In addition, the 2-way interactions between eGFR, SAPS and C72mean were taken into account. Collinearity between variables was checked prior to modelling. Cox regression was used to model the (censored) survival time and the time to onset of acute renal failure as a function of several predictor variables (supplementary material **Table S 9**). The same predictors were used as in the logistic regression. Following an analysis of variance (ANOVA) for each of the models described above, post-hoc tests with Bonferroni correction were performed to compare the three C72mean classes. DOOR analysis was performed to show the associations between five C72mean classes (<15, 15-<20, 20-<25, 25-30, >30 mg/L) and overall patient outcome (supplementary material **Table S 9**). Each patient was assigned an overall outcome based on ICU mortality, AKI and clinical and microbiological treatment success or failure. The 9 outcome levels, from most to least desirable, were: (1) no AKI, microbiological and clinical success, (2) no AKI, microbiological success and clinical failure with survival, (3) no AKI, microbiological failure and clinical success, (4) no AKI, microbiological failure and clinical failure with survival, (5) AKI, microbiological and clinical success, (6) AKI, microbiological success and clinical failure with survival, (7) AKI, microbiological failure and clinical success, (8) AKI, microbiological failure and clinical failure with survival, (9) ICU mortality. ROC and CART analyses were used to identify alternative optimal C72mean ranges associated with the outcome parameters (supplementary material **Table S 9**). The Youden index was used to determine an ‘optimal’ cut-off in the ROC analysis. In the CART models, the complexity parameter was set to -0.01 and the maximal depth to 2. Odds ratios, 95% confidence intervals (CIs) and p-values were calculated to assess the cut-offs. **Figure 2** shows an illustration of the analysis plan.

**Figure 2.**
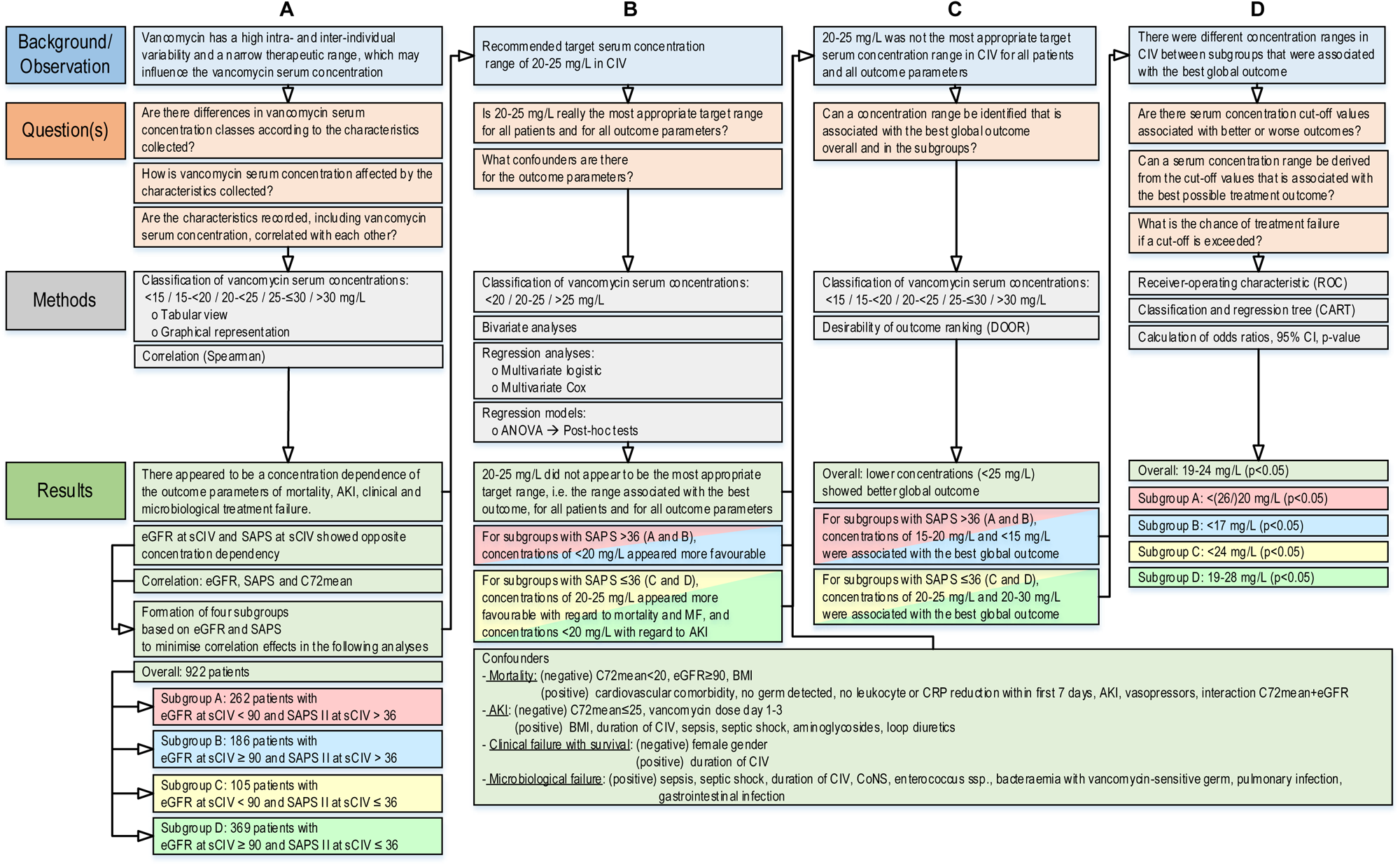
Flowchart illustrating the conduct of the study, including observations, questions, methods and summary of results. Overall (n=922); subgroup A (n=262): eGFR at sCIV <90 mL/min/1.73 m^2^ and SAPS II at sCIV >36; subgroup B (n=186): eGFR at sCIV ≥90 mL/m 563 in/1.73 m^2^ and SAPS II at sCIV >36; subgroup C (n=105): eGFR at sCIV <90 mL/min/1.73 m^2^ and SAPS II at sCIV ≤36; subgroup D (n=369): eGFR at sCIV ≥90 mL/min/1.73 m^2^ and SAPS II at sCIV ≤36. Abbreviations: AKI, acute kidney injury; ANOVA, analysis of variance; best global outcome, combination outcome consisting of no AKI, clinical and microbiological treatment success; BMI, body mass index [kg/m^2^]; C72mean, mean vancomycin serum concentration during the first three days of continuous infusion of vancomycin [mg/L], CART, classification and regression tree; CI, confidence interval; CIV, continuous infusion of vancomycin; CoNS, coagulase-negative staphylococci; CRP, C-reactive protein; DOOR, desirability of outcome ranking; eGFR, estimated glomerular filtration rate [mL/min/1.73 m^2^]; LLM, linear mixed model; ROC, receiver-operating characteristic; SAPS, Simplified Acute Physiology Score II; sCIV, start of continuous infusion of vancomycin SCr, serum creatinine. p-value <0.05 was considered significant.

**Table 3.** Primary and secondary outcome parameters.

|  | N | Overall<br>(n = 922<sup>1</sup>) | Subgroup A<br>(n = 262<sup>1</sup>) | Subgroup B<br>(n = 186<sup>1</sup>) | Subgroup C<br>(n = 105<sup>1</sup>) | Subgroup D<br>(n = 369<sup>1</sup>) | p-value<sup>2</sup> |
| --- | --- | --- | --- | --- | --- | --- | --- |
| ICU mortality | 922 | 157 (17) | 79 (30) | 31 (17) | 13 (12) | 34 (9.2) | <0.001 |
| In-hospital mortality | 922 | 225 (24) | 110 (42) | 43 (23) | 21 (20) | 51 (14) | <0.001 |
| 30-day mortality | 922 | 166 (18) | 83 (32) | 27 (15) | 16 (15) | 40 (11) | <0.001 |
| Time to mortality [days] | 225 | 13 (7, 31) | 12 (7, 28) | 17 (10, 39) | 9 (6, 20) | 12 (8, 23) | 0.3 |
| Total AKI | 922 | 173 (19) | 77 (29) | 34 (18) | 16 (15) | 46 (12) | <0.001 |
| Early AKI | 922 | 70 (86) | 33 (13) | 16 (9) | 5 (5) | 16 (4) | <0.001 |
| Late AKI | 922 | 103 (11) | 44 (17) | 18 (10) | 11 (10) | 30 (8) | 0.007 |
| Dialysis | 922 | 17 (2) | 12 (5) | 2 (1) | 3 (3) | 0 (0) | <0.001 |
| AKIN / KDIGO stage 1 | 922 | 20 (2) / 38 (4) | 4 (2) / 13 (5) | 6 (3) / 7 (4) | 1 (1) / 3 (3) | 9 (2) / 15 (4) | 0.6 / 0.9 |
| AKIN / KDIGO stage 2 | 922 | 81 (9) / 63 (7) | 36 (14) / 27 (10) | 15 (8) / 14 (8) | 6 (6) / 4 (4) | 24 (7) / 18 (5) | 0.008 / 0.032 |
| AKIN / KDIGO stage 3 | 922 | 72 (8) / 72 (8) | 37 (14) / 37 (14) | 13 (7) / 13 (7) | 9 (9) / 9 (9) | 13 (4) / 13 (4) | <0.001 / <0.001 |
| Time to total AKI [hours] | 173 | 64 (24, 125) | 55 (24, 125) | 55 (33, 98) | 68 (35, 146) | 85 (26, 162) | 0.5 |
| Time to early AKI [hours] | 70 | 21 (15, 34) | 22 (16, 31) | 32 (18, 41) | 20 (14, 21) | 19 (14, 26) | 0.3 |
| Time to late AKI [hours] | 103 | 117 (70, 167) | 120 (69, 156) | 96 (71, 119) | 107 (68, 153) | 125 (86, 186) | 0.3 |
| Duration of total AKI [days] | 75 | 10 (4, 20) | 11 (4, 17) | 18 (8, 34) | 11 (6, 19) | 6 (3, 13) | 0.3 |
| Duration of early AKI [days] | 35 | 10 (3, 22) | 11 (3, 22) | 15 (8, 24) | 12 (7, 19) | 5 (3, 13) | 0.7 |
| Duration of late AKI [days] | 40 | 10 (4, 16) | 11 (5, 14) | 30 (21, 34) | 11 (6, 16) | 6 (4, 12) | 0.3 |
| No return to SCr at sCIV 28 days after sCIV<sup>3</sup> |  |  |  |  |  |  |  |
| Total AKI | 105 | 26 (25) | 11 (29) | 7 (30) | 2 (22) | 6 (17) | 0.033 |
| Early AKI | 46 | 12 (26) | 5 (31) | 3 (25) | 0 (0) | 4 (27) | 0.7 |
| Late AKI | 59 | 14 (24) | 6 (27) | 4 (36) | 2 (33) | 2 (10) | 0.024 |
| Clinical failure with survival | 922 | 163 (18) | 32 (12) | 23 (12) | 24 (23) | 84 (23) | <0.001 |
| Microbiological failure | 922 | 309 (31) | 87 (33) | 67 (36) | 33 (31) | 122 (33) | 0.9 |

Subgroup A: eGFR at sCIV <90 mL/min/1.73 m<sup>2</sup> and SAPS II at sCIV >36; subgroup B: eGFR at sCIV ≥90 mL/min/1.73 m<sup>2</sup> and SAPS II at sCIV >36; subgroup C: eGFR at sCIV <90 mL/min/1.73 m<sup>2</sup> and SAPS II at sCIV ≤36; subgroup D: eGFR at sCIV ≥90 mL/min/1.73 m<sup>2</sup> and SAPS II at sCIV ≤36.
Abbreviations: AKI, acute kidney injury; AKIN, Acute Kidney Injury Network; eGFR, estimated glomerular filtration rate; ICU, intensive care unit; KDIGO, Kidney Disease Improving Global Outcomes; SAPS, Simplified Acute Physiology Score II; sCIV, start of continuous infusion of vancomycin; SCr, serum creatinine.
<sup>1</sup> n (%); Median (IQR); <sup>2</sup> Kruskal-Wallis rank sum test; Pearson's Chi-squared test; Fisher's exact test; p-value <0.05 was considered significant; <sup>3</sup> only hospital survivors

## 3 Results

### 3.1 Demographic data

During the 12-year study period, 1,741 out of 15,907 ICU patients received vancomycin. 750 did not meet the inclusion criteria and 69 were excluded because of therapy duration or dialysis. Subsequently, 922 patients were included (**Figure 1**). 262 patients were in subgroup A, 186 patients in subgroup B, 105 patients in subgroup C and 369 patients in subgroup D. Their characteristics are shown in **Table 1**, vancomycin therapy in **Table 2**, and outcome parameters in **Table 3**. Patients were treated in neurosurgery (49%), visceral surgery (23%), trauma surgery (13%), thoracic surgery (9%) and vascular surgery (6%), with particularly high proportions of vascular (29/52, 56%) and visceral (88/213, 41%) patients in subgroup A and neurosurgery (225/454, 50%) and trauma (57/122, 47%) patients in subgroup D.

### 3.2 Target ranges

A range of vancomycin serum concentrations that was most favourable for all outcome parameters and subgroups could not be determined due to the heterogeneity of the results. Higher C72mean, especially >25 mg/L, was associated with more mortality and AKI (p<0.05) (**Table 4**, **Figure 3**, and **Table 5**). An exception was 30-day mortality in subgroups B and D (**Table 4**, **Table 5 A/B 3+5** and supplementary material **Table S 20** and **Table S 21**). Clinical failure with survival and microbiological failure were each inconsistently associated with low or high C72mean, respectively (**Table 4** and **Table 5**). Significance was found in subgroup B with more clinical treatment failures at higher concentrations (>25 mg/L vs <20 mg/L: OR 3.20, p=0.037 [**Table 4**, supplementary material **Table S 28**]; ≥24.05 mg/L: OR 3.85, p=0.003 [**Table 5 A/B 3**]) or in subgroup D with more microbiological treatment failures at lower concentrations (<20 mg/L vs 20-25 mg/L: OR 1.82, p=0.032 [**Table 4**, supplementary material **Table S 10**] or <19.43 mg/L: OR 1.99, p=0.005 [**Table 5 A/B 5]**). The best global outcome was achieved in the overall cohort with C72mean <25 mg/L (44% [95% CI, 40-48%] **Figure 3 a**), in subgroup A 15-20 mg/L (44% [95% CI, 28 62%] **Figure 3 b**), in subgroup B <15 mg/L (64% [95% CI, 45-79%] **Figure 3 c**), in subgroup C 20-25 mg/L (56% [95% CI, 35-72%] **Figure 3 d**) and in subgroup D ≤30 mg/L (43% [95% CI, 38-48%]; **Figure 3 e**). Cut-offs yielded a favourable C72mean of 19-24 mg/L for the overall cohort, <26 mg/L or <20 mg/L for subgroup A, <17 mg/L for subgroup B, <24 mg/L for subgroup C and 19-28 mg/L for subgroup D (p<0.05, **Table 5**), which were associated with lower odds of mortality, AKI and clinical and microbiological failure.

**Figure 3.**
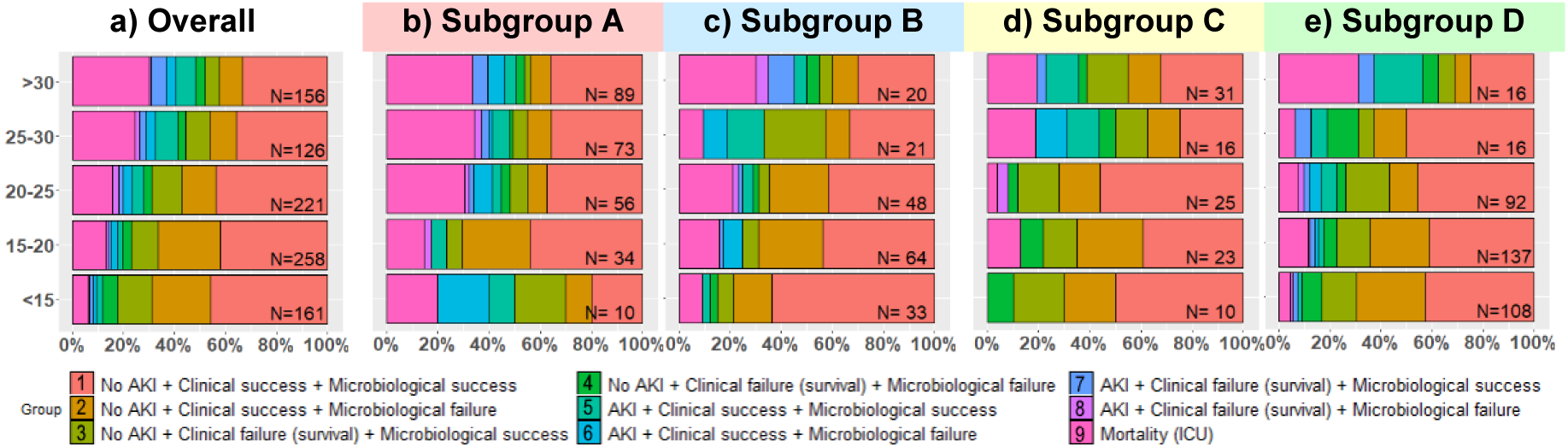
Desirability of outcome ranking (DOOR) analysis by mean vancomycin serum concentration during the first three days of continuous infusion of vancomycin in five concentration groups [y-axis in mg/L]. Each patient was assigned an overall outcome based on ICU mortality, AKI and clinical and microbiological treatment success or failure. 9 outcome level were defined, from (1) most desirable to (9) least desirable (see legend for classification). Overall (n=922); subgroup A (n=262): eGFR at sCIV <90 mL/min/1.73 m^2^ and SAPS II at sCIV >36; subgroup B (n=186): eGFR at sCIV ≥90 mL/min/1.73 m^2^ and SAPS II at sCIV >36; subgroup C (n=105): eGFR at sCIV <90 mL/min/1.73 m^2^ and SAPS II at sCIV ≤36; subgroup D (n=369): eGFR at sCIV ≥90 mL/min/1.73 m^2^ and SAPS II at sCIV ≤36. Abbreviations: AKI, acute kidney injury; eGFR, estimated glomerular filtration rate of vancomycin therapy [mL/min/1.73 m^2^]; ICU, intensive care unit; SAPS, Simplified Acute Physiology Score II; sCIV, start of continuous infusion of vancomycin.

**Table 4.**
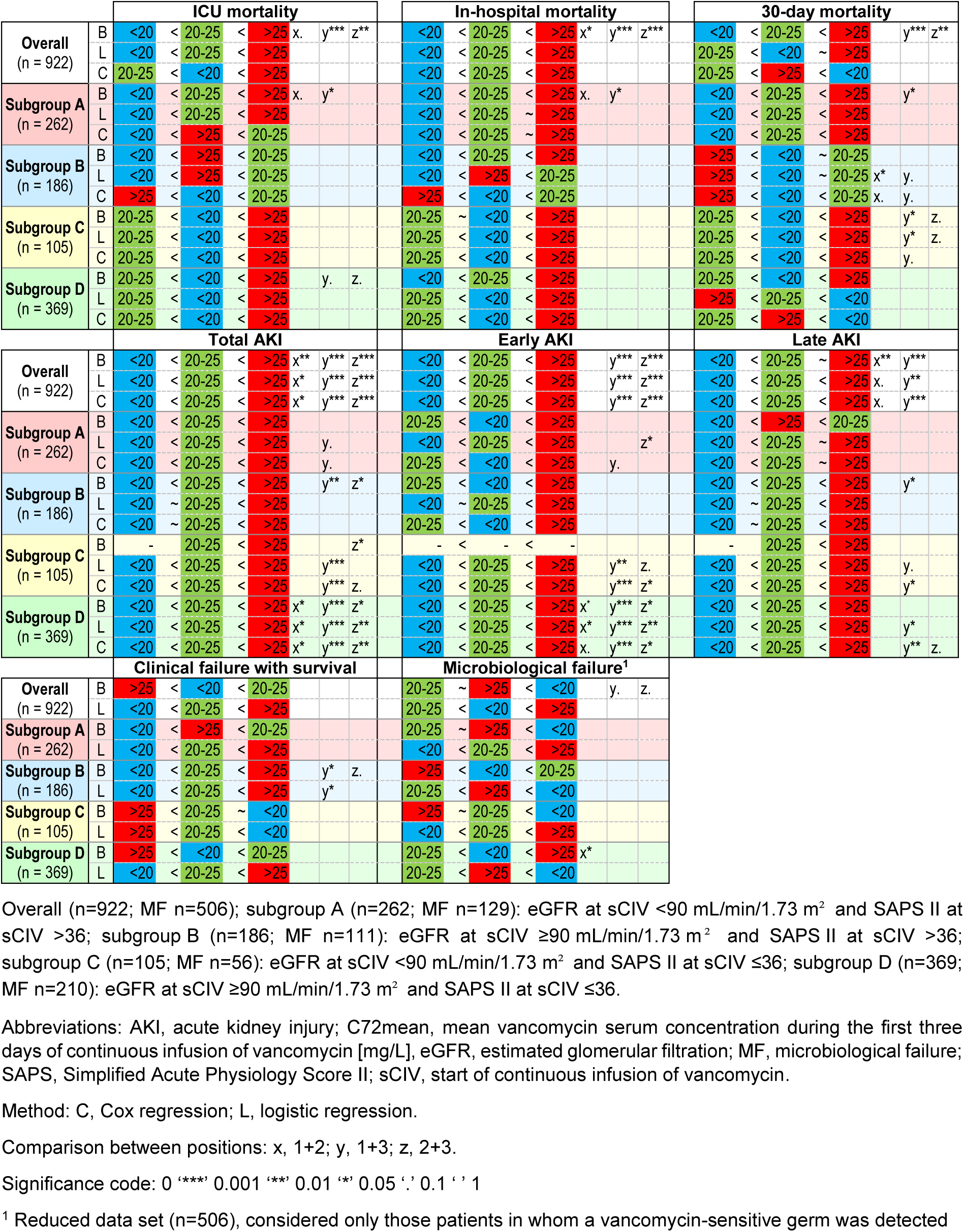
For the outcome parameters, the order of the mean vancomycin serum concentration during the first three days of continuous infusion of vancomycin (C72mean) classes within the overall cohort and the subgroups of estimated glomerular filtration rate (eGFR) and Simplified Acute Physiology Score II (SAPS II) determined by (B) bivariate comparison or post-hoc comparison after logistic (L) or Cox (C) regression analysis, in each case from left with the lowest to right with the highest predicted probability (L) or hazard (C).

**Table 5.**
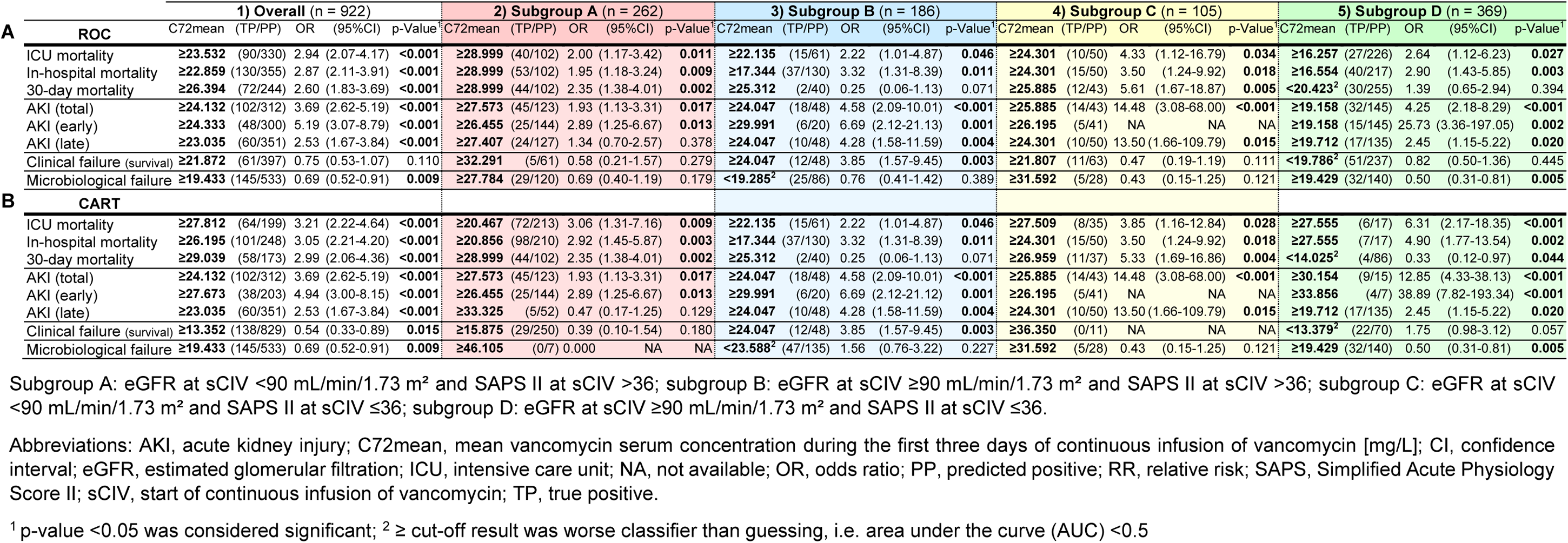
Mean vancomycin serum concentration during the first three days of continuous infusion of vancomycin (C72mean) cut-off values during continuous infusion of vancomycin for therapeutic efficacy and safety determined by (A) receiver-operating characteristic (ROC) and (B) classification and regression tree (CART) analysis.

### 3.3 Predictors of Mortality

C72mean <20 mg/L, eGFR ≥90 mL/min/1.73 m^2^ and body mass index (BMI) were identified as significant negative predictors of (time to) mortality, with a relevant interaction between C72mean and eGFR for 30-day mortality (**Figure 4**, supplementary material **Table S 11**). AKI, the use of vasopressors, cardiovascular comorbidity, the absence of bacterial detection and no reduction in C-reductive protein (CRP) or leukocytes within the first 7 days after start of CIV were significantly associated with higher odds of (time to) mortality.

**Figure 4.**
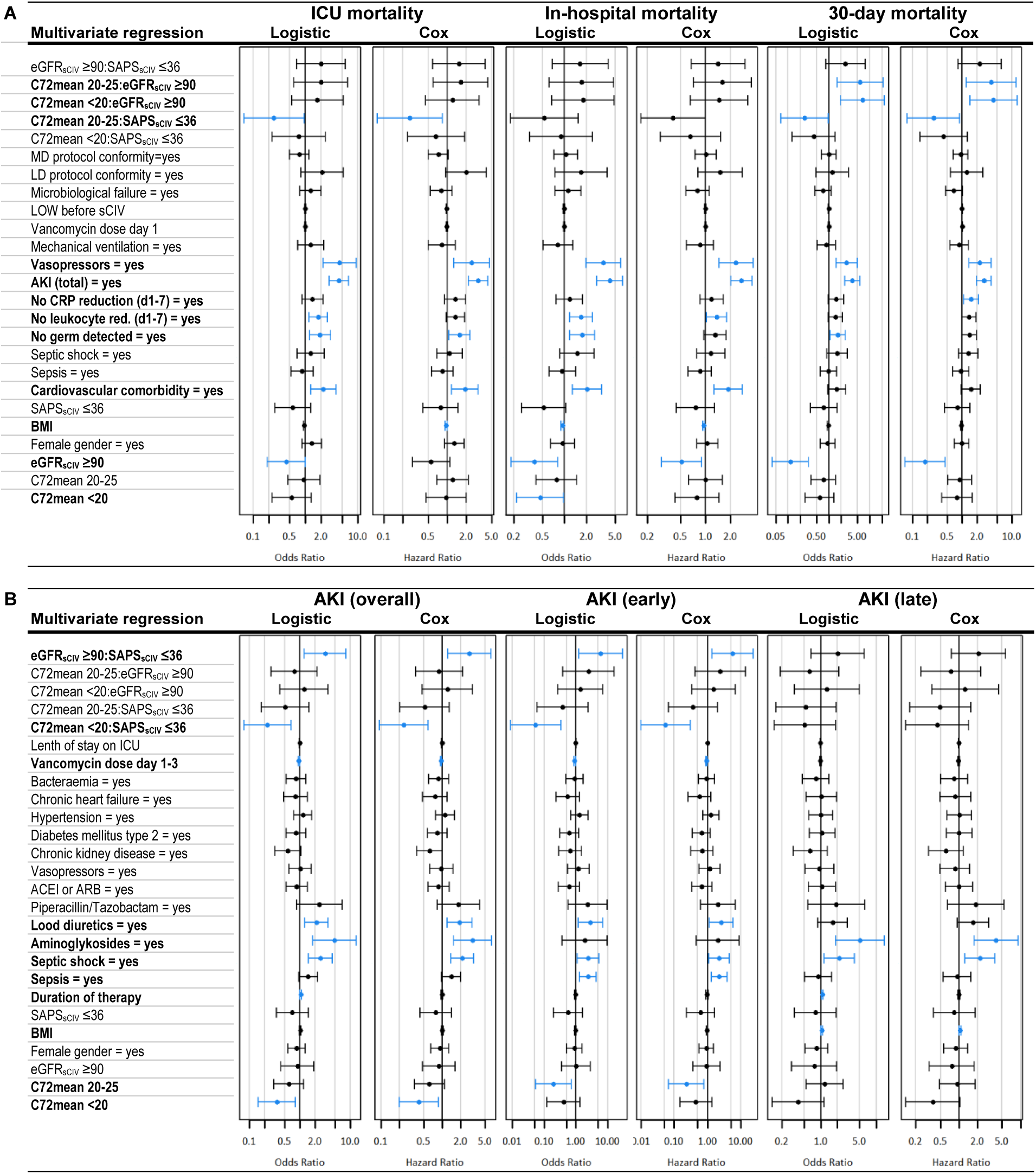
Predictors for (A) mortality (intensive care unit (ICU), hospital, 30-day) and (B) the development of acute kidney injury (AKI; total AKI, early AKI, late AKI) during continuous infusion of vancomycin determined by multivariate logistic and Cox regression analysis (n = 922, no missing values). Blue coloured odds ratios and confidence intervals are significantly different from 1 at the 5% level. Abbreviations: ACEI, angiotensin-converting enzyme inhibitor; AKI, acute kidney injury; ARB, angiotensin receptor blocker; BMI, body mass index [kg/m^2^]; C72mean, mean vancomycin serum concentration during the first three days of continuous infusion of vancomycin [mg/L]; CI, confidence interval; CKD, chronic kindey disease; CRP, C-reactive protein; eGFR_sCIV_, estimated glomerular filtration rate at the start of continuous infusion of vancomycin [mL/min/1.73 m^2^]; HR, hazard ratio; ICU, intensive care unit; OR, odds ratio; SAPS_sCIV_, Simplified Acute Physiology Score II at the start of continuous infusion of vancomycin; sCIV, start of continuous infusion of vancomycin. Reference values: C72mean, >25 mg/L; eGFR_sCIV_, <90 mL/min/1.73 m^2^; SAPS_sCIV_, >36.

### 3.4 Predictors of Acute Kidney Injury

Significant negative predictors of (time to) AKI were C72mean ≤25 mg/L and mean vancomycin dose within the first three days of therapy (**Figure 4**, supplementary material **Table S 12**). Significant positive predictors were duration of CIV, sepsis, septic shock, concomitant use of aminoglycosides or loop diuretics and BMI. There were relevant interactions between C72mean and SAPS.

### 3.5 Predictors of Clinical Treatment Failure with Survival

Female gender was significantly associated with lower clinical failure with survival (OR=0.637 (0.428-0.947), p=0.026), while longer duration of CIV was a significant positive predictor (OR=1.054 (1.017-1.093), p=0.004) (supplementary material **Table S 13**).

### 3.6 Predictors of Microbiological Treatment Failure

Significant positive predictors of microbiological failure were sepsis (OR=1.957 (1.190-3.219), p=0.008), septic shock (OR=3.562 (1.805-7.027), p<0.001), gastrointestinal infection (OR=2.549 (1.328-4.891), p=0.005), pulmonary infection (OR=2.015 (1.279-3.173), p=0.002), bacteraemia with vancomycin-susceptible pathogens (OR=2.031 (1.270-3.246), p=0.003), Enterococcus ssp. (OR=1.903 (1.088-3.328), p=0.024), CoNS (OR=1.758 (1.055-2.928), p=0.030) and duration of CIV (OR=1.05 (1.005-1.097), p=0.030) (supplementary material **Table S 14**).

## 4 Discussion

We investigated how vancomycin serum concentrations during the first three days of CIV (C72mean) were associated with treatment outcome (i.e. mortality, AKI, clinical and microbiological failure) in critically ill patients. Analysis of all 922 consecutive patients revealed inhomogeneities, so we divided the overall cohort into four homogeneous subgroups based on eGFR (</≥90 mL/min/1.73 m^2^) and SAPS II (≤/>36) at CIV initiation. Generally, a higher C72mean was associated with higher mortality, AKI and clinical treatment failure, but less microbiological treatment failure. However, the four subgroups yielded different results: For the two subgroups with SAPS >36 (subgroups A and B), C72mean <20 mg/L (i.e. 15-20 mg/L or <17 mg/L) was associated with the best treatment outcome, indicating that lower concentrations increase the chance of an optimal treatment outcome. For SAPS ≤36 (subgroups C and D), C72mean >19 mg/L (i.e. 20-25 mg/L or 19-28 mg/L) was best, indicating that in this case, higher concentrations increase the chance of an optimal treatment outcome.

There is a paucity of large-scale studies on CIV. Our cohort study is one of the largest, along with that of Hanrahan et al. (653 CIV patients) [22]. To determine the optimal C72mean targets, we considered which study design would be best. Spadaro et al. investigated subgroups with poor and better renal function (creatinine clearance ≤/>50 mL/min), but did not include SAPS as a subgrouping parameter [12]. Other researchers identified renal function and SAPS as predictors of outcome parameters, but did not create subgroups with these variables in their analyses [14]. We expected that subgroups including eGFR and SAPS would eventually provide usable results for everyday clinical practice and help us to make progress in the area of target ranges for CIV. It seemed likely that different targets would be needed for different populations [4, 23]. Apart from logistic regression to determine associations with outcomes such as death, treatment failure or AKI [12–14, 22], to our knowledge none of the techniques we used had previously been used to study vancomycin serum cocentration during CIV. However, some of our methods have been applied in individual IIV studies (supplementary material **Table S 9**). By using these methods in combination, we were able to corroborate our findings (**Figure 2**). Post-hoc tests after ANOVA for our logistic and Cox regression models confirmed our initial results from bivariate comparison. DOOR analyses helped to refine favourable concentration ranges for optimal outcomes. Using CART and ROC thresholding methods, we added more accurate C72mean thresholds to these ranges. This approach allowed us to identify specific target ranges to maximise CIV efficacy and minimise toxicity for each subgroup.

The revised guideline on vancomycin use in severe MRSA infections recommended aiming for 20-25 mg/L for CIV to maintain efficacy but reduce nephrotoxicity [4]. This target range, frequently used in the literature [10], corresponds to a pharmacokinetic/pharmacodynamic (PK/PD) target of the ratio of the area under the curve over 24 hours to the minimum inhibitory concentration of the isolated strain (AUC24/MIC, at MIC of 1 mg/L based on broth microdilution methods) of 480-600 [4]. However, the recommendation for a PK/PD target >400 is based solely on IIV, in vitro and animal studies [4, 24]. Additionally, other investigators have not found better clinical outcomes with AUC24 >400 [25]. Therefore, the guideline authors themselves admitted that their proposed CIV serum range is not considered validated [4]. In addition, the recommendation only referred to infections caused by MRSA. In our cohort with different infections and pathogens (only 6% MRSA, **Table 1**), we showed that the range of 20-25 mg/L was beneficial in healthier critically ill patients (SAPS ≤36, subgroups C and D), but needs to be reconsidered in more severely ill patients (SAPS >36, subgroups A and B). For IIV, the guideline recommended to target AUC24/MIC 400-600, which corresponds to 16.7-25 mg/L for CIV [24]. The lower limit aligned with our favourable concentration range identified for patients with increased disease severity (i.e. SAPS >36).

The reported high mortality rate for critically ill patients with CIV ranged from 8-64% [10] (in our study: ICU mortality 17%, in-hospital mortality 24%, 30-day mortality 18%), so special attention should be paid to possible factors influencing mortality. Like us, other researchers have found higher mortality at higher drug concentrations for both vancomycin [13, 14, 26] and other anti-infectives [27]. It was previously discussed that higher mortality at higher drug concentrations was related to a sicker clientele, with poorer kidney function due to illness leading to higher drug levels [27]. Reduced kidney function also increased the risk of death [28]. Our regression analysis of the whole cohort confirmed that renal function (i.e. eGFR) and disease severity (i.e. SAPS) affected mortality. Therefore, our subgrouping was useful to investigate the association between concentration and mortality. Comparison within the subgroups did not confirm that sicker individuals had a higher mortality rate simply because of their poorer health (supplementary material **Table S 32**), supporting the idea of concentration-related mortality. Additionally, we identified C72mean <20 mg/L as a significantly favourable predictor of mortality in the regression analysis. However, we also found significant interactions between C72mean and eGFR for 30-day mortality. Thus, depending on the cohort, low concentrations (i.e. C72mean <20 mg/L) were associated with higher (for eGFR ≥90 mL/min/1.73 m^2^, subgroups B and D) or lower (for eGFR <90 mL/min/1.73 m^2^, subgroups A and C) mortality. Cohort-dependent effects of drug concentration may also explain why benefits of higher [6, 11, 12], or lower [13, 14, 26] concentrations were reported in the literature. The guideline recommendations that emphasised the importance of early, adequate concentrations [4] therefore have an additional aspect to consider: adequate concentration does not mean the same for all patients, but must be assessed taking into account the patient’s particular health situation. We suggest using our subgroup definition. The question remains as to whether high drug levels are harmful per se or a prognostic factor. We also found, as previously described, AKI [8, 22], the use of vasopressors [29], underlying cardiovascular disease [30], lack of bacterial detection [31], and no reduction in CRP [32] or leukocytes [33] within the first 7 days of CIV were associated with higher mortality.

The concentration-dependent nephrotoxicity of vancomycin, ranging from 2-77% in the literature, is well known [7, 12, 15, 22] and consistent with our results (AKI 12-29%). Vancomycin serum concentrations <24-30 mg/L were significantly beneficial [10, 14, 22]. Impaired renal function (e.g. eGFR <90 mL/min/1.73 m^2^) [12, 13], high severity of illness (e.g. SAPS >36) [12, 14, 34], longer duration of CIV [13, 22, 34], higher BMI [34], sepsis, septic shock [14, 34], concomitant aminoglycosides [34] or loop diuretics [34] were significantly associated with an increased risk of AKI.

Higher mean vancomycin serum concentrations (e.g. >24 mg/L) were associated with more clinical treatment failures in the literature and in our study [26]. In addition, male gender was detrimental, which may be due to existing gender differences [35].

Higher vancomycin concentrations (C72mean >19 mg/L in our study) resulted in fewer microbiological treatment failures [26]. In contrast, we identified CoNS, enterococci, bacteraemia (with 52% CoNS or 28% enterococci) and pneumonia as significantly detrimental. This may be due to the low vancomycin levels in the respective cohorts (C72mean ≤20 mg/L). As recommendations for non-MRSA infections are less well studied [4] and the tissue penetration of vancomycin is described as low [23], the concentration may have been insufficient. However Duszynska et al. showed that 15-20 mg/L eradicated MRSA as other Gram-positives [36]. Microbiological treatment failure had no significant effect on mortality or clinical treatment failure in our study. Therefore, aiming for high serum concentrations (especially >25 mg/L) to eliminate all pathogens does not seem reasonable, as higher concentrations were associated with higher mortality, AKI and clinical failure rates. However, it is important to note that infections were not the only cause of death and clinical failure. Thus, further nuanced research comparing the efficacy of vancomycin concentration classes in vivo against different pathogens is needed to better define the necessary lower limit of the target range of vancomycin serum concentrations during CIV.

## 5 Limitations

This study has several limitations.

First, the data were collected retrospectively in a single centre, so causality cannot be inferred, and the data on mortality, AKI and failure must be treated with caution due to possible confounding. However, retrospective analyses have the advantage of including all patients, even those with low survival rates, to provide a more accurate picture.

Second, due to its use in our clinical routine, we only analysed the total serum vancomycin (bound and free). This approach has a potential for inaccuracy, as we discussed earlier [10].

Third, vancomycin serum concentrations were used, but these do not always reflect the concentration at the site of infection [23].

Fourth, eGFR was determined using the CKD-EPI formula, as CrCl determination does not correspond to the clinical practice at our institution and CKD-EPI has a good overall correlation for vancomycin dosing and in critically ill patients [37].

Fifth, our data did not allow us to focus sufficiently on the infectious genesis of mortality and clinical treatment failure. Consequently, only all-cause mortality and clinical failure, rather than infection-related mortality and clinical failure, were considered. However, it is likely that factors influencing mortality and the development of organ failure, such as the extent of subarachnoid haemorrhage or injury, the inability to rehabilitate an abdomen, or the alteration of treatment goals due to a poor neurological prognosis, played an important role in this context [38, 39].

Sixth, more than 90% of patients received combination antimicrobial therapy. Seventh, only surgical patients were considered.

Finally, different definitions of sepsis were used because there was a long observation period during which accepted international definitions, which were implemented in our ICU, changed.

## 6 Conclusions

In conclusion, retrospective analyses of vancomycin serum concentrations during CIV suggest that ICU patients’ disease severity should be considered when selecting a target concentration. The results challenge the recommendation to use the commonly reported serum concentrations of 20-25 mg/L [4] or 20-30 mg/L [10] as a general predictor of treatment success. Instead, our results emphasise that the target concentration might be sought inversely related to SAPS to achieve the most successful and least toxic therapy. However, confirmation requires prospective, randomised evaluation in vancomycin serum concentration cohorts, adjusting for patients’ initial physical condition and further potential confounders.

## Data Availability

All data produced in the present study are available upon reasonable request to the authors.

## Declarations

### Ethical Approval

The study was conducted in accordance with the Declaration of Helsinki (version 2013) and was approved by the Ethics Committee of the University of Witten Herdecke with a waiver for the need of patient’s informed consent (Chair: Prof. Dr. med. P.W. Gaidzik, No. S 55/2022).

### Informed Consent Statement

Not applicable.

### Sequence Information

Not applicable

## Acknowledgments

We acknowledge Viola Fuchs for her support in supervising the research project. In addition, we express our gratitude to Andreas Plenert and Anna Marina Funk for facilitating access to the PDMS database. We would also like to express our gratitude to Sven Kiebler for providing access to the old hospital information system and to Jérôme Defosse for offering insight into the archived anaesthesia data. Finally, we thank Stephanie Knippschild, Sabrina Voß, Alexander and Renate Viertel for their initial assistance with data analysis.

## Author contributions

K.V. and F.M. conceived of and designed the study. K.V. performed the data curation. K.V., F.M., C.v.M and S.H. analysed the data. K.V. wrote the manuscript with support and critical revision from F.M., C.v.M, S.H. and T.A.. F.M. supervised the project. All authors have reviewed and contributed to the final manuscript. All authors have read and agreed to the published version of the manuscript.

## Declaration of generative AI and AI-assisted technologies in the writing process

During the preparation of this work the author(s) used the free internet translational programme Deepl (www.Deepl.com/Translator) in order to improve the linguistic quality of the manuscript. After using this tool/service, the author(s) reviewed and edited the content as needed and take(s) full responsibility for the content of the publication.

---

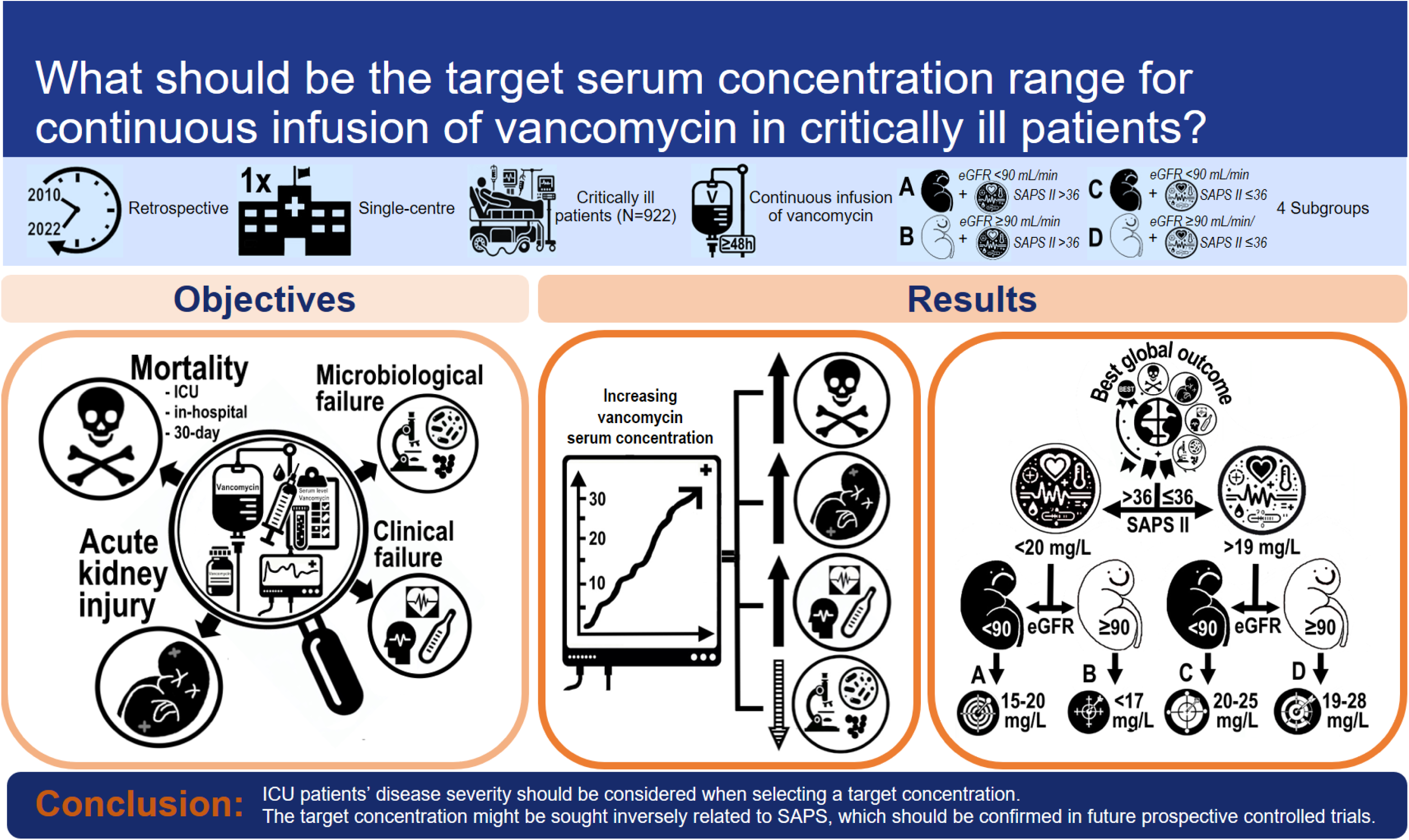

