## Supplementary material for "No one-size-fits-all approach: Retrospective analysis of efficacy and safety of serum concentrations of continuously administered vancomycin in critically ill adults reveals different target serum concentrations depending on disease severity"

### 1.1 Indication for subgroup formation

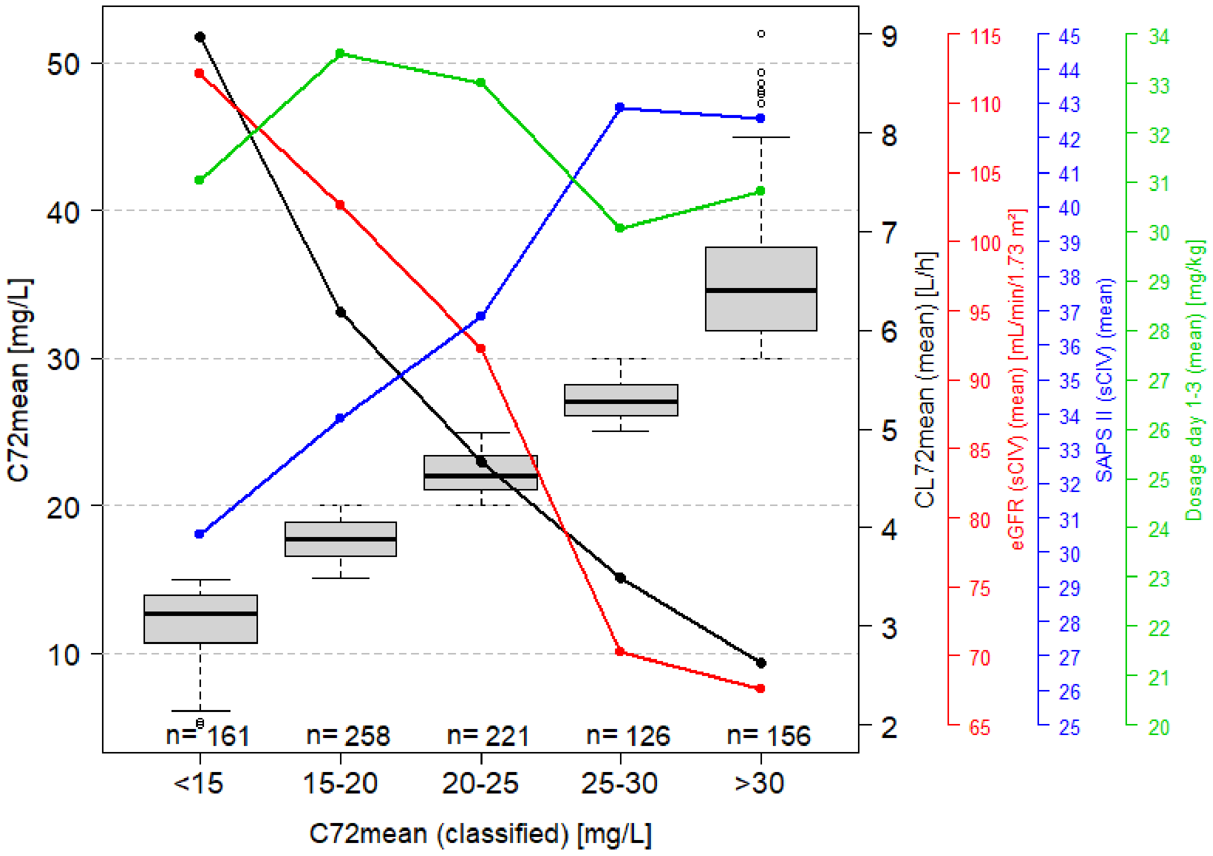


| Characteristics | N | Overall, (n = 922) | <15 mg/L (n = 161) | 15-20 mg/L (n = 258) | 20-25 mg/L (n = 221) | 25-30 mg/L (n = 126) | >30 mg/L (n = 156) | p-value[2] |
|---|---|---|---|---|---|---|---|---|
| ICU mortality | 922 | 157 (17%) | 10 (6%) | 34 (13%) | 35 (16%) | 31 (25%) | 47 (30%) | <0.001 |
| In-hospital mortality | 922 | 225 (24%) | 14 (9%) | 50 (19%) | 52 (24%) | 43 (34%) | 66 (42%) | <0.001 |
| 30-day mortality | 922 | 166 (18%) | 13 (8%) | 41 (16%) | 35 (16%) | 29 (23%) | 48 (31%) | <0.001 |
| AKI (total) | 922 | 173 (19%) | 12 (7%) | 29 (11%) | 41 (19%) | 36 (29%) | 55 (35%) | <0.001 |
| AKI (early) | 922 | 70 (8%) | 5 (3%) | 9 (3%) | 12 (5%) | 16 (13%) | 28 (18%) | <0.001 |
| AKI (late) | 922 | 103 (11%) | 7 (4%) | 20 (8%) | 29 (13%) | 20 (16%) | 27 (17%) | <0.001 |
| Clinical failure (survival) | 922 | 163 (18%) | 34 (21%) | 41 (16%) | 42 (19%) | 21 (17%) | 25 (16%) | 0.6 |
| Microbiological failure | 922 | 282 (31%) | 55 (34%) | 88 (34%) | 61 (28%) | 37 (29%) | 41 (26%) | 0.3 |

**Figure S 1** Mean vancomycin serum concentration during the first three days of continuous infusion of vancomycin (C72mean) (left y-axis, mg/L), C72mean classes (x-axis, box plots (median, interquartile range, and range)), vancomycin clearance over day 1-3 of vancomycin therapy (right black y-axis, black line, L/h) over day 1-3, estimated glomerular filtration rate at the start of continuous infusion of vancomycin (right red y-axis, red line, mL/min/1.73 $m^2$), Simplified Acute Physiology Score II at the start of continuous infusion of vancomycin (right blue y-axis, blue line), mean vancomycin dosage over day 1-3 of vancomycin therapy (right green y-axis, green line, mg/kg total body weight) and corresponding intensive care unit mortality, in-hospital mortality, 30-day mortality, total acute kidney injury (AKI), early AKI, late AKI, clinical failure with survival, microbiological failure (table, n (%)).

Abbreviations: AKI, acute kidney injury; C72mean, mean vancomycin serum concentration during the first three days of continuous infusion of vancomycin; CL72mean, mean vancomycin clearance during the first three days of vancomycin therapy; eGFR, estimated glomerular filtration rate; ICU, intensive care unit; SAPS II, Simplified Acute Physiology Score II; sCIV, start of continuous infusion of vancomycin.

[1] n (%); [2] Pearson's Chi-squared test.

### 1.2 Correlation matrix and regression models

There was a correlation between estimated glomerular filtration rate (eGFR) at the start of continuous infusion of vancomycin, Simplified Acute Physiology Score (SAPS) II at the start of continuous infusion of vancomycin and mean vancomycin serum concentration during the first three days of continuous infusion of vancomycin (C72mean) (**Table S 1** and **Figure S 2**, **Figure S 3**, **Figure S 4**, **Figure S 5**).

**Table S 1** Correlation between estimated glomerular filtration rate at the start of continuous infusion of vancomycin, Simplified Acute Physiology Score II at the start of continuous infusion of vancomycin and mean vancomycin serum concentration during the first three days of continuous infusion of vancomycin.

| Variables | r (Spearman correlation coefficient) | $R^2_{adj}$ (adjusted coefficient of determination) |
|---|---|---|
| $eGFR_{sCIV}$ + C72mean | -0.5598553 | 0.2864 |
| $SAPS_{sCIV}$ + C72mean | 0.3973284 | 0.1365 |
| $SAPS_{sCIV}$ + $eGFR_{sCIV}$ | -0.5579495 | 0.2844 |

Abbreviations: C72mean, mean vancomycin serum concentration during the first three days of continuous infusion of vancomycin [mg/L]; $eGFR_{sCIV}$, estimated glomerular filtration rate at the start of continuous infusion of vancomycin [mL/min/1.73 m²]; $SAPS_{sCIV}$, Simplified Acute Physiology Score II at the start of continuous infusion of vancomycin.

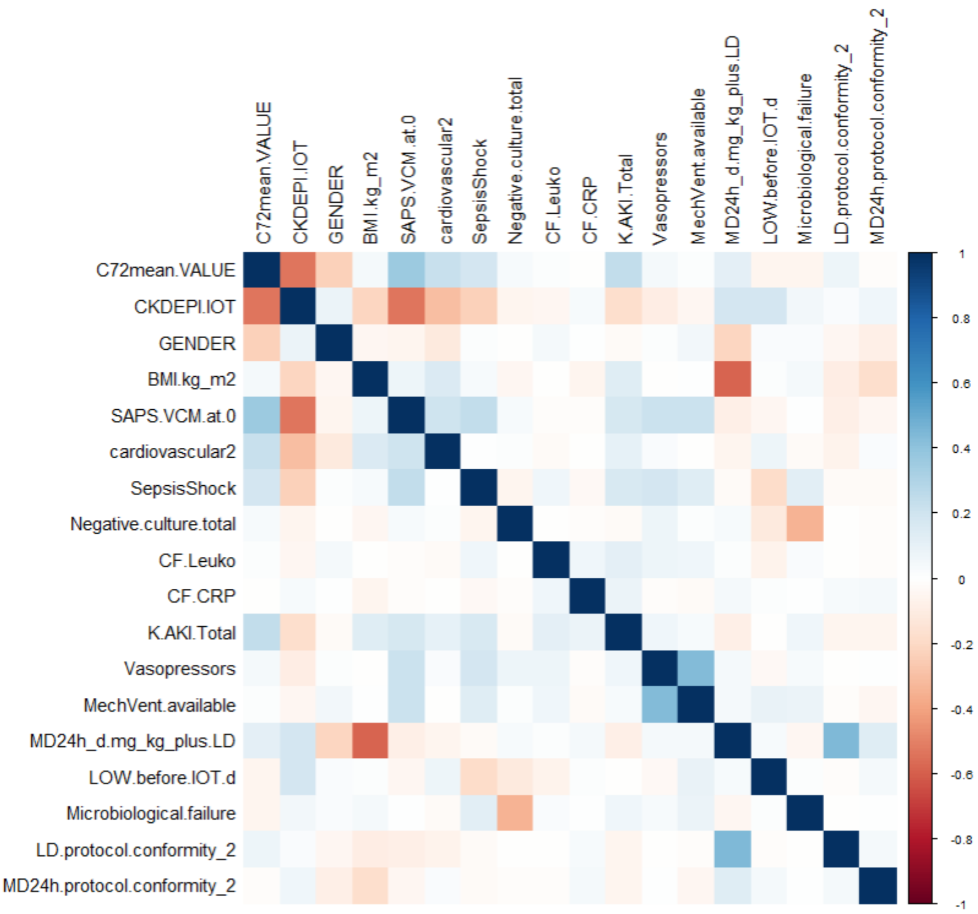


**Figure S 2** Correlation matrix with Pearson correlation coefficient of the variables for the mortality models.

Abbreviations: BMI.kg_m2, body mass index [kg/m²]; C72mean.VALUE, mean vancomycin serum concentration during the first three days of continuous infusion of vancomycin; cardiovascular2, cardiovascular comorbidity; CF.CRP, no C-reactive protein reduction within first seven days of vancomycin therapy; CF.Leuko, no leukocyte reduction within first seven days of vancomycin therapy; CKDEPI.IOT, estimated glomerular filtration rate at the start of continuous infusion of vancomycin ; K.AKI.Total, total acute kidney injury; LD.protocol.conformity_2, loading dose according to protocol; LOW.before.IOT.d, length of stay in the intensive care unit before initiation of vancomycin therapy; MD24h.protocol.conformity_2, initial maintenance dose according to protocol; MD24h_d.mg_kg_plus.LD, vancomycin dose day 1 [mg/kg total body weight]; MechVent.available, availability of mechanical ventilation; Negative.culture.total, negative culture (no germ detected); SAPS.VCM.at.0, Simplified Acute Physiology Score II at the start of continuous infusion of vancomycin; SepsisShock, sepsis or septic shock;

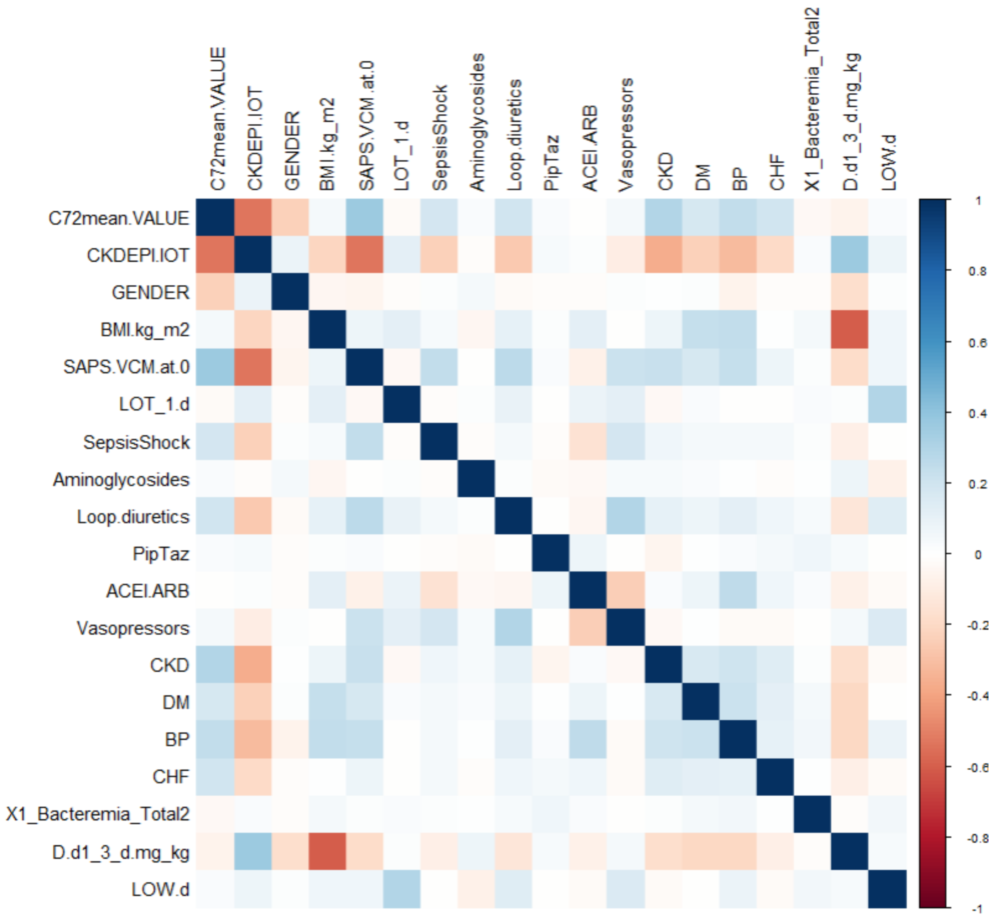


**Figure S 3** Correlation matrix with Pearson correlation coefficient of the variables for the renal insufficiency models.

Abbreviations: ACEI.ARB, Angiotensin-converting enzyme inhibitor or angiotensin receptor blocker; BMI.kg_m2, body mass index [kg/m²]; BP, hypertension; C72mean.VALUE, mean vancomycin serum concentration during the first three days of continuous infusion of vancomycin; CHF, congestive heart failure; CKD, chronic kidney disease; CKDEPI.IOT, estimated glomerular filtration rate at the start of continuous infusion of vancomycin; D.d1_3_d.mg_kg, vancomycin dose day 1-3 [mg/kg total body weight/day]; DM, diabetes mellitus; LOT_1.d, duration of vancomycin therapy [days]; LOW.d, length of stay in the intensive care unit [days]; PipTaz, piperacillin/Tazobactam; SAPS.VCM.at.0, Simplified Acute Physiology Score II at the start of continuous infusion of vancomycin; SepsisShock, sepsis or septic shock; X1_Bacteremia_Total2, bacteraemia.

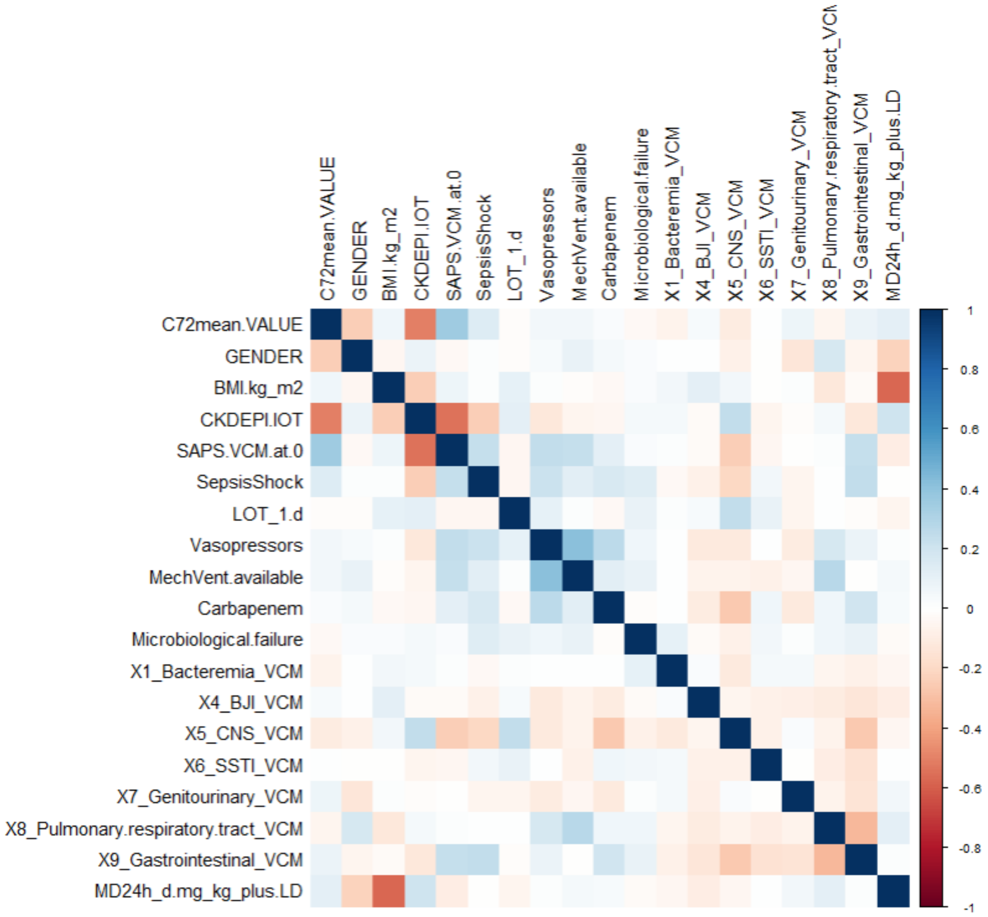


**Figure S 4** Correlation matrix with Pearson correlation coefficient of the variables for the clinical failure with survival model.

Abbreviations: BMI.kg_m2, body mass index [kg/m²]; C72mean.VALUE, mean vancomycin serum concentration during the first three days of continuous infusion of vancomycin; CKDEPI.IOT, estimated glomerular filtration rate at the start of continuous infusion of vancomycin; LOT_1.d, duration of vancomycin therapy [days]; MD24h_d.mg_kg_plus.LD, vancomycin dose day 1 [mg/kg total body weight]; MechVent.available, availability of mechanical ventilation; SAPS.VCM.at.0, Simplified Acute Physiology Score II at the start of continuous infusion of vancomycin; SepsisShock, sepsis or septic shock; X1_Bacteremia_VCM, bacteraemia with vancomycin-sensitive germs; X4_BJI_VCM, bone and joint infection; X5_CNS_VCM, central nervous system infection; X6_SSTI_VCM, skin and soft tissue infection; X7_Genitourinary_VCM, genitourinary infection; X8_Pulmonary.respiratory.tract_VCM, pulmonary infection; X9_Gastrointenstinal_VCM, gastrointestinal infection.

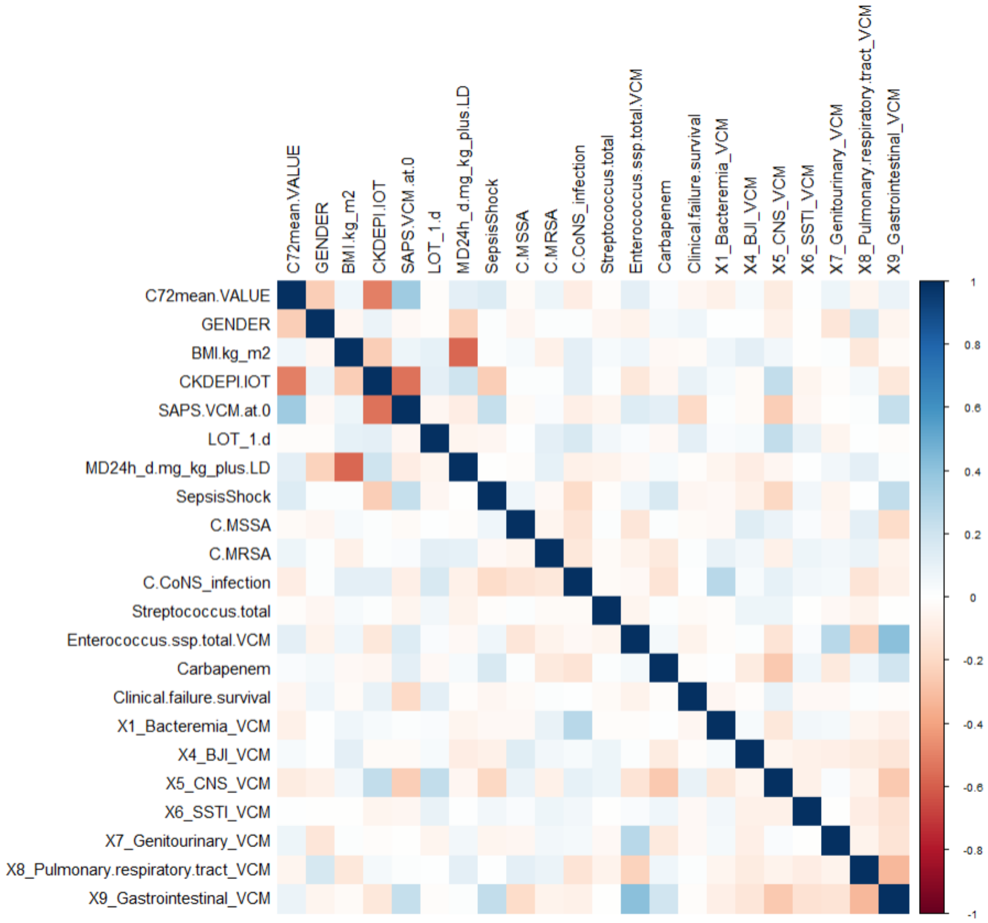


**Figure S 5** Correlation matrix with Pearson correlation coefficient of the variables for the microbiological failure model.

Abbreviations: BMI.kg_m2, body mass index [kg/m²]; C.CoNS_infection, coagulase-negative staphylococci; C.MRSA, methicillin-resistant *Staphylococcus aureus*; C.MSSA, methicillin-sensitive *Staphylococcus aureus*; C72mean.VALUE, mean vancomycin serum concentration during the first three days of continuous infusion of vancomycin; CKDEPI.IOT, estimated glomerular filtration rate at the start of continuous infusion of vancomycin; Enterococcus.ssp.total.VCM, enterococci; LOT_1.d, duration of vancomycin therapy [days]; MD24h_d.mg_kg_plus.LD, vancomycin dose day 1 [mg/kg total body weight]; SAPS.VCM.at.0, Simplified Acute Physiology Score II at the start of continuous infusion of vancomycin; SepsisShock, sepsis or septic shock; Streptococcus.total, streptococci; X1_Bacteremia_VCM, bacteraemia with vancomycin-sensitive germs; X4_BJI_VCM, bone and joint infection; X5_CNS_VCM, central nervous system infection; X6_SSTI_VCM, skin and soft tissue infection; X7_Genitourinary_VCM, genitourinary infection; X8_Pulmonary.respiratory.tract_VCM, pulmonary infection; X9_Gastrointenstinal_VCM, gastrointestinal infection.

**Table S 2** Interpretation of the stronger correlations that could be determined from the correlation matrices.

| Correlation | Variables and interpretation |
|---|---|
| Negative | **C72mean.VALUE + CKDEPI.IOT**<br>Meaning: With better kidney function, the serum level is lower<br>Reliable, as more active substance is excreted with good kidney function. |
| Positive | **SAPS.VCM.at.0 + C72mean.VALUE**<br>Meaning: The vancomycin serum level was higher with higher SAPS.<br>Reliable, as excretion is supposedly worse with greater illness. |
| Negative | **SAPS.VCM.at.0 + CKDEPI.IOT**<br>Meaning: Kidney function was lower with higher SAPS.<br>Reliable, as excretion is supposedly worse with greater disease. |
| Negative | **CKD + CKDEPI.IOT**<br>Meaning: With a history of CKD (no=1, yes=2), renal function was lower at the start of therapy.<br>Reliable, as this is the definition of CKD |
| Negative | **cadiovascular2 + CKDEPI.IOT**<br>Meaning: With underlying cardiovascular disease, renal function was worse.<br>Reliable, as the renal vessels are also affected if the vascular structure is impaired. |
| Positive | **MechVent.available + Vasopressors**<br>Meaning: Vasopressors were often used with artificial ventilation.<br>Reliable, as blood pressure often has to be maintained with vasopressors when artificial ventilation is required in severe illnesses. |
| Negative | **MD24h_d.mg_kg_plus.LD + BMI.kg_m2**<br>Meaning: Dosage in mg/kg total body weight on the first day of therapy was lower for high BMI, less dose was given per kg bw<br>Reliable, as a standard dose was often administered, although the home guideline recommended otherwise, so that the dose per kg body weight was lower in heavier patients. |
| Negative | **Microbiological failure + Negative.culture.total**<br>Meaning: Microbiological treatment failure was greater if no culture was found.<br>Reliable (a little): the variable "Microbiological.failure" also includes "escalation" and "de-escalation" as well as "return to the intensive care unit". If no germ was detected, it could not be treated in a targeted manner and the patient had to be escalated more frequently or readmitted to the ICU |
| Positive | **LD.protocol.conformity_2** (=the LD specified in the in-house guideline was adhered to) **+ MD24h_d.mg_kg_plus.LD** (=dosage in mg/kg total body weight on the first day of therapy)<br>Meaning: If the LD specified in the house guideline was adhered to, the dose on the first day of therapy was higher.<br>Reliable: If the house guideline was followed, a higher LD was administered to heavier patients (i.e. according to BW). If only the "standard dose" was used, this was naturally lower per kg bw for patients with more body weight. |
| Negative | **D.d1_3_d.mg_kg + BMI.kg_m2**<br>Meaning: Dosage in mg/kg total body weight on the first three days was lower with a high BMI, less was dosed per kg bw.<br>Reliable, as a standard dose was often administered, although the home guideline recommended otherwise, so that the dose per kg body weight was lower in heavier patients. |

Abbreviations: BMI.kg_m2, body mass index [kg/m²]; C72mean.VALUE, mean vancomycin serum concentration during the first three days of continuous infusion of vancomycin; cardiovascular2, cardiovascular comorbidity; CKD, chronic kidney disease; CKDEPI.IOT, estimated glomerular filtration rate at the start of continuous infusion of vancomycin; D.d1_3_d.mg_kg, vancomycin dose day 1-3 [mg/kg total body weight/day]; LD.protocol.conformity_2, loading dose according to protocol; MD24h_d.mg_kg_plus.LD, vancomycin dose day 1 [mg/kg total body weight]; MechVent.available, availability of mechanical ventilation; Negative.culture.total, negative culture (no germ detected); SAPS.VCM.at.0, Simplified Acute Physiology Score II at the start of continuous infusion of vancomycin.

#### 1.3 Classification

In the following analysis of the outcome parameters (mortality, AKI, clinical failure with survival, microbiological failure), the variables estimated glomerular filtration rate (eGFR) at the start of continuous infusion of vancomycin, Simplified Acute Physiology Score (SAPS) II at the start of continuous infusion of vancomycin and mean vancomycin serum concentration during the first three days of continuous infusion of vancomycin (C72mean) were analysed in a classified manner (**Table S 3**). SAPS and eGFR were each divided into two classes. A median split (median=36) was used for SAPS, while eGFR was divided into good (eGFR ≥90 mL/min/1.73 m²) and poor (eGFR <90 mL/min/1.73 m²). Three classes were defined for drug concentration [mg/L]: low (<20), optimal ([20, 25]) and high (>25).

**Table S 3** Classified analysis of the frequencies of the variables estimated glomerular filtration rate at the start of continuous infusion of vancomycin (A), Simplified Acute Physiology Score II at the start of continuous infusion of vancomycin (B) and mean vancomycin serum concentration during the first three days of continuous infusion of vancomycin (C).

**a)**

| **$eGFR_{sCIV}$** | Sum |
|---|---|
| < 90 | 367 (39.8%) |
| ≥ 90 | 555 (60.2%) |
| Sum | 922 (100.0%) |

**b)**

| **$SAPS_{sCIV}$** | Sum |
|---|---|
| ≤ 36 | 474 (51.4%) |
| > 36 | 448 (48.6%) |
| Sum | 992 (100.0%) |

**c)**

| **C72mean** | Sum |
|---|---|
| < 20 | 419 (45.4%) |
| [20, 25] | 221 (24.0%) |
| > 25 | 282 (30.6%) |
| Sum | 922 (100.0%) |

Abbreviations: C72mean, mean vancomycin serum concentration during the first three days of continuous infusion of vancomycin [mg/L]; $eGFR_{sCIV}$, estimated glomerular filtration rate at the start of continuous infusion of vancomycin [mL/min/1.73 m²]; $SAPS_{sCIV}$, Simplified Acute Physiology Score II at the start of continuous infusion of vancomycin.

### 1.4 Patient Eligibility and Data Collection

Eligible patients were identified using the electronic patient data management system (PDMS) "Integrated Care Manager (ICM)" (Drägerwerk AG & Co. KGaA, version 12.01 build 0122, database version 46.0 (Rev. 1162)) with a query using the keywords "Vancomycin perf.", i.e. CIV. Only the data of the first CIV cycle of a patient were included in the analysis. Patient and laboratory data were retrieved from the PDMS, the hospital information system ClinicCentre (i-SOLUTIONS Health GmbH, version 22.04. (00.01)), the hospital information system RadCentre (i-SOLUTIONS Health GmbH, Version 35.0.9350.0), the anaesthesia software MEDLINQ (MEDLINQ Softwaresysteme GmbH, Version 2.21.45) and the hospital hygiene analysis system HyBASE Statistik (epiNET AG, V.6.2022.02.30, database version 1063). A data merge was performed using Microsoft Access and Microsoft Excel (Microsoft Corporation, Microsoft Office Professional Plus 2016 (16.0.5317.1000)).

##### *1.4.1 Parameters extracted*

**Table S 4** List of data collected from electronic patient data management and documentation systems.

| | |
|---|---|
| **Demographic characteristics** | Age, gender, height, total body weight, body mass index, length of hospital stay (total, at initiation and after termination of vancomycin therapy), associated clinic, reason for surgery |
| **Medical history** | Comorbidities |
| **ICU** | Severity of illness at baseline (= start of CIV) and within first week of CIV (Sepsis-related Organ Failure Assessment score, Simplified Acute Physiology Score II, Therapeutic Intervention Scoring System), length of mechanical ventilation, length of stay (total, at initiation and after termination of vancomycin therapy), mortality |
| **Concomitant medications** | Concomitant nephrotoxins (aminoglycosides, angiotensin-converting enzyme inhibitors, angiotensin II receptor blockers, diuretics, piperacillin/tazobactam, vasopressors, chemotherapy, nonsteroidal anti-inflammatory drugs, aciclovir, calcineurin inhibitors, colistin), anti-infective therapy administered during CIV |
| **Infection** | Type of infection, sepsis, septic shock. Sepsis was defined according to the criteria valid at the time of treatment |
| **Microbiology** | Microbiological cultures taken, pathogens found, vancomycin minimal inhibitory concentration |
| **Infection laboratory** | Leukocytes, C-reactive protein |
| **Renal function** | Previous stable serum creatinine, serum creatinine for 28 days after termination of CIV or until hospital discharge, creatinine clearance for 28 days after termination of CIV or until hospital discharge (Cockcroft-Gault formula [1] with total body weight at BMI <18.5 $kg/m^2$, with ideal body weight at BMI 18.5-30 $kg/m^2$ and adjusted body weight at BMI >30 $kg/m^2$), estimated glomerular filtration for 28 days after termination of CIV or until hospital discharge (2021 CKD-EPI formula [2]), daily urine output determined at midnight until ICU discharge, renal replacement therapy until hospital discharge |
| **Vancomycin** | Loading dose, regimen of CIV including changes and bolus applications, daily morning serum level check (exclusion of levels obtained after the end of CIV, levels <1 mg/L during running CIV or exceptionally high levels followed by low levels measured on the same day of CIV without conduct of dialysis, as the probability of measurement errors was high).<br>The following vancomycin serum levels were determined: the vancomycin concentration after the first day of CIV (C24, 16-<40 h after CIV initiation) [3], on the second day of CIV (C48, 40-<64 h after CIV initiation), on the third day of CIV (C72, 64-<88 h after the start of CIV), the mean Css of C24, C48 and C72 (C72mean, 16-<88 h after the start of CIV), the lowest Css within the first three days of CIV (C72mean.LOWEST), the highest Css within the first three days of CIV (C72mean.HIGHEST), the mean Css of the entire cycle of CIV (Cmean), the lowest Css within the entire cycle of CIV (Cmean.LOWEST), the highest Css within the entire cycle of CIV (Cmean.HIGHEST) |

Abbreviations: BMI, body mass index; CKD-EPI, Chronic Kidney Disease Epidemiology Collaboration; CIV, continuous infusion of vancomycin; Css, steady-state serum concentration.

### 1.5 Objectives and assessment criteria

The efficacy and safety of steady state serum concentrations during CIV were evaluated using primary and secondary endpoints.

#### 1.5.1 Primary outcome

##### 1.5.1.1 Efficacy

The primary efficacy endpoint was all cause mortality in the ICU. In addition, all cause in-hospital mortality and in-hospital mortality at day 30 after CIV initiation were evaluated.

##### 1.5.1.2 Safety

The primary safety endpoint was the incidence of nephrotoxicity, defined as new-onset of acute kidney injury (AKI) according to AKIN [4] or KDIGO [5] criteria (supplementary material **Table S 5**), between the start of treatment until three days after the end of CIV, distinguishing between early AKI (within the first 48 hours after the start of CIV) and late AKI (48 hours after the start to 72 hours after the end of CIV) [6]. The diagnostic criteria for AKI were an increase in baseline SCr (where baseline=SCr at the start of CIV) of at least 50% within seven days, an increase in SCr of ≥0.3 mg/dL within 48 hours, urine output <0.5 mL/kg TBW/h for 6-12 hours, or initiation of dialysis.

#### 1.5.2 Secondary outcomes

##### 1.5.2.1 Efficacy

Secondary markers of effectiveness were treatment failure and treatment success.

Treatment failure was classified as clinical or microbiological failure.

Clinical failure was defined as persistence, worsening (= progression) or recurrence of baseline symptoms (including the infectious laboratory parameters CRP or leukocytes, Sepsis-related Organ Failure Assessment (SOFA) score [7], SAPS II [8], Therapeutic Intervention Scoring System (TISS) [9]) after seven days of treatment consistent with active infection. Microbiological failure was defined as persistence or spread (= progression) of the same Gram-positive organisms in laboratory samples after three or more days of treatment following a positive culture, a (necessary) switch from vancomycin to a new antibiotic with a spectrum similar to vancomycin (e.g. linezolid, daptomycin or ceftaroline) due to lack of response (= escalation), superinfection, recurrence (= relapse) of a Gram-positive infection five or more days after a interim negative microbiological sample, or repeated vancomycin therapy within 14 days after completion of the first cycle of CIV.

Therapeutic success was divided into clinical and microbiological success. Clinical success was defined as cure or improvement in clinical signs and symptoms of infection (e.g. reduction in elevated leukocyte counts, etc.). Microbiological success was defined as a negative culture of the same type of biological specimen (e.g. blood, intra-abdominal fluids, tissue, cerebrospinal fluid (CSF), etc.) after three or more days of therapy or 14 days after discontinuation of CIV following an initial positive culture, de-escalation of antibiotic therapy, and no need for antibiotic escalation or additional antibiotic therapy.

Risk factors for mortality, clinical and microbiological failure during CIV were also assessed.

##### 1.5.2.2 Safety

Secondary safety endpoints included time to onset of AKI, frequency of different AKI stages, reversibility of AKI at hospital discharge, change in renal function between the start of CIV and day 28, and risk factors for the development of AKI.

#### *1.5.3 AKI Definition*

**Table S 5** Definition of acute kidney injury (AKI)

| | | Serum creatinine | RRT | Urine output |
|---|---|---|---|---|
| **AKIN** (2007) [4] | **1** | SCr ↑ 1.5-2.0 x baseline SCr (7 d) *or* SCr ↑ ≥0.3 mg/dL (48 h) | | <0.5 mL/kg/h for >6 h |
| | **2** | SCr ↑ 2.0-3.0 x baseline SCr (7 d) *or* SCr ↑ ≥0.5 mg/dL | | <0.5 mL/kg/h for >12 h |
| | **3** | SCr ↑ >3.0 x baseline SCr (7 d) *or* acute SCr ↑ ≥0.5 mg/dL if SCr is ≥4 mg/dL | *or* initiation of RRT | <0.3 mL/kg/h for ≥24 h *or* anuria for ≥12 h |
| **KDIGO** (2012) [4, 5] | **1** | SCr ↑ 1.5-1.9 x baseline SCr (7 d) *or* SCr ↑ ≥0.3 mg/dL (48 h) | | <0.5 mL/kg/h for 6-12 h |
| | **2** | SCr ↑ 2.0-2.9 x baseline SCr (7 d) | | <0.5 mL/kg/h for ≥12 h |
| | **3** | SCr ↑ ≥3.0 x baseline SCr (7 d) *or* SCr ↑ ≥4.0 mg/dL | *or* initiation of RRT | <0.3 mL/kg/h for ≥24 h *or* anuria for ≥12 h |

Abbreviations: AKIN, Acute Kidney Injury Network; KDIGO, Kidney Disease Improving Global Outcomes; RRT, renal replacement therapy; SCr, serum creatinine.

#### 1.6 Dosing protocol

**Table S 6** Dosing of the standard vancomycin perfusor.

| kg TBW | Bolus<br>30 mg/kg TBW [g] | Maintenance dose<br>30 mg/kg TBW/d [g/d] | At 40 mg/mL (2 g/50 mL)<br>corresponding to a rate of [mL/h] |
|---|---|---|---|
| **50** | 1.5 | 1.5 | 1.6 |
| **60** | 1.75 | 1.8 | 1.9 |
| **70** | 2.0 | 2.1 | 2.2 |
| **80** | 2.5 | 2.4 | 2.5 |
| **90** | 2.75 | 2.7 | 2.8 |
| **100** | 3.0 | 3.0 | 3.1 |
| **120** | 3.5 | 3.6 | 3.8 |

Abbreviations: kg, kilogram; TBW, total body weight.

**Table S 7** Dosing of the vancomycin perfusor in impaired renal function.

| eGFR | 20-60 mL/min | | <20 mL/min |
|---|---|---|---|
| **kg TBW** | **Maintenance dose<br>20 mg/kg TBW/d [g/d]** | **At 40 mg/mL (2 g/50 mL)<br>corresponding to a rate of [mL/h]** | |
| **50** | 1.0 | 1.0 | |
| **60** | 1.2 | 1.3 | |
| **70** | 1.4 | 1.5 | |
| **80** | 1.6 | 1.7 | **No perfusor** |
| **90** | 1.8 | 1.9 | |
| **100** | 2.0 | 2.1 | |
| **120** | 2.4 | 2.5 | |

Abbreviations: eGFR, estimated glomerular filtration rate; kg, kilogram; TBW, total body weight.

**Table S 8** Rate adjustment of vancomycin perfusor (target range: 20-30 mg/L)

| Measured level [mg/L] | Bolus [mg] | Pause of Perfusor | Change of rate [mL/h] |
|---|---|---|---|
| **<20** | 500 | No pause | +0.5 |
| **20-30** | **Target range** | | |
| **30-40** | No bolus | 4 hours | -0.5 |
| **>40** | No bolus | 12 hours | -1.0 |

### 1.7 Method overview

The following is an overview of the methods employed in this study, together with details of the sources used.

**Table S 9** Overview of methods with sources.

| Method | Source(s) |
|---|---|
| **Logistic regression**<br>- Logistic regression was used to model the odds of mortality, developing renal insufficiency and clinical or microbiological treatment failure as a function of various influencing variables. | [10] |
| **Cox regression**<br>- Cox regression was used to model the (censored) survival time and the time to onset of acute renal failure as a function of several predictor variables. | [11, 12] |
| **Desirability of outcome ranking (DOOR)**<br>- DOOR analysis was performed to show the associations between five C72mean classes (<15, 15-<20, 20-<25, 25-30, >30 mg/L) and overall patient outcome. | [13-15] |
| **Receiver-operating characteristic (ROC)**<br>- ROC analyses were used to identify alternative optimal C72mean ranges associated with the outcome parameters. | [16] |
| **Classification and regression tree (CART)**<br>- CART analyses were used to identify alternative optimal C72mean ranges associated with the outcome parameters. | [17] |
| Some of our methods have been applied in individual studies with intermittent infusion of vancomycin:<br>- **Post-hoc tests after ANOVA** for our logistic and Cox regression models<br>- confirmed our initial results from **bivariate comparison**.<br>- **Desirability of outcome ranking (DOOR)** analyses helped to refine favourable concentration ranges for optimal outcomes.<br>- Using **classification and regression tree (CART)** thresholding methods, we added C72mean thresholds to these ranges. | <br><br>[18-21]<br><br>[22]<br>[18]<br><br>[22] |

### 1.8 Bivariate analysis

**Table S 10** Bivariate comparisons between vancomycin serum concentration during the first three days of vancomycin therapy (C72mean) and treatment outcomes (intensive care unit mortality, in-hospital mortality, 30-day mortality, total acute kidney injury, early acute kidney injury, late acute kidney injury, clinical failure with survival, microbiological failure).

**Overall (n = 922)**

[[1]]

| | C72mean | ICU.MORTALITY | comparison | RR (95% CI) | p-value[1] | OR (95% CI) | p-value[1] |
|---|---|---|---|---|---|---|---|
| 1 | < 20 (n = 419) | 44 (10.50%) | > 25 vs. [20, 25] | 1.75 (1.22, 2.50) | **0.0023** | 2.03 (1.30, 3.17) | **0.0018** |
| 2 | [20, 25] (n = 221) | 35 (15.84%) | > 25 vs. < 20 | 2.63 (1.88, 3.69) | **<0.001** | 3.26 (2.17, 4.90) | **<0.001** |
| 3 | > 25 (n = 282) | 78 (27.66%) | [20, 25] vs. < 20 | 1.51 (1.00, 2.28) | 0.0511 | 1.60 (0.99, 2.59) | 0.0525 |

[[2]]

| | C72mean | HOSPITAL.MORTALITY | comparison | RR (95% CI) | p-value[1] | OR (95% CI) | p-value[1] |
|---|---|---|---|---|---|---|---|
| 1 | < 20 (n = 419) | 64 (15.27%) | > 25 vs. [20, 25] | 1.64 (1.24, 2.17) | **<0.001** | 2.05 (1.38, 3.03) | **<0.001** |
| 2 | [20, 25] (n = 221) | 52 (23.53%) | > 25 vs. < 20 | 2.53 (1.93, 3.31) | **<0.001** | 3.49 (2.44, 5.00) | **<0.001** |
| 3 | > 25 (n = 282) | 109 (38.65%) | [20, 25] vs. < 20 | 1.54 (1.11, 2.14) | **0.0097** | 1.71 (1.13, 2.57) | **0.0105** |

[[3]]

| | C72mean | 30D.MORTALITY | comparison | RR (95% CI) | p-value[1] | OR (95% CI) | p-value[1] |
|---|---|---|---|---|---|---|---|
| 1 | < 20 (n = 419) | 54 (12.89%) | > 25 vs. [20, 25] | 1.72 (1.20, 2.47) | **0.0029** | 2.00 (1.28, 3.12) | **0.0024** |
| 2 | [20, 25] (n = 221) | 35 (15.84%) | > 25 vs. < 20 | 2.12 (1.55, 2.90) | **<0.001** | 2.54 (1.72, 3.74) | **<0.001** |
| 3 | > 25 (n = 282) | 77 (27.30%) | [20, 25] vs. < 20 | 1.23 (0.83, 1.82) | 0.3039 | 1.27 (0.80, 2.02) | 0.3060 |

[[4]]

| | C72mean | AKI.Total | comparison | RR (95% CI) | p-value[1] | OR (95% CI) | p-value[1] |
|---|---|---|---|---|---|---|---|
| 1 | < 20 (n = 419) | 41 ( 9.79%) | > 25 vs. [20, 25] | 1.74 (1.26, 2.40) | **<0.001** | 2.09 (1.37, 3.19) | **<0.001** |
| 2 | [20, 25] (n = 221) | 41 (18.55%) | > 25 vs. < 20 | 3.30 (2.36, 4.62) | **<0.001** | 4.39 (2.92, 6.60) | **<0.001** |
| 3 | > 25 (n = 282) | 91 (32.27%) | [20, 25] vs. < 20 | 1.90 (1.27, 2.83) | **0.0018** | 2.10 (1.32, 3.35) | **0.0019** |

[[5]]

| | C72mean | AKI.EARLY | comparison | RR (95% CI) | p-value[1] | OR (95% CI) | p-value[1] |
|---|---|---|---|---|---|---|---|
| 1 | < 20 (n = 419) | 14 ( 3.34%) | > 25 vs. [20, 25] | 2.87 (1.56, 5.31) | **<0.001** | 3.22 (1.66, 6.26) | **<0.001** |
| 2 | [20, 25] (n = 221) | 12 ( 5.43%) | > 25 vs. < 20 | 4.67 (2.61, 8.36) | **<0.001** | 5.35 (2.87, 9.97) | **<0.001** |
| 3 | > 25 (n = 282) | 44 (15.60%) | [20, 25] vs. < 20 | 1.63 (0.76, 3.45) | 0.2067 | 1.66 (0.75, 3.66) | 0.2075 |

[[6]]

| | C72mean | AKI.LATE | comparison | RR (95% CI) | p-value[1] | OR (95% CI) | p-value[1] |
|---|---|---|---|---|---|---|---|
| 1 | < 20 (n = 419) | 27 ( 6.44%) | > 25 vs. [20, 25] | 1.27 (0.83, 1.95) | 0.2735 | 1.32 (0.80, 2.18) | 0.2716 |
| 2 | [20, 25] (n = 221) | 29 (13.12%) | > 25 vs. < 20 | 2.59 (1.65, 4.05) | **<0.001** | 2.90 (1.76, 4.79) | **<0.001** |
| 3 | > 25 (n = 282) | 47 (16.67%) | [20, 25] vs. < 20 | 2.04 (1.24, 3.35) | **0.0051** | 2.19 (1.26, 3.81) | **0.0053** |

[[7]]

| | C72mean | Clinical.failure.survival | comparison | RR (95% CI) | p-value[1] | OR (95% CI) | p-value[1] |
|---|---|---|---|---|---|---|---|
| 1 | < 20 (n = 419) | 75 (17.90%) | > 25 vs. [20, 25] | 0.86 (0.59, 1.25) | 0.4300 | 0.83 (0.52, 1.32) | 0.4306 |
| 2 | [20, 25] (n = 221) | 42 (19.00%) | > 25 vs. < 20 | 0.91 (0.65, 1.27) | 0.5864 | 0.89 (0.60, 1.34) | 0.5856 |
| 3 | > 25 (n = 282) | 46 (16.31%) | [20, 25] vs. < 20 | 1.06 (0.76, 1.49) | 0.7305 | 1.08 (0.71, 1.64) | 0.7310 |

[[8]]

| | C72mean | Microbiological.failure | comparison | RR (95% CI) | p-value[1] | OR (95% CI) | p-value[1] |
|---|---|---|---|---|---|---|---|
| 1 | < 20 (n = 419) | 143 (34.13%) | > 25 vs. [20, 25] | 1.00 (0.75, 1.33) | 0.9885 | 1.00 (0.68, 1.49) | 0.9885 |
| 2 | [20, 25] (n = 221) | 61 (27.60%) | > 25 vs. < 20 | 0.81 (0.64, 1.02) | 0.0744 | 0.74 (0.53, 1.03) | 0.0711 |
| 3 | > 25 (n = 282) | 78 (27.66%) | [20, 25] vs. < 20 | 0.81 (0.63, 1.04) | 0.0982 | 0.74 (0.51, 1.05) | 0.0926 |

**Subgroup A (n = 262)**

[[1]]

| | C72mean | ICU.MORTALITY | comparison | RR (95% CI) | p-value[1] | OR (95% CI) | p-value[1] |
|---|---|---|---|---|---|---|---|
| 1 | < 20 (n = 44) | 7 (15.91%) | > 25 vs. [20, 25] | 1.12 (0.71, 1.76) | 0.6269 | 1.18 (0.61, 2.27) | 0.6223 |
| 2 | [20, 25] (n = 56) | 17 (30.36%) | > 25 vs. < 20 | 2.13 (1.05, 4.35) | **0.0370** | 2.72 (1.14, 6.49) | **0.0245** |
| 3 | > 25 (n = 162) | 55 (33.95%) | [20, 25] vs. < 20 | 1.91 (0.87, 4.19) | 0.1074 | 2.30 (0.86, 6.19) | 0.0979 |

[[2]]

| | C72mean | HOSPITAL.MORTALITY | comparison | RR (95% CI) | p-value[1] | OR (95% CI) | p-value[1] |
|---|---|---|---|---|---|---|---|
| 1 | < 20 (n = 44) | 11 (25.00%) | > 25 vs. [20, 25] | 1.08 (0.77, 1.53) | 0.6609 | 1.15 (0.62, 2.12) | 0.6560 |
| 2 | [20, 25] (n = 56) | 24 (42.86%) | > 25 vs. < 20 | 1.85 (1.08, 3.17) | **0.0248** | 2.59 (1.22, 5.47) | **0.0129** |
| 3 | > 25 (n = 162) | 75 (46.30%) | [20, 25] vs. < 20 | 1.71 (0.95, 3.11) | 0.0756 | 2.25 (0.95, 5.34) | 0.0657 |

[[3]]

| | C72mean | 30D.MORTALITY | comparison | RR (95% CI) | p-value[1] | OR (95% CI) | p-value[1] |
|---|---|---|---|---|---|---|---|
| 1 | < 20 (n = 44) | 8 (18.18%) | > 25 vs. [20, 25] | 1.18 (0.75, 1.84) | 0.4695 | 1.28 (0.67, 2.46) | 0.4602 |
| 2 | [20, 25] (n = 56) | 17 (30.36%) | > 25 vs. < 20 | 1.97 (1.02, 3.81) | **0.0441** | 2.51 (1.09, 5.76) | **0.0299** |
| 3 | > 25 (n = 162) | 58 (35.80%) | [20, 25] vs. < 20 | 1.67 (0.80, 3.51) | 0.1756 | 1.96 (0.76, 5.10) | 0.1666 |

[[4]]

| | C72mean | AKI.Total | comparison | RR (95% CI) | p-value[1] | OR (95% CI) | p-value[1] |
|---|---|---|---|---|---|---|---|
| 1 | < 20 (n = 44) | 9 (20.45%) | > 25 vs. [20, 25] | 1.12 (0.70, 1.80) | 0.6279 | 1.18 (0.61, 2.30) | 0.6235 |
| 2 | [20, 25] (n = 56) | 16 (28.57%) | > 25 vs. < 20 | 1.57 (0.84, 2.93) | 0.1571 | 1.84 (0.82, 4.11) | 0.1374 |
| 3 | > 25 (n = 162) | 52 (32.10%) | [20, 25] vs. < 20 | 1.40 (0.68, 2.85) | 0.3595 | 1.56 (0.61, 3.96) | 0.3539 |

[[5]]

| | C72mean | AKI.EARLY | comparison | RR (95% CI) | p-value[1] | OR (95% CI) | p-value[1] |
|---|---|---|---|---|---|---|---|
| 1 | < 20 (n = 44) | 4 ( 9.09%) | > 25 vs. [20, 25] | 3.00 (0.94, 9.52) | 0.0628 | 3.38 (0.98,11.63) | 0.0537 |
| 2 | [20, 25] (n = 56) | 3 ( 5.36%) | > 25 vs. < 20 | 1.77 (0.65, 4.79) | 0.2646 | 1.91 (0.63, 5.80) | 0.2526 |
| 3 | > 25 (n = 162) | 26 (16.05%) | [20, 25] vs. < 20 | 0.59 (0.14, 2.50) | 0.4729 | 0.57 (0.12, 2.67) | 0.4724 |

[[6]]

| | C72mean | AKI.LATE | comparison | RR (95% CI) | p-value[1] | OR (95% CI) | p-value[1] |
|---|---|---|---|---|---|---|---|
| 1 | < 20 (n = 44) | 5 (11.36%) | > 25 vs. [20, 25] | 0.69 (0.38, 1.25) | 0.2220 | 0.63 (0.30, 1.34) | 0.2303 |
| 2 | [20, 25] (n = 56) | 13 (23.21%) | > 25 vs. < 20 | 1.41 (0.58, 3.46) | 0.4507 | 1.49 (0.54, 4.14) | 0.4431 |
| 3 | > 25 (n = 162) | 26 (16.05%) | [20, 25] vs. < 20 | 2.04 (0.79, 5.30) | 0.1417 | 2.36 (0.77, 7.22) | 0.1329 |

[[7]]

| | C72mean | Clinical.failure.survival | comparison | RR (95% CI) | p-value[1] | OR (95% CI) | p-value[1] |
|---|---|---|---|---|---|---|---|
| 1 | < 20 (n = 44) | 5 (11.36%) | > 25 vs. [20, 25] | 0.82 (0.38, 1.77) | 0.6148 | 0.80 (0.33, 1.94) | 0.6171 |
| 2 | [20, 25] (n = 56) | 8 (14.29%) | > 25 vs. < 20 | 1.03 (0.41, 2.61) | 0.9467 | 1.04 (0.36, 2.95) | 0.9467 |
| 3 | > 25 (n = 162) | 19 (11.73%) | [20, 25] vs. < 20 | 1.26 (0.44, 3.58) | 0.6679 | 1.30 (0.39, 4.29) | 0.6669 |

[[8]]

| | C72mean | Microbiological.failure | comparison | RR (95% CI) | p-value[1] | OR (95% CI) | p-value[1] |
|---|---|---|---|---|---|---|---|
| 1 | < 20 (n = 44) | 15 (34.09%) | > 25 vs. [20, 25] | 1.01 (0.61, 1.67) | 0.9567 | 1.02 (0.51, 2.02) | 0.9566 |
| 2 | [20, 25] (n = 56) | 15 (26.79%) | > 25 vs. < 20 | 0.80 (0.49, 1.29) | 0.3555 | 0.72 (0.35, 1.47) | 0.3684 |
| 3 | > 25 (n = 162) | 44 (27.16%) | [20, 25] vs. < 20 | 0.79 (0.43, 1.43) | 0.4284 | 0.71 (0.30, 1.67) | 0.4296 |

### Subgroup B (n = 186)

[[1]]

| | C72mean | ICU.MORTALITY | comparison | RR (95% CI) | p-value[1] | OR (95% CI) | p-value[1] |
|---|---|---|---|---|---|---|---|
| 1 | < 20 (n = 97) | 13 (13.40%) | > 25 vs. [20, 25] | 0.94 (0.41, 2.15) | 0.8772 | 0.92 (0.33, 2.61) | 0.8771 |
| 2 | [20, 25] (n = 48) | 10 (20.83%) | > 25 vs. < 20 | 1.46 (0.65, 3.25) | 0.3583 | 1.57 (0.59, 4.13) | 0.3637 |
| 3 | > 25 (n = 41) | 8 (19.51%) | [20, 25] vs. < 20 | 1.55 (0.74, 3.29) | 0.2479 | 1.70 (0.69, 4.22) | 0.2524 |

[[2]]

| | C72mean | HOSPITAL.MORTALITY | comparison | RR (95% CI) | p-value[1] | OR (95% CI) | p-value[1] |
|---|---|---|---|---|---|---|---|
| 1 | < 20 (n = 97) | 18 (18.56%) | > 25 vs. [20, 25] | 1.27 (0.65, 2.47) | 0.4835 | 1.39 (0.55, 3.52) | 0.4836 |
| 2 | [20, 25] (n = 48) | 12 (25.00%) | > 25 vs. < 20 | 1.71 (0.93, 3.15) | 0.0867 | 2.04 (0.89, 4.69) | 0.0942 |
| 3 | > 25 (n = 41) | 13 (31.71%) | [20, 25] vs. < 20 | 1.35 (0.71, 2.56) | 0.3639 | 1.46 (0.64, 3.35) | 0.3689 |

[[3]]

| | C72mean | 30D.MORTALITY | comparison | RR (95% CI) | p-value[1] | OR (95% CI) | p-value[1] |
|---|---|---|---|---|---|---|---|
| 1 | < 20 (n = 97) | 16 (16.49%) | > 25 vs. [20, 25] | 0.44 (0.12, 1.55) | 0.2003 | 0.39 (0.10, 1.60) | 0.1929 |
| 2 | [20, 25] (n = 48) | 8 (16.67%) | > 25 vs. < 20 | 0.44 (0.14, 1.44) | 0.1762 | 0.40 (0.11, 1.45) | 0.1641 |
| 3 | > 25 (n = 41) | 3 ( 7.32%) | [20, 25] vs. < 20 | 1.01 (0.47, 2.19) | 0.9791 | 1.01 (0.40, 2.56) | 0.9791 |

[[4]]

| | C72mean | AKI.Total | comparison | RR (95% CI) | p-value[1] | OR (95% CI) | p-value[1] |
|---|---|---|---|---|---|---|---|
| 1 | < 20 (n = 97) | 13 (13.40%) | > 25 vs. [20, 25] | 2.34 (1.05, 5.24) | **0.0385** | 3.04 (1.09, 8.50) | **0.0344** |
| 2 | [20, 25] (n = 48) | 7 (14.58%) | > 25 vs. < 20 | 2.55 (1.32, 4.93) | **0.0055** | 3.35 (1.40, 8.00) | **0.0065** |
| 3 | > 25 (n = 41) | 14 (34.15%) | [20, 25] vs. < 20 | 1.09 (0.46, 2.55) | 0.8458 | 1.10 (0.41, 2.97) | 0.8461 |

[[5]]

| | C72mean | AKI.EARLY | comparison | RR (95% CI) | p-value[1] | OR (95% CI) | p-value[1] |
|---|---|---|---|---|---|---|---|
| 1 | < 20 (n = 97) | 7 ( 7.22%) | > 25 vs. [20, 25] | 2.34 (0.62, 8.78) | 0.2071 | 2.57 (0.60,11.01) | 0.2032 |
| 2 | [20, 25] (n = 48) | 3 ( 6.25%) | > 25 vs. < 20 | 2.03 (0.73, 5.67) | 0.1775 | 2.20 (0.69, 7.02) | 0.1811 |
| 3 | > 25 (n = 41) | 6 (14.63%) | [20, 25] vs. < 20 | 0.87 (0.23, 3.20) | 0.8293 | 0.86 (0.21, 3.47) | 0.8290 |

[[6]]

| | C72mean | AKI.LATE | comparison | RR (95% CI) | p-value[1] | OR (95% CI) | p-value[1] |
|---|---|---|---|---|---|---|---|
| 1 | < 20 (n = 97) | 6 ( 6.19%) | > 25 vs. [20, 25] | 2.34 (0.76, 7.22) | 0.1385 | 2.67 (0.74, 9.61) | 0.1338 |
| 2 | [20, 25] (n = 48) | 4 ( 8.33%) | > 25 vs. < 20 | 3.15 (1.17, 8.52) | **0.0234** | 3.68 (1.19,11.39) | **0.0240** |
| 3 | > 25 (n = 41) | 8 (19.51%) | [20, 25] vs. < 20 | 1.35 (0.40, 4.55) | 0.6312 | 1.38 (0.37, 5.14) | 0.6322 |

[[7]]

| | C72mean | Clinical.failure.survival | comparison | RR (95% CI) | p-value[1] | OR (95% CI) | p-value[1] |
|---|---|---|---|---|---|---|---|
| 1 | < 20 (n = 97) | 8 ( 8.25%) | > 25 vs. [20, 25] | 2.34 (0.87, 6.30) | 0.0919 | 2.77 (0.86, 8.93) | 0.0870 |
| 2 | [20, 25] (n = 48) | 5 (10.42%) | > 25 vs. < 20 | 2.96 (1.26, 6.95) | **0.0129** | 3.59 (1.30, 9.91) | **0.0137** |
| 3 | > 25 (n = 41) | 10 (24.39%) | [20, 25] vs. < 20 | 1.26 (0.44, 3.65) | 0.6666 | 1.29 (0.40, 4.19) | 0.6677 |

[[8]]

| | C72mean | Microbiological.failure | comparison | RR (95% CI) | p-value[1] | OR (95% CI) | p-value[1] |
|---|---|---|---|---|---|---|---|
| 1 | < 20 (n = 97) | 31 (31.96%) | > 25 vs. [20, 25] | 0.72 (0.38, 1.33) | 0.2926 | 0.61 (0.25, 1.51) | 0.2861 |
| 2 | [20, 25] (n = 48) | 18 (37.50%) | > 25 vs. < 20 | 0.84 (0.47, 1.50) | 0.5564 | 0.78 (0.35, 1.76) | 0.5500 |

| | | | | | | | |
|---|---|---|---|---|---|---|---|
| 3 | > 25 (n = 41) | 11 (26.83%) | [20, 25] vs. < 20 | 1.17 (0.74, 1.87) | 0.5018 | 1.28 (0.62, 2.63) | 0.5072 |

### Subgroup C (n = 105)

[[1]]

| | C72mean | ICU.MORTALITY | comparison | RR (95% CI) | p-value[1] | OR (95% CI) | p-value[1] |
|---|---|---|---|---|---|---|---|
| 1 | < 20 (n = 33) | 3 ( 9.09%) | > 25 vs. [20, 25] | 4.79 (0.64,35.66) | 0.1264 | 5.68 (0.68,47.75) | 0.1095 |
| 2 | [20, 25] (n = 25) | 1 ( 4.00%) | > 25 vs. < 20 | 2.11 (0.62, 7.20) | 0.2346 | 2.37 (0.59, 9.52) | 0.2246 |
| 3 | > 25 (n = 47) | 9 (19.15%) | [20, 25] vs. < 20 | 0.44 (0.05, 3.98) | 0.4651 | 0.42 (0.04, 4.27) | 0.4607 |

[[2]]

| | C72mean | HOSPITAL.MORTALITY | comparison | RR (95% CI) | p-value[1] | OR (95% CI) | p-value[1] |
|---|---|---|---|---|---|---|---|
| 1 | < 20 (n = 33) | 4 (12.12%) | > 25 vs. [20, 25] | 2.48 (0.79, 7.83) | 0.1208 | 3.11 (0.80,12.10) | 0.1016 |
| 2 | [20, 25] (n = 25) | 3 (12.00%) | > 25 vs. < 20 | 2.46 (0.89, 6.80) | 0.0835 | 3.08 (0.91,10.40) | 0.0706 |
| 3 | > 25 (n = 47) | 14 (29.79%) | [20, 25] vs. < 20 | 0.99 (0.24, 4.03) | 0.9888 | 0.99 (0.20, 4.88) | 0.9888 |

[[3]]

| | C72mean | 30D.MORTALITY | comparison | RR (95% CI) | p-value[1] | OR (95% CI) | p-value[1] |
|---|---|---|---|---|---|---|---|
| 1 | < 20 (n = 33) | 3 ( 9.09%) | > 25 vs. [20, 25] | 6.38 (0.88,46.30) | 0.0667 | 8.23 (1.00,67.54) | **0.0497** |
| 2 | [20, 25] (n = 25) | 1 ( 4.00%) | > 25 vs. < 20 | 2.81 (0.86, 9.18) | 0.0874 | 3.43 (0.88,13.30) | 0.0749 |
| 3 | > 25 (n = 47) | 12 (25.53%) | [20, 25] vs. < 20 | 0.44 (0.05, 3.98) | 0.4651 | 0.42 (0.04, 4.27) | 0.4607 |

[[4]]

| | C72mean | AKI.Total | comparison | RR (95% CI) | p-value[1] | OR (95% CI) | p-value[1] |
|---|---|---|---|---|---|---|---|
| 1 | < 20 (n = 33) | 0 ( 0.00%) | > 25 vs. [20, 25] | 3.72 (0.92,15.10) | 0.0657 | 4.88 (1.01,23.55) | **0.0485** |
| 2 | [20, 25] (n = 25) | 2 ( 8.00%) | > 25 vs. < 20 | -[2] (-[2], -[2]) | -[2] | -[2] (-[2], -[2]) | -[2] |
| 3 | > 25 (n = 47) | 14 (29.79%) | [20, 25] vs. < 20 | -[2] (-[2], -[2]) | -[2] | -[2] (-[2], -[2]) | -[2] |

[[5]]

| | C72mean | AKI.EARLY | comparison | RR (95% CI) | p-value[1] | OR (95% CI) | p-value[1] |
|---|---|---|---|---|---|---|---|
| 1 | < 20 (n = 33) | 0 ( 0.00%) | > 25 vs. [20, 25] | -[2] (-[2], -[2]) | -[2] | -[2] (-[2], -[2]) | -[2] |
| 2 | [20, 25] (n = 25) | 0 ( 0.00%) | > 25 vs. < 20 | -[2] (-[2], -[2]) | -[2] | -[2] (-[2], -[2]) | -[2] |
| 3 | > 25 (n = 47) | 5 (10.64%) | [20, 25] vs. < 20 | -[2] (-[2], -[2]) | -[2] | -[2] (-[2], -[2]) | -[2] |

[[6]]

| | C72mean | AKI.LATE | comparison | RR (95% CI) | p-value[1] | OR (95% CI) | p-value[1] |
|---|---|---|---|---|---|---|---|
| 1 | < 20 (n = 33) | 0 ( 0.00%) | > 25 vs. [20, 25] | 2.39 (0.56,10.24) | 0.2392 | 2.72 (0.54,13.73) | 0.2246 |
| 2 | [20, 25] (n = 25) | 2 ( 8.00%) | > 25 vs. < 20 | -[2] (-[2], -[2]) | -[2] | -[2] (-[2], -[2]) | -[2] |
| 3 | > 25 (n = 47) | 9 (19.15%) | [20, 25] vs. < 20 | -[2] (-[2], -[2]) | -[2] | -[2] (-[2], -[2]) | -[2] |

[[7]]

| | C72mean | Clinical.failure.survival | comparison | RR (95% CI) | p-value[1] | OR (95% CI) | p-value[1] |
|---|---|---|---|---|---|---|---|
| 1 | < 20 (n = 33) | 8 (24.24%) | > 25 vs. [20, 25] | 0.89 (0.36, 2.16) | 0.7904 | 0.86 (0.27, 2.71) | 0.7914 |
| 2 | [20, 25] (n = 25) | 6 (24.00%) | > 25 vs. < 20 | 0.88 (0.39, 1.99) | 0.7540 | 0.84 (0.29, 2.44) | 0.7546 |
| 3 | > 25 (n = 47) | 10 (21.28%) | [20, 25] vs. < 20 | 0.99 (0.39, 2.49) | 0.9830 | 0.99 (0.29, 3.33) | 0.9830 |

[[8]]

| | C72mean | Microbiological.failure | comparison | RR (95% CI) | p-value[1] | OR (95% CI) | p-value[1] |
|---|---|---|---|---|---|---|---|
| 1 | < 20 (n = 33) | 11 (33.33%) | > 25 vs. [20, 25] | 0.99 (0.45, 2.16) | 0.9755 | 0.98 (0.33, 2.90) | 0.9755 |
| 2 | [20, 25] (n = 25) | 7 (28.00%) | > 25 vs. < 20 | 0.83 (0.43, 1.62) | 0.5842 | 0.76 (0.29, 2.01) | 0.5861 |
| 3 | > 25 (n = 47) | 13 (27.66%) | [20, 25] vs. < 20 | 0.84 (0.38, 1.86) | 0.6663 | 0.78 (0.25, 2.42) | 0.6640 |

### Subgroup D (n = 369)

[[1]]

| | C72mean | ICU.MORTALITY | comparison | RR (95% CI) | p-value[1] | OR (95% CI) | p-value[1] |
|---|---|---|---|---|---|---|---|
| 1 | < 20 (n = 245) | 21 ( 8.57%) | > 25 vs. [20, 25] | 2.46 (0.89, 6.79) | 0.0811 | 2.80 (0.86, 9.08) | 0.0858 |
| 2 | [20, 25] (n = 92) | 7 ( 7.61%) | > 25 vs. < 20 | 2.19 (0.95, 5.01) | 0.0643 | 2.46 (0.91, 6.65) | 0.0757 |
| 3 | > 25 (n = 32) | 6 (18.75%) | [20, 25] vs. < 20 | 0.89 (0.39, 2.02) | 0.7761 | 0.88 (0.36, 2.14) | 0.7756 |

[[2]]

| | C72mean | HOSPITAL.MORTALITY | comparison | RR (95% CI) | p-value[1] | OR (95% CI) | p-value[1] |
|---|---|---|---|---|---|---|---|
| 1 | < 20 (n = 245) | 31 (12.65%) | > 25 vs. [20, 25] | 1.55 (0.68, 3.54) | 0.2998 | 1.70 (0.61, 4.73) | 0.3085 |
| 2 | [20, 25] (n = 92) | 13 (14.13%) | > 25 vs. < 20 | 1.73 (0.83, 3.60) | 0.1431 | 1.93 (0.77, 4.84) | 0.1598 |
| 3 | > 25 (n = 32) | 7 (21.88%) | [20, 25] vs. < 20 | 1.12 (0.61, 2.04) | 0.7190 | 1.14 (0.57, 2.28) | 0.7200 |

[[3]]

| | C72mean | 30D.MORTALITY | comparison | RR (95% CI) | p-value[1] | OR (95% CI) | p-value[1] |
|---|---|---|---|---|---|---|---|
| 1 | < 20 (n = 245) | 27 (11.02%) | > 25 vs. [20, 25] | 1.28 (0.42, 3.87) | 0.6643 | 1.32 (0.38, 4.61) | 0.6663 |
| 2 | [20, 25] (n = 92) | 9 ( 9.78%) | > 25 vs. < 20 | 1.13 (0.42, 3.03) | 0.8017 | 1.15 (0.38, 3.54) | 0.8030 |
| 3 | > 25 (n = 32) | 4 (12.50%) | [20, 25] vs. < 20 | 0.89 (0.43, 1.82) | 0.7441 | 0.88 (0.40, 1.94) | 0.7433 |

[[4]]

| | C72mean | AKI.Total | comparison | RR (95% CI) | p-value[1] | OR (95% CI) | p-value[1] |
|---|---|---|---|---|---|---|---|
| 1 | < 20 (n = 245) | 19 ( 7.76%) | > 25 vs. [20, 25] | 1.98 (1.03, 3.80) | **0.0411** | 2.49 (1.00, 6.16) | **0.0489** |
| 2 | [20, 25] (n = 92) | 16 (17.39%) | > 25 vs. < 20 | 4.43 (2.33, 8.45) | **<0.001** | 6.23 (2.62,14.82) | **<0.001** |
| 3 | > 25 (n = 32) | 11 (34.38%) | [20, 25] vs. < 20 | 2.24 (1.21, 4.17) | **0.0107** | 2.50 (1.23, 5.11) | **0.0117** |

[[5]]

| | C72mean | AKI.EARLY | comparison | RR (95% CI) | p-value[1] | OR (95% CI) | p-value[1] |
|---|---|---|---|---|---|---|---|
| 1 | < 20 (n = 245) | 3 ( 1.22%) | > 25 vs. [20, 25] | 3.35 (1.22, 9.24) | **0.0193** | 4.01 (1.24,13.03) | **0.0208** |
| 2 | [20, 25] (n = 92) | 6 ( 6.52%) | > 25 vs. < 20 | 17.86 (4.86,65.64) | **<0.001** | 22.59 (5.49,92.87) | **<0.001** |
| 3 | > 25 (n = 32) | 7 (21.88%) | [20, 25] vs. < 20 | 5.33 (1.36,20.86) | **0.0163** | 5.63 (1.38,23.00) | **0.0161** |

[[6]]

| | C72mean | AKI.LATE | comparison | RR (95% CI) | p-value[1] | OR (95% CI) | p-value[1] |
|---|---|---|---|---|---|---|---|
| 1 | < 20 (n = 245) | 16 ( 6.53%) | > 25 vs. [20, 25] | 1.15 (0.39, 3.41) | 0.8011 | 1.17 (0.34, 4.03) | 0.8019 |
| 2 | [20, 25] (n = 92) | 10 (10.87%) | > 25 vs. < 20 | 1.91 (0.68, 5.37) | 0.2175 | 2.04 (0.64, 6.55) | 0.2284 |
| 3 | > 25 (n = 32) | 4 (12.50%) | [20, 25] vs. < 20 | 1.66 (0.78, 3.53) | 0.1847 | 1.75 (0.76, 4.00) | 0.1881 |

[[7]]

| | C72mean | Clinical.failure.survival | comparison | RR (95% CI) | p-value[1] | OR (95% CI) | p-value[1] |
|---|---|---|---|---|---|---|---|
| 1 | < 20 (n = 245) | 54 (22.04%) | > 25 vs. [20, 25] | 0.88 (0.42, 1.84) | 0.7251 | 0.84 (0.32, 2.20) | 0.7224 |
| 2 | [20, 25] (n = 92) | 23 (25.00%) | > 25 vs. < 20 | 0.99 (0.49, 1.99) | 0.9830 | 0.99 (0.41, 2.41) | 0.9830 |
| 3 | > 25 (n = 32) | 7 (21.88%) | [20, 25] vs. < 20 | 1.13 (0.74, 1.74) | 0.5614 | 1.18 (0.67, 2.06) | 0.5646 |

[[8]]

| | C72mean | Microbiological.failure | comparison | RR (95% CI) | p-value[1] | OR (95% CI) | p-value[1] |
|---|---|---|---|---|---|---|---|
| 1 | < 20 (n = 245) | 86 (35.10%) | > 25 vs. [20, 25] | 1.37 (0.72, 2.59) | 0.3335 | 1.54 (0.63, 3.75) | 0.3451 |
| 2 | [20, 25] (n = 92) | 21 (22.83%) | > 25 vs. < 20 | 0.89 (0.52, 1.53) | 0.6739 | 0.84 (0.38, 1.86) | 0.6670 |
| 3 | > 25 (n = 32) | 10 (31.25%) | [20, 25] vs. < 20 | 0.65 (0.43, 0.98) | **0.0409** | 0.55 (0.31, 0.95) | **0.0324** |

Overall (n=922); subgroup A (n=262): eGFR at sCIV <90 mL/min/1.73 m² and SAPS II at sCIV >36; subgroup B (n=186): eGFR at sCIV ≥90 mL/min/1.73 m² and SAPS II at sCIV >36; subgroup C (n=105): eGFR at sCIV <90 mL/min/1.73 m² and SAPS II at sCIV ≤36; subgroup D (n=369): eGFR at sCIV ≥90 mL/min/1.73 m² and SAPS II at sCIV ≤36.

Abbreviations: AKI, acute kidney injury; AKI.Early, early acute kidney injury; AKI.Late, late acute kidney injury; AKI.Total, total acute kidney injury; C72mean, mean vancomycin serum concentration during the first three days of continuous infusion of vancomycin [mg/L]; CI, confidence interval; $eGFR_{sCIV}$, estimated glomerular filtration rate at the start of continuous infusion of vancomycin [mL/min/1.73 m²]; ICU, intensive care unit; OR, odds ratio; RR, relative risk; $SAPS_{sCIV}$, Simplified Acute Physiology Score II at the start of continuous infusion of vancomycin.

[1] p-value <0.05 was considered significant.

[2] Value could not be calculated due to missing AKI events

### 1.9 Regression Tables

**Table S 11** Predictors for mortality (intensive care unit (ICU), hospital, 30-day) during continuous infusion of vancomycin determined by multivariate logistic and Cox regression analysis (n=922, no missing values).

| | ICU mortality | | | | | | In-hospital mortality | | | | | | 30-day mortality | | | | | |
|---|---|---|---|---|---|---|---|---|---|---|---|---|---|---|---|---|---|---|
| | Logistic regression | | | Cox regression | | | Logistic regression | | | Cox regression | | | Logistic regression | | | Cox regression | | |
| **Predictors** | **OR** | **95% CI** | **p-value[1]** | **HR** | **95% CI** | **p-value[1]** | **OR** | **95% CI** | **p-value[1]** | **HR** | **95% CI** | **p-value[1]** | **OR** | **95% CI** | **p-value[1]** | **HR** | **95% CI** | **p-value[1]** |
| C72mean = <20 mg/L | 0.545 | 0.232-1.285 | 0.166 | 0.962 | 0.461-2.006 | 0.918 | 0.463 | 0.218-0.981 | **0.045** | 0.789 | 0.431-1.447 | 0.444 | 0.593 | 0.263-1.339 | 0.208 | 0.790 | 0.395-1.582 | 0.506 |
| C72mean = 20-25 mg/L | 0.937 | 0.459-1.912 | 0.859 | 1.222 | 0.694-2.123 | 0.487 | 0.777 | 0.406-1.490 | 0.448 | 0.993 | 0.624-1.581 | 0.977 | 0.738 | 0.369-1.476 | 0.390 | 0.895 | 0.515-1.557 | 0.696 |
| eGFR$_{sCIV}$ = ≥90 mL/min/1.73 m² | 0.433 | 0.190-0.989 | **0.047** | 0.558 | 0.282-1.103 | 0.094 | 0.387 | 0.185-0.810 | **0.012** | 0.517 | 0.297-0.902 | **0.020** | 0.114 | 0.040-0.319 | **<0.001** | 0.178 | 0.071-0.451 | **<0.001** |
| Gender = female | 1.316 | 0.855-2.024 | 0.212 | 1.288 | 0.906-1.829 | 0.158 | 0.945 | 0.646-1.382 | 0.771 | 1.048 | 0.783-1.405 | 0.751 | 0.934 | 0.619-1.408 | 0.743 | 0.984 | 0.701-1.380 | 0.924 |
| BMI [kg/m²] | 0.961 | 0.920-1.004 | 0.073 | 0.961 | 0.927-0.996 | **0.030** | 0.941 | 0.903-0.981 | **0.004** | 0.960 | 0.931-0.989 | **0.008** | 0.966 | 0.925-1.008 | 0.106 | 0.974 | 0.941-1.009 | 0.144 |
| SAPS$_{sCIV}$ = ≤36 | 0.578 | 0.261-1.277 | 0.175 | 0.783 | 0.411-1.494 | 0.458 | 0.519 | 0.256-1.052 | 0.069 | 0.157 | 0.446-1.284 | 0.302 | 0.751 | 0.357-1.579 | 0.450 | 0.805 | 0.446-1.453 | 0.472 |
| Cardiovascular comorbidity = yes | 2.210 | 1.266-3.858 | **0.005** | 1.913 | 1.177-3.109 | **0.009** | 2.045 | 1.281-3.266 | **0.003** | 1.868 | 1.263-2.763 | **0.002** | 1.568 | 0.948-2.592 | 0.080 | 1.501 | 0.997-2.332 | 0.070 |
| Sepsis = yes | 0.872 | 0.540-1.410 | 0.577 | 0.840 | 0.567-1.246 | 0.387 | 0.933 | 0.610-1.427 | 0.750 | 0.853 | 0.615-1.181 | 0.338 | 0.979 | 0.617-1.553 | 0.927 | 0.939 | 0.641-1.376 | 0.748 |
| Septic shock = yes | 1.265 | 0.701-2.284 | 0.435 | 1.085 | 0.683-1.724 | 0.729 | 1.494 | 0.866-2.577 | 0.149 | 1.166 | 0.788-1.726 | 0.441 | 1.598 | 0.900-2.837 | 0.110 | 1.331 | 0.852-2.080 | 0.210 |
| No germ detected = yes | 1.919 | 0.127-3.026 | **0.005** | 1.565 | 1.072-2.284 | **0.020** | 1.758 | 1.176-2.629 | **0.006** | 1.299 | 0.955-1.768 | 0.096 | 1.637 | 1.070-2.505 | **0.023** | 1.391 | 0.984-1.966 | 0.062 |
| No leukocyte reduction (day 1-7) = yes | 1.771 | 1.187-2.641 | **0.005** | 1.350 | 0.971-1.876 | 0.074 | 1.696 | 1.187-2.424 | **0.004** | 1.362 | 1.035-1.792 | **0.027** | 1.458 | 0.991-2.144 | 0.055 | 1.363 | 0.990-1.876 | 0.058 |
| No CRP reduction (day 1-7) = yes | 1.363 | 0.855-2.174 | 0.193 | 1.334 | 0.917-1.940 | 0.132 | 1.177 | 0.775-1.788 | 0.445 | 1.183 | 0.861-1.627 | 0.300 | 1.498 | 0.968-2.318 | 0.070 | 1.515 | 1.062-2.162 | **0.022** |
| Total AKI = yes | 4.389 | 2.836-6.792 | **<0.001** | 3.085 | 2.170-4.386 | **<0.001** | 4.213 | 2.792-6.358 | **<0.001** | 2.699 | 2.009-3.627 | **<0.001** | 3.729 | 2.424-5.737 | **<0.001** | 2.748 | 1.951-3.869 | **<0.001** |
| Vasopressors = yes | 4.533 | 2.203-9.326 | **<0.001** | 2.420 | 1.263-4.636 | **0.008** | 3.416 | 1.971-5.922 | **<0.001** | 2.325 | 1.450-3.729 | **<0.001** | 2.717 | 1.506-4.904 | **<0.001** | 2.282 | 1.356-3.840 | **0.002** |
| Mechanical ventilation = yes | 1.258 | 0.709-2.231 | 0.432 | 0.821 | 0.504-1.338 | 0.429 | 0.813 | 0.507-1.304 | 0.390 | 0.860 | 0.593-1.249 | 0.429 | 0.860 | 0.515-1.437 | 0.565 | 0.876 | 0.568-1.349 | 0.547 |
| Vancomycin dose day 1 [mg/kg TBW] | 0.988 | 0.962-1.015 | 0.373 | 0.992 | 0.972-1.012 | 0.416 | 0.994 | 0.972-1.018 | 0.628 | 0.998 | 0.982-1.015 | 0.845 | 1.001 | 0.977-1.026 | 0.920 | 1.001 | 0.982-1.021 | 0.911 |
| Length of stay in the ICU before IOT [days] | 0.993 | 0.971-1.016 | 0.563 | 0.989 | 0.969-1.009 | 0.282 | 0.987 | 0.966-1.007 | 0.199 | 0.992 | 0.976-1.009 | 0.369 | 0.995 | 0.974-1.017 | 0.677 | 0.998 | 0.980-1.017 | 0.837 |
| Microbiological failure = yes | 1.251 | 0.787-1.989 | 0.344 | 0.812 | 0.548-1.203 | 0.299 | 1.127 | 0.747-1.701 | 0.569 | 0.803 | 0.580-1.112 | 0.186 | 0.729 | 0.461-1.154 | 0.178 | 0.682 | 0.463-1.005 | 0.053 |
| Loading dose according to protocol = yes | 2.092 | 0.820-5.336 | 0.122 | 1.989 | 0.950-4.166 | 0.068 | 1.690 | 0.738-3.872 | 0.215 | 1.497 | 0.806-2.781 | 0.202 | 1.225 | 0.491-3.057 | 0.664 | 1.243 | 0.586-2.638 | 0.571 |
| Initial maintenance dose according to protocol = yes | 0.766 | 0.494-1.189 | 0.235 | 0.730 | 0.506-1.051 | 0.091 | 1.042 | 0.715-1.518 | 0.831 | 1.010 | 0.755-1.351 | 0.949 | 0.997 | 0.663-1.500 | 0.990 | 0.936 | 0.666-1.314 | 0.701 |
| C72mean = <20 mg/L:SAPS$_{sCIV}$ = ≤36 | 0,742 | 0,231-2,389 | 0,617 | 0.669 | 0.239-1.872 | 0.444 | 0,899 | 0,329-2,451 | 0,834 | 0.657 | 0.286-1.509 | 0.322 | 0,423 | 0,125-1,428 | 0,166 | 0.425 | 0.143-1.263 | 0.124 |
| C72mean = 20-25 mg/L:SAPS$_{sCIV}$ = ≤36 | 0,251 | 0,067-0,941 | **0,040** | 0.257 | 0.078-0.843 | **0.025** | 0,530 | 0,180-1,559 | 0,249 | 0.408 | 0.167-1.001 | 0.050 | 0,254 | 0,067-0,967 | **0,045** | 0.265 | 0.079-0.885 | **0.031** |
| C72mean = <20 mg/L:eGFR$_{sCIV}$ = ≥90 mL/min/1.73 m² | 1,697 | 0,545-5,279 | 0,361 | 1.211 | 0.459-3.200 | 0.699 | 1,808 | 0,674-4,846 | 0,240 | 1.455 | 0.659-3.212 | 0.353 | 6,620 | 1,938-22,611 | **0,003** | 4.253 | 1.429-12.659 | **0.009** |
| C72mean = 20-25 mg/L:eGFR$_{sCIV}$ = ≥90 mL/min/1.73 m² | 1,971 | 0,608-6,389 | 0,258 | 1.628 | 0.601-4.411 | 0.338 | 1,712 | 0,615-4,764 | 0,304 | 1.600 | 0.714-3.584 | 0.254 | 5,821 | 1,583-21,403 | **0,008** | 3.844 | 1.215-12.164 | **0.022** |
| eGFR$_{sCIV}$ = ≥90 mL/min/1.73 m²:SAPS$_{sCIV}$ = ≤36 | 1,985 | 0,679-5,800 | 0,210 | 1.534 | 0.589-3.996 | 0.381 | 1,652 | 0,680-4,017 | 0,268 | 1.414 | 0.674-2.967 | 0.360 | 2,552 | 0,843-7,727 | 0,097 | 2.268 | 0.829-6.209 | 0.111 |

Abbreviations: AKI, acute kidney injury; BMI, body mass index; C72mean, mean vancomycin serum concentration during the first three days of continuous infusion of vancomycin; CI, confidence interval; CRP, C-reactive protein; eGFR$_{sCIV}$, estimated glomerular filtration rate at the start of continuous infusion of vancomycin; HR, hazard ratio; ICU, intensive care unit; IOT, initiation of vancomycin therapy; OR, odds ratio; SAPS$_{sCIV}$, Simplified Acute Physiology Score II at the start of continuous infusion of vancomycin; TBW, total body weight.

Reference values: C72mean, >25 mg/L; eGFR$_{sCIV}$, <90 mL/min/1.73 m²; SAPS$_{sCIV}$, >36

[1]p-value <0.05 was considered significant.

**Table S 12** Predictors for the development of acute kidney injury (AKI, total AKI, early AKI, late AKI) during continuous infusion of vancomycin determined by multivariate logistic and Cox regression analysis (n=922, no missing values).

| | Total AKI | | | | | | Early AKI | | | | | | Late AKI | | | | | |
|---|---|---|---|---|---|---|---|---|---|---|---|---|---|---|---|---|---|---|
| | Logistic regression | | | Cox regression | | | Logistic regression | | | Cox regression | | | Logistic regression | | | Cox regression | | |
| **Predictors** | **OR** | **95% CI** | **p-value[1]** | **HR** | **95% CI** | **p-value[1]** | **OR** | **95% CI** | **p-value[1]** | **HR** | **95% CI** | **p-value[1]** | **OR** | **95% CI** | **p-value[1]** | **HR** | **95% CI** | **p-value[1]** |
| C72mean = <20 mg/L | 0,352 | 0,150-0,829 | **0,017** | 0,421 | 0,202-0,877 | **0,021** | 0,411 | 0,125-1,354 | 0,144 | 0,451 | 0,151-1,352 | 0,155 | 0,392 | 0,132-1,161 | 0,091 | 0,379 | 0,137-1,048 | 0,061 |
| C72mean = 20-25 mg/L | 0,611 | 0,310-1,203 | 0,154 | 0,618 | 0,353-1,082 | 0,092 | 0,200 | 0,055-0,722 | **0,014** | 0,233 | 0,068-0,792 | **0,020** | 1,178 | 0,549-2,528 | 0,673 | 0,939 | 0,481-1,834 | 0,854 |
| eGFR$_{sCIV}$ = ≥90 mL/min/1.73 m² | 0,903 | 0,426-1,914 | 0,790 | 0,883 | 0,480-1,623 | 0,689 | 1,020 | 0,361-2,880 | 0,970 | 0,948 | 0,374-2,400 | 0,910 | 0,763 | 0,298-1,952 | 0,573 | 0,771 | 0,333-1,787 | 0,544 |
| Gender = female | 0,878 | 0,591-1,302 | 0,516 | 0,918 | 0,656-1,283 | 0,616 | 0,908 | 0,515-1,600 | 0,738 | 0,921 | 0,550-1,542 | 0,755 | 0,846 | 0,524-1,368 | 0,495 | 0,893 | 0,572-1,394 | 0,619 |
| BMI [kg/m²] | 1,023 | 0,987-1,061 | 0,210 | 1,013 | 0,984-1,042 | 0,391 | 0,973 | 0,924-1,025 | 0,304 | 0,976 | 0,933-1,022 | 0,304 | 1,048 | 1,005-1,093 | **0,029** | 1,041 | 0,1003-1,080 | **0,036** |
| SAPS$_{sCIV}$ = ≤36 | 0,720 | 0,351-1,476 | 0,370 | 0,788 | 0,438-1,418 | 0,427 | 0,572 | 0,197-1,665 | 0,305 | 0,615 | 0,232-1,626 | 0,327 | 0,805 | 0,339-1,914 | 0,624 | 0,841 | 0,388-1,825 | 0,662 |
| Duration of vancomycin therapy [days] | 1,045 | 1,007-1,084 | **0,019** | 0,995 | 0,961-1,031 | 0,7782 | 0,970 | 0,907-1,038 | 0,380 | 0,976 | 0,917-1,039 | 0,445 | 1,072 | 1,030-1,116 | **<0,001** | 1,007 | 0,964-1,051 | 0,758 |
| Sepsis = yes | 1,467 | 0,952-2,260 | 0,082 | 1,411 | 0,981-2,028 | 0,063 | 2,390 | 1,318-4,336 | **0,04** | 2,237 | 1,302-3,843 | **0,004** | 0,902 | 0,516-1,576 | 0,717 | 0,935 | 0,559-1,566 | 0,799 |
| Septic shock = yes | 2,594 | 1,504-4,472 | **<0,001** | 2,143 | 1,384-3,317 | **<0,001** | 2,434 | 1,105-5,363 | **0,027** | 2,240 | 1,095-4,584 | **0,027** | 2,173 | 1,165-4,054 | **0,015** | 2,203 | 1,262-3,848 | **0,006** |
| Aminoglycosides = yes | 4,875 | 1,823-13,035 | **0,002** | 3,155 | 1,554-6,406 | **0,002** | 1,917 | 0,378-9,721 | 0,432 | 2,027 | 0,469-8,767 | 0,345 | 5,092 | 1,847-14,041 | **0,002** | 3,993 | 1,758-9,071 | **<0,001** |
| Loop diuretics= yes | 2,156 | 1,266-3,670 | **0,005** | 1,937 | 1,205-3,115 | **0,006** | 2,892 | 1,188-7,043 | **0,019** | 2,587 | 1,122-5,963 | **0,026** | 1,655 | 0,890-3,077 | 0,112 | 1,702 | 0,954-3,039 | 0,072 |
| Piperacillin/Tazobactam = yes | 2,450 | 0,876-6,848 | 0,088 | 1,840 | 0,831-4,073 | 0,133 | 2,339 | 0,580-9,437 | 0,233 | 2,040 | 0,611-6,813 | 0,247 | 1,910 | 0,587-6,213 | 0,282 | 1,872 | 0,653-5,365 | 0,243 |
| ACEI or ARB = yes | 0,871 | 0,538-1,411 | 0,575 | 0,873 | 0,586-1,302 | 0,506 | 0,613 | 0,285-1,321 | 0,212 | 0,675 | 0,339-1,347 | 0,265 | 1,064 | 0,608-1,862 | 0,828 | 1,006 | 0,611-1,656 | 0,982 |
| Vasopressors = yes | 1,024 | 0,615-1,705 | 0,928 | 0,967 | 0,623-1,501 | 0,881 | 1,205 | 0,543-2,670 | 0,647 | 1,159 | 0,556-2,415 | 0,694 | 0,955 | 0,524-1,742 | 0,882 | 0,869 | 0,499-1,513 | 0,619 |
| CKD = yes | 0,586 | 0,327-1,049 | 0,072 | 0,622 | 0,381-1,014 | 0,057 | 0,663 | 0,290-1,517 | 0,330 | 0,686 | 0,322-1,465 | 0,331 | 0,650 | 0,319-1,323 | 0,235 | 0,619 | 0,326-1,177 | 0,144 |
| Diabetes mellitus = yes | 0,844 | 0,537-1,327 | 0,463 | 0,837 | 0,577-1,215 | 0,349 | 0,623 | 0,314-1,237 | 0,176 | 0,660 | 0,354-1,233 | 0,193 | 1,060 | 0,623-1,804 | 0,829 | 1,008 | 0,627-1,618 | 0,975 |
| Hypertension = yes | 1,155 | 0,763-1,750 | 0,496 | 1,116 | 0,782-1,592 | 0,546 | 1,309 | 0,705-2,433 | 0,394 | 1,255 | 0,713-2,208 | 0,431 | 1,019 | 0,619-1,678 | 0,942 | 1,018 | 0,643-1,613 | 0,938 |
| Congestive heart failure = yes | 0,820 | 0,475-1,415 | 0,475 | 0,767 | 0,485-1,213 | 0,257 | 0,554 | 0,237-1,294 | 0,172 | 0,574 | 0,264-1,251 | 0,163 | 1,032 | 0,548-1,943 | 0,923 | 0,876 | 0,494-1,554 | 0,650 |
| Bacteraemia = yes | 0,846 | 0,543-1,318 | 0,460 | 0,877 | 0,601-1,279 | 0,495 | 0,903 | 0,480-1,698 | 0,751 | 0,940 | 0,531-1,662 | 0,831 | 0,817 | 0,473-1,413 | 0,470 | 0,831 | 0,503-1,375 | 0,472 |
| Vancomycin dose day 1-3 [mg/kg TBW/day] | 0,967 | 0,939-0,996 | **0,025** | 0,967 | 0,942-0,992 | **0,009** | 0,931 | 0,889-0,974 | **0,002** | 0,936 | 0,898-0,976 | **0,002** | 0,996 | 0,962-1,031 | 0,809 | 0,989 | 0,957-1,022 | 0,494 |
| Length of stay in the ICU [days] | 0,999 | 0,990-1,008 | 0,805 | 0,999 | 0,992-1,007 | 0,838 | 1,004 | 0,993-1,015 | 0,531 | 1,004 | 0,994-1,013 | 0,457 | 0,995 | 0,983-1,007 | 0,373 | 0,995 | 0,984-1,006 | 0,332 |
| C72mean = <20 mg/L:SAPS$_{sCIV}$ = ≤36 | 0,230 | 0,079-0,671 | **0,007** | 0,235 | 0,094-0,590 | **0,002** | 0,055 | 0,09-0,344 | **0,002** | 0,055 | 0,010-0,304 | **<0,001** | 0,517 | 0,144-1,858 | 0,312 | 0,447 | 0,138-1,454 | 0,181 |
| C72mean = 20-25 mg/L:SAPS$_{sCIV}$ = ≤36 | 0,515 | 0,175-1,522 | 0,230 | 0,519 | 0,204-1,322 | 0,169 | 0,380 | 0,060-2,388 | 0,302 | 0,369 | 0,068-2,015 | 0,250 | 0,543 | 0,155-1,897 | 0,339 | 0,497 | 0,159-1,556 | 0,230 |
| C72mean = <20 mg/L:eGFR$_{sCIV}$ = ≥90 mL/min/1.73 m² | 1,212 | 0,403-3,644 | 0,733 | 1,225 | 0,474-3,169 | 0,675 | 1,387 | 0,270-7,110 | 0,695 | 1,525 | 0,337-6,897 | 0,583 | 1,297 | 0,337-4,996 | 0,706 | 1,263 | 0,362-4,410 | 0,714 |
| C72mean = 20-25 mg/L:eGFR$_{sCIV}$ = ≥90 mL/min/1.73 m² | 0,781 | 0,275-2,220 | 0,643 | 0,895 | 0,365-2,191 | 0,808 | 2,488 | 0,386-16,025 | 0,338 | 2,421 | 0,420-13,958 | 0,323 | 0,630 | 0,185-2,138 | 0,458 | 0,745 | 0,245-2,264 | 0,603 |
| eGFR$_{sCIV}$ = ≥90 mL/min/1.73 m²:SAPS$_{sCIV}$ = ≤36 | 3,186 | 1,229-8,259 | **0,017** | 2,788 | 1,239-6,277 | **0,013** | 6,025 | 1,230-29,518 | **0,027** | 5,599 | 1,347-23,279 | **0,018** | 2,050 | 0,678-6,198 | 0,204 | 2,085 | 0,752-5,781 | 0,158 |

Abbreviations: ACEI, angiotensin-converting enzyme inhibitor; AKI, acute kidney injury; ARB, angiotensin receptor blocker; BMI, body mass index; C72mean, mean vancomycin serum concentration during the first three days of continuous infusion of vancomycin; CI, confidence interval; CKD, chronic kindey disease; eGFR$_{sCIV}$, estimated glomerular filtration rate at the start of continuous infusion of vancomycin; HR, hazard ratio; ICU, intensive care unit; OR, odds ratio; SAPS$_{sCIV}$, Simplified Acute Physiology Score II at the start of continuous infusion of vancomycin; TBW, total body weight.

Reference values: C72mean, >25 mg/L; eGFR$_{sCIV}$, <90 mL/min/1.73 m²; SAPS$_{sCIV}$, >36.

p-value <0.05 was considered significant.

**Table S 13** Estimated model coefficients of the logistic regression for clinical failure with survival. (n=922, no missing values).

<table>
<tr><th></th><th>Estimate</th><th>Std. Error</th><th>z value</th><th>Pr(>|z|)[1]</th><th>Odds Ratio</th><th>2.5%</th><th>97.5%</th></tr>
<tr><td>(Intercept)</td><td>-1.2996</td><td>0.8915</td><td>-1.4579</td><td>0.1449</td><td>0.2726</td><td>0.0475</td><td>1.5646</td></tr>
<tr><td>C72mean = < 20 mg/L</td><td>-0.4339</td><td>0.4741</td><td>-0.9151</td><td>0.3602</td><td>0.6480</td><td>0.2559</td><td>1.6412</td></tr>
<tr><td>C72mean = 20-25 mg/L</td><td>-0.2782</td><td>0.4299</td><td>-0.6472</td><td>0.5175</td><td>0.7571</td><td>0.3260</td><td>1.7583</td></tr>
<tr><td>Gender = female</td><td>-0.4512</td><td>0.2027</td><td>-2.2265</td><td>0.0260</td><td>0.6368</td><td>0.4281</td><td>0.9474</td></tr>
<tr><td>BMI [kg/m²]</td><td>-0.0110</td><td>0.0200</td><td>-0.5484</td><td>0.5834</td><td>0.9891</td><td>0.9510</td><td>1.0286</td></tr>
<tr><td>$eGFR_{sCIV}$ = ≥ 90 mL/min/1.73 m²</td><td>0.5760</td><td>0.4042</td><td>1.4253</td><td>0.1541</td><td>1.7790</td><td>0.8057</td><td>3.9282</td></tr>
<tr><td>$SAPS_{sCIV}$ = ≤36</td><td>0.3892</td><td>0.3988</td><td>0.9760</td><td>0.3291</td><td>1.4758</td><td>0.6755</td><td>3.2244</td></tr>
<tr><td>Sepsis = yes</td><td>-0.0593</td><td>0.2293</td><td>-0.2588</td><td>0.7958</td><td>0.9424</td><td>0.6013</td><td>1.4770</td></tr>
<tr><td>Septic shock = yes</td><td>-0.3025</td><td>0.3468</td><td>-0.8721</td><td>0.3832</td><td>0.7390</td><td>0.3745</td><td>1.4584</td></tr>
<tr><td>Duration of vancomycin therapy [days]</td><td>0.0529</td><td>0.0186</td><td>2.8522</td><td>0.0043</td><td>1.0544</td><td>1.0167</td><td>1.0934</td></tr>
<tr><td>Vasopressors = yes</td><td>-0.0273</td><td>0.2380</td><td>-0.1149</td><td>0.9085</td><td>0.9730</td><td>0.6103</td><td>1.5515</td></tr>
<tr><td>Mechanical ventilation = yes</td><td>-0.0452</td><td>0.2288</td><td>-0.1974</td><td>0.8435</td><td>0.9558</td><td>0.6104</td><td>1.4968</td></tr>
<tr><td>Carbapenem = yes</td><td>-0.0883</td><td>0.2072</td><td>-0.4261</td><td>0.6700</td><td>0.9155</td><td>0.6100</td><td>1.3741</td></tr>
<tr><td>Microbiological failure = yes</td><td>-0.1360</td><td>0.2070</td><td>-0.6570</td><td>0.5112</td><td>0.8729</td><td>0.5818</td><td>1.3096</td></tr>
<tr><td>Bacteraemia with vancomycin-sensitive germ = yes</td><td>-0.0325</td><td>0.2458</td><td>-0.1321</td><td>0.8949</td><td>0.9680</td><td>0.5979</td><td>1.5672</td></tr>
<tr><td>Bone and joint infection = yes</td><td>-0.2164</td><td>0.4052</td><td>-0.5339</td><td>0.5934</td><td>0.8054</td><td>0.3640</td><td>1.7823</td></tr>
<tr><td>CNS infection with vancomycin-sensitive germ = yes</td><td>0.2687</td><td>0.2583</td><td>1.0403</td><td>0.2982</td><td>1.3083</td><td>0.7885</td><td>2.1708</td></tr>
<tr><td>Skin and soft tissue infection = yes</td><td>-0.4173</td><td>0.4397</td><td>-0.9490</td><td>0.3426</td><td>0.6588</td><td>0.2783</td><td>1.5598</td></tr>
<tr><td>Genitourinary infection = yes</td><td>-0.1577</td><td>0.3021</td><td>-0.5221</td><td>0.6016</td><td>0.8541</td><td>0.4725</td><td>1.5439</td></tr>
<tr><td>Pulmonary infection = yes</td><td>0.0327</td><td>0.2128</td><td>0.1537</td><td>0.8779</td><td>1.0332</td><td>0.6809</td><td>1.5678</td></tr>
<tr><td>Gastrointestinal infection = yes</td><td>0.2203</td><td>0.2740</td><td>0.8042</td><td>0.4213</td><td>1.2465</td><td>0.7286</td><td>2.1326</td></tr>
<tr><td>Vancomycin dose day 1 [mg/kg TBW]</td><td>-0.0061</td><td>0.0102</td><td>-0.5986</td><td>0.5495</td><td>0.9939</td><td>0.9742</td><td>1.0140</td></tr>
<tr><td>C72mean = <20 mg/L: $eGFR_{sCIV}$ = ≥90 mL/min/1.73 m²</td><td>-.07306</td><td>0.5177</td><td>-1.4114</td><td>0.1581</td><td>0.4816</td><td>0.1746</td><td>1.3284</td></tr>
<tr><td>C72mean = 20-25 mg/L: $eGFR_{sCIV}$ = ≥90 mL/min/1.73 m²</td><td>-0.5041</td><td>0.5461</td><td>-0.9231</td><td>0.3560</td><td>0.6040</td><td>0.2071</td><td>1.7617</td></tr>
<tr><td>C72mean = <20 mg/L:$SAPS_{sCIV}$ = ≤36</td><td>0.8870</td><td>0.5158</td><td>1.7199</td><td>0.0855</td><td>2.4279</td><td>0.8835</td><td>6.6719</td></tr>
<tr><td>C72mean = 20-25 mg/L:$SAPS_{sCIV}$ = ≤36</td><td>0.6825</td><td>0.5435</td><td>1.2557</td><td>0.2092</td><td>1.9787</td><td>0.6820</td><td>5.7410</td></tr>
<tr><td>$eGFR_{sCIV}$ = ≥90 mL/min/1.73 m²:$SAPS_{sCIV}$ = ≤36</td><td>-0.2473</td><td>0.4431</td><td>-0.5580</td><td>0.5768</td><td>0.7809</td><td>0.3277</td><td>1.8611</td></tr>
</table>

Abbreviations: BMI, body mass index; C72mean, mean vancomycin serum concentration during the first three days of continuous infusion of vancomycin; CNS, central nervous system; $eGFR_{sCIV}$, estimated glomerular filtration rate at the start of continuous infusion of vancomycin; $SAPS_{sCIV}$, Simplified Acute Physiology Score II at the start of continuous infusion of vancomycin; TBW, total body weight.

Reference values: C72mean, >25 mg/L; $eGFR_{sCIV}$, <90 mL/min/1.73 m²; $SAPS_{sCIV}$, >36.

[1]p-value <0.05 was considered significant.

**Table S 14** Estimated model coefficients of the logistic regression for microbiological failure. (n=506, reduced data set, considers only those patients in whom a vancomycin-sensitive germ was detected).

| | Estimate | Std. Error | z value | Pr(>\|z\|)[1] | Odds Ratio | 2.5% | 97.5% |
|---|---|---|---|---|---|---|---|
| (Intercept) | -2.6997 | 0.9493 | -2.8438 | 0.0045 | 0.0672 | 0.0105 | 0.4321 |
| C72mean = <20 mg/L | -0.5921 | 0.4896 | -1.2093 | 0.2265 | 0.5532 | 0.2119 | 1.4442 |
| C72mean = 20-25 mg/L | -0.1064 | 0.4545 | -0.2340 | 0.8150 | 0.8991 | 0.3889 | 2.1911 |
| Gender = female | -0.0073 | 0.2227 | -0.0328 | 0.9739 | 0.9927 | 0.6416 | 1.5360 |
| BMI [kg/m²] | 0.0127 | 0.0193 | 0.6580 | 0.5106 | 1.0128 | 0.9752 | 1.0517 |
| eGFR$_{sCIV}$ = ≥90 mL/min/1.73 m² | -0.1939 | 0.4604 | -0.4211 | 0.6737 | 0.8237 | 0.3341 | 2.0309 |
| SAPS$_{sCIV}$ = ≤36 | 0.5279 | 0.4481 | 1.1781 | 0.2388 | 1.6953 | 0.7045 | 4.0799 |
| Sepsis = yes | 0.6713 | 0.2540 | 2.6432 | **0.0082** | 1.9567 | 1.1895 | 3.2189 |
| Sepsic shock = yes | 1.2703 | 0.3467 | 3.6641 | **0.0002** | 3.5618 | 1.8054 | 7.0270 |
| Duration of vancomycin therapy [days] | 0.0485 | 0.0224 | 2.1703 | **0.0300** | 1.0497 | 1.0047 | 1.0967 |
| Vancomycin dose day 1 [mg/kg TBW] | $-5.927*10^{-5}$ | 0.0113 | -0.0052 | 0.9958 | 0.9999 | 0.9780 | 1.0224 |
| MSSA = yes | 0.4436 | 0.2881 | 1.5399 | 0.1236 | 1.5583 | 0.8860 | 2.7408 |
| MRSA = yes | 0.7007 | 0.3743 | 1.8718 | 0.0612 | 2.0151 | 0.9675 | 4.1969 |
| CoNS = yes | 0.5641 | 0.2604 | 2.1665 | **0.0303** | 1.7578 | 1.0552 | 2.9283 |
| Streptococcus ssp = yes | 0.2377 | 0.3401 | 0.6989 | 0.4846 | 1.2683 | 0.6513 | 2.4698 |
| Enterococcus ssp = yes | 0.6434 | 0.2851 | 2.2566 | **0.0240** | 1.9030 | 1.0883 | 3.3276 |
| Carbapenem = yes | -0.4072 | 0.2280 | -1.7857 | 0.0741 | 0.6655 | 0.4257 | 1.0405 |
| Clinical failure (survival) = yes | -0.3024 | 0.2750 | -1.0996 | 0.2715 | 0.7391 | 0.4312 | 1.2669 |
| Bacteraemia with vancomycin-sensitive germ = yes | 0.7083 | 0.2394 | 2.9587 | **0.0031** | 2.0306 | 1.2701 | 3.2464 |
| Bone and joint infection = yes | 0.0301 | 0.3943 | 0.0764 | 0.9391 | 1.0306 | 0.4758 | 2.2321 |
| CNS infection = yes | -0.0986 | 0.3055 | -0.3228 | 0.7468 | 0.9061 | 0.4979 | 1.6489 |
| Skin and soft tissue infection = yes | 0.6363 | 0.3942 | 1.6140 | 0.1065 | 1.8895 | 0.8725 | 4.0918 |
| Genitourinary infection = yes | 0.2504 | 0.3059 | 0.8184 | 0.4131 | 1.2845 | 0.7052 | 2.3397 |
| Pulmonary infection = yes | 0.7005 | 0.2317 | 3.0235 | **0.0025** | 2.0148 | 1.2794 | 3.1728 |
| Gastrointestinal infection = yes | 0.9355 | 0.3326 | 2.8129 | **0.0049** | 2.5486 | 1.3280 | 4.8909 |
| C72mean = <20 mg/L: eGFR$_{sCIV}$ = ≥90 mL/min/1.73 m² | 1.2853 | 0.5827 | 2.2058 | **0.0274** | 3.6157 | 1.1540 | 11.3284 |
| C72mean = 20-25 mg/L: eGFR$_{sCIV}$ = ≥90 mL/min/1.73 m² | 0.1035 | 0.6185 | 0.1674 | 0.8670 | 1.1091 | 0.3300 | 3.7275 |
| C72mean = <20 mg/L: SAPS$_{sCIV}$ = ≤36 | -0.3572 | 0.5561 | -0.6423 | 0.5207 | 0.6997 | 0.2353 | 2.0808 |
| C72mean = 20-25 mg/L: SAPS$_{sCIV}$ = ≤36 | -0.4239 | 0.6135 | -0.6909 | 0.4896 | 0.6545 | 0.1966 | 2.1784 |
| eGFR$_{sCIV}$ = ≥90 mL/min/1.73 m²: SAPS$_{sCIV}$ = ≤36 | 0.0240 | 0.5006 | 0.0480 | 0.9617 | 1.0243 | 0.3840 | 2.7322 |

Abbreviations: BMI, body mass index; C72mean, mean vancomycin serum concentration during the first three days of continuous infusion of vancomycin; CNS, central nervous system; CoNS, coagulase-negative staphylococci; eGFR$_{sCIV}$, estimated glomerular filtration rate at the start of continuous infusion of vancomycin; MRSA, methicillin-resistant Staphylococcus aureus; MSSA, methicillin-sensitive Staphylococcus aureus; SAPS$_{sCIV}$, Simplified Acute Physiology Score II at the start of continuous infusion of vancomycin; TBW, total body weight.

Reference values: C72mean, >25 mg/L; eGFR$_{sCIV}$, <90 mL/min/1.73 m²; SAPS$_{sCIV}$, >36.

[1]p-value <0.05 was considered significant.

**Table S 15** Quality measures based on a 10-fold cross-validation with optimised cut-off values using the Youden index determined in the course of multivariate logistic regression analysis.

| Outcome | Error Rate | Sensitivity | Specificity |
|---|---|---|---|
| ICU mortality | 0.2863 | 0.7962 | 0.6967 |
| In-hospital mortality | 0.2690 | 0.7111 | 0.7374 |
| 30-day mortality | 0.2484 | 0.6807 | 0.7672 |
| Total AKI | 0.3048 | 0.7572 | 0.6809 |
| Early AKI | 0.3200 | 0.8714 | 0.6643 |
| Late AKI | 0.3980 | 0.7767 | 0.5800 |
| Clinical failure (survival) | 0.4056 | 0.6748 | 0.5771 |
| Microbiological failure | 0.3339 | 0.5322 | 0.7207 |

Abbreviations: AKI, acute kidney injury; ICU, intensive care unit.

### 1.10 Post-Hoc-Tables Outcomes and Css

#### *1.10.1 ICU Mortality*

**Table S 16** Post-hoc comparison of the classes of mean vancomycin serum concentration during the first three days of continuous infusion of vancomycin (C72mean) within the subgroups for estimated glomerular filtration rate at the start of continuous infusion of vancomycin ($eGFR_{SCIV}$) and Simplified Acute Physiology Score II at the start of continuous infusion of vancomycin ($SAPS_{SCIV}$) for intensive care unit (ICU) mortality with Bonferroni correction based on multivariate logistic regression.

| | C72mean contrast | $eGFR_{SCIV}$ | $SAPS_{SCIV}$ | OR | SE | df | null | z ratio | p-value[1] |
|---|---|---|---|---|---|---|---|---|---|
| Subgroup A (n = 262) | [20, 25] / < 20 | < 90 | > 36 | 1.7188 | 0.8527 | ∞ | 1 | 1.0917 | 0.8249 |
| | > 25 / < 20 | < 90 | > 36 | 1.8337 | 0.8016 | ∞ | 1 | 1.3870 | 0.4964 |
| | > 25 / [20, 25] | < 90 | > 36 | 1.0669 | 0.3881 | ∞ | 1 | 0.1779 | 1.0000 |
| Subgroup B (n = 186) | [20, 25] / < 20 | ≥ 90 | > 36 | 1.9966 | 0.9692 | ∞ | 1 | 1.4243 | 0.4630 |
| | > 25 / < 20 | ≥ 90 | > 36 | 1.0806 | 0.5237 | ∞ | 1 | 0.1600 | 1.0000 |
| | > 25 / [20, 25] | ≥ 90 | > 36 | 0.5412 | 0.2830 | ∞ | 1 | -1.1742 | 0.7210 |
| Subgroup C (n = 105) | [20, 25] / < 20 | < 90 | ≤ 36 | 0.5805 | 0.4147 | ∞ | 1 | -0.7613 | 1.0000 |
| | > 25 / < 20 | < 90 | ≤ 36 | 2.4705 | 1.5207 | ∞ | 1 | 1.4693 | 0.4253 |
| | > 25 / [20, 25] | < 90 | ≤ 36 | 4.2555 | 2.9490 | ∞ | 1 | 2.0898 | 0.1099 |
| Subgroup D (n = 369) | [20, 25] / < 20 | ≥ 90 | ≤ 36 | 0.6744 | 0.3130 | ∞ | 1 | -0.8489 | 1.0000 |
| | > 25 / < 20 | ≥ 90 | ≤ 36 | 1.4559 | 0.7554 | ∞ | 1 | 0.7240 | 1.0000 |
| | > 25 / [20, 25] | ≥ 90 | ≤ 36 | 2.1589 | 1.2667 | ∞ | 1 | 1.3116 | 0.5690 |

Abbreviations: C72mean, mean vancomycin serum concentration during the first three days of continuous infusion of vancomycin [mg/L]; df, degree of freedom; $eGFR_{SCIV}$, estimated glomerular filtration rate at the start of continuous infusion of vancomycin [mL/min/1.73 m²]; OR, odds ratio; $SAPS_{SCIV}$, Simplified Acute Physiology Score II at the start of continuous infusion of vancomycin; SE, standard error.

[1] p-value <0.05 was considered significant.

**Table S 17** Post-hoc comparison of the classes of mean vancomycin serum concentration during the first three days of continuous infusion of vancomycin (C72mean) within the subgroups for estimated glomerular filtration rate at the start of continuous infusion of vancomycin ($eGFR_{SCIV}$) and Simplified Acute Physiology Score II at the start of continuous infusion of vancomycin ($SAPS_{SCIV}$) for intensive care unit (ICU) mortality with Bonferroni correction based on multivariate Cox regression.

| | C72mean contrast | $eGFR_{SCIV}$ | $SAPS_{SCIV}$ | HR | SE | df | null | z ratio | p-value[1] |
|---|---|---|---|---|---|---|---|---|---|
| Subgroup A (n = 262) | [20. 25] / < 20 | < 90 | > 36 | 1.2703 | 0.5289 | ∞ | 1 | 0.5747 | 1.0000 |
| | > 25 / < 20 | < 90 | > 36 | 1.0393 | 0.3897 | ∞ | 1 | 0.1029 | 1.0000 |
| | > 25 / [20. 25] | < 90 | > 36 | 0.8182 | 0.2363 | ∞ | 1 | -0.6949 | 1.0000 |
| Subgroup B (n = 186) | [20. 25] / < 20 | ≥ 90 | > 36 | 1.7069 | 0.7248 | ∞ | 1 | 1.2592 | 0.6239 |
| | > 25 / < 20 | ≥ 90 | > 36 | 0.8581 | 0.3545 | ∞ | 1 | -0.3705 | 1.0000 |
| | > 25 / [20. 25] | ≥ 90 | > 36 | 0.5027 | 0.2209 | ∞ | 1 | -1.5649 | 0.3528 |
| Subgroup C (n = 105) | [20. 25] / < 20 | < 90 | ≤ 36 | 0.4886 | 0.3172 | ∞ | 1 | -1.1032 | 0.8098 |
| | > 25 / < 20 | < 90 | ≤ 36 | 1.5545 | 0.8577 | ∞ | 1 | 0.7995 | 1.0000 |
| | > 25 / [20. 25] | < 90 | ≤ 36 | 3.1815 | 2.0089 | ∞ | 1 | 1.8329 | 0.2005 |
| Subgroup D (n = 369) | [20. 25] / < 20 | ≥ 90 | ≤ 36 | 0.6565 | 0.2766 | ∞ | 1 | -0.9986 | 0.9540 |
| | > 25 / < 20 | ≥ 90 | ≤ 36 | 1.2834 | 0.5813 | ∞ | 1 | 0.5508 | 1.0000 |
| | > 25 / [20. 25] | ≥ 90 | ≤ 36 | 1.9548 | 1.0271 | ∞ | 1 | 1.2756 | 0.6062 |

Abbreviations: C72mean, mean vancomycin serum concentration during the first three days of continuous infusion of vancomycin [mg/L]; df, degree of freedom; $eGFR_{SCIV}$, estimated glomerular filtration rate at the start of continuous infusion of vancomycin; HR, hazard ratio; $SAPS_{SCIV}$, Simplified Acute Physiology Score II at the start of continuous infusion of vancomycin; SE, standard error.

[1] p-value <0.05 was considered significant.

#### *1.10.2 In-hospital mortality*

**Table S 18** Post-hoc comparison of the classes of mean vancomycin serum concentration during the first three days of continuous infusion of vancomycin (C72mean) within the subgroups for estimated glomerular filtration rate at the start of continuous infusion of vancomycin ($eGFR_{sCIV}$) and Simplified Acute Physiology Score II at the start of continuous infusion of vancomycin ($SAPS_{sCIV}$) for in-hospital mortality with Bonferroni correction based on multivariate logistic regression.

| | C72mean contrast | $eGFR_{sCIV}$ | $SAPS_{sCIV}$ | OR | SE | df | null | z ratio | p-value[1] |
|---|---|---|---|---|---|---|---|---|---|
| Subgroup A (n = 262) | [20. 25] / < 20 | < 90 | > 36 | 1.6799 | 0.7276 | ∞ | 1 | 1.1978 | 0.6930 |
| | > 25 / < 20 | < 90 | > 36 | 2.1609 | 0.8286 | ∞ | 1 | 2.0095 | 0.1334 |
| | > 25 / [20. 25] | < 90 | > 36 | 1.2863 | 0.4270 | ∞ | 1 | 0.7584 | 1.0000 |
| Subgroup B (n = 186) | [20. 25] / < 20 | ≥ 90 | > 36 | 1.5908 | 0.6772 | ∞ | 1 | 1.0906 | 0.8263 |
| | > 25 / < 20 | ≥ 90 | > 36 | 1.1955 | 0.5097 | ∞ | 1 | 0.4189 | 1.0000 |
| | > 25 / [20. 25] | ≥ 90 | > 36 | 0.7515 | 0.3486 | ∞ | 1 | -0.6158 | 1.0000 |
| Subgroup C (n = 105) | [20. 25] / < 20 | < 90 | ≤ 36 | 0.9910 | 0.5664 | ∞ | 1 | -0.0158 | 1.0000 |
| | > 25 / < 20 | < 90 | ≤ 36 | 2.4051 | 1.2449 | ∞ | 1 | 1.6956 | 0.2699 |
| | > 25 / [20. 25] | < 90 | ≤ 36 | 2.4269 | 1.3343 | ∞ | 1 | 1.6127 | 0.3204 |
| Subgroup D (n = 369) | [20. 25] / < 20 | ≥ 90 | ≤ 36 | 0.9384 | 0.3432 | ∞ | 1 | -0.1737 | 1.0000 |
| | > 25 / < 20 | ≥ 90 | ≤ 36 | 1.3307 | 0.6041 | ∞ | 1 | 0.6293 | 1.0000 |
| | > 25 / [20. 25] | ≥ 90 | ≤ 36 | 1.4179 | 0.6917 | ∞ | 1 | 0.7158 | 1.0000 |

Abbreviations: C72mean, mean vancomycin serum concentration during the first three days of continuous infusion of vancomycin [mg/L]; df, degree of freedom; $eGFR_{sCIV}$, estimated glomerular filtration rate at the start of continuous infusion of vancomycin [mL/min/1.73 m²]; OR, odds ratio; $SAPS_{sCIV}$, Simplified Acute Physiology Score II at the start of continuous infusion of vancomycin; SE, standard error.

[1] p-value <0.05 was considered significant.

**Table S 19** Post-hoc comparison of the classes of mean vancomycin serum concentration during the first three days of continuous infusion of vancomycin (C72mean) within the subgroups for estimated glomerular filtration rate at the start of continuous infusion of vancomycin ($eGFR_{sCIV}$) and Simplified Acute Physiology Score II at the start of continuous infusion of vancomycin ($SAPS_{sCIV}$) for in-hospital mortality with Bonferroni correction based on multivariate Cox regression.

| | C72mean contrast | $eGFR_{sCIV}$ | $SAPS_{sCIV}$ | HR | SE | df | null | z ratio | p-value[1] |
|---|---|---|---|---|---|---|---|---|---|
| Subgroup A (n = 262) | [20. 25] / < 20 | < 90 | > 36 | 1.2584 | 0.4328 | ∞ | 1 | 0.6681 | 1.0000 |
| | > 25 / < 20 | < 90 | > 36 | 1.2669 | 0.3916 | ∞ | 1 | 0.7654 | 1.0000 |
| | > 25 / [20. 25] | < 90 | > 36 | 1.0068 | 0.2388 | ∞ | 1 | 0.0286 | 1.0000 |
| Subgroup B (n = 186) | [20. 25] / < 20 | ≥ 90 | > 36 | 1.3836 | 0.4933 | ∞ | 1 | 0.9105 | 1.0000 |
| | > 25 / < 20 | ≥ 90 | > 36 | 0.8708 | 0.2953 | ∞ | 1 | -0.4079 | 1.0000 |
| | > 25 / [20. 25] | ≥ 90 | > 36 | 0.6294 | 0.2301 | ∞ | 1 | -1.2665 | 0.6160 |
| Subgroup C (n = 105) | [20. 25] / < 20 | < 90 | ≤ 36 | 0.7822 | 0.3838 | ∞ | 1 | -0.5006 | 1.0000 |
| | > 25 / < 20 | < 90 | ≤ 36 | 1.9295 | 0.8409 | ∞ | 1 | 1.5081 | 0.3946 |
| | > 25 / [20. 25] | < 90 | ≤ 36 | 2.4668 | 1.1424 | ∞ | 1 | 1.9498 | 0.1536 |
| Subgroup D (n = 369) | [20. 25] / < 20 | ≥ 90 | ≤ 36 | 0.8600 | 0.2730 | ∞ | 1 | -0.4751 | 1.0000 |
| | > 25 / < 20 | ≥ 90 | ≤ 36 | 1.3263 | 0.4971 | ∞ | 1 | 0.7533 | 1.0000 |
| | > 25 / [20. 25] | ≥ 90 | ≤ 36 | 1.5421 | 0.6299 | ∞ | 1 | 1.0606 | 0.8666 |

Abbreviations: C72mean, mean vancomycin serum concentration during the first three days of continuous infusion of vancomycin [mg/L]; df, degree of freedom; $eGFR_{sCIV}$, estimated glomerular filtration rate at the start of continuous infusion of vancomycin; HR, hazard ratio; $SAPS_{sCIV}$, Simplified Acute Physiology Score II at the start of continuous infusion of vancomycin; SE, standard error.

[1] p-value <0.05 was considered significant.

#### *1.10.3 30-day mortality*

**Table S 20** Post-hoc comparison of the classes of mean vancomycin serum concentration during the first three days of continuous infusion of vancomycin (C72mean) within the subgroups for estimated glomerular filtration rate at the start of continuous infusion of vancomycin ($eGFR_{sCIV}$) and Simplified Acute Physiology Score II at the start of continuous infusion of vancomycin ($SAPS_{sCIV}$) for 30-day mortality with Bonferroni correction based on multivariate logistic regression.

| | C72mean contrast | $eGFR_{sCIV}$ | $SAPS_{sCIV}$ | OR | SE | df | null | z ratio | p-value[1] |
|---|---|---|---|---|---|---|---|---|---|
| Subgroup A (n = 262) | [20. 25] / < 20 | < 90 | > 36 | 1.2445 | 0.5888 | ∞ | 1 | 0.4624 | 1.0000 |
| | > 25 / < 20 | < 90 | > 36 | 1.6871 | 0.7012 | ∞ | 1 | 1.2583 | 0.6248 |
| | > 25 / [20. 25] | < 90 | > 36 | 1.3556 | 0.4798 | ∞ | 1 | 0.8597 | 1.0000 |
| Subgroup B (n = 186) | [20. 25] / < 20 | ≥ 90 | > 36 | 1.0942 | 0.5223 | ∞ | 1 | 0.1887 | 1.0000 |
| | > 25 / < 20 | ≥ 90 | > 36 | 0.2549 | 0.1447 | ∞ | 1 | -2.4074 | **0.0482** |
| | > 25 / [20. 25] | ≥ 90 | > 36 | 0.2329 | 0.1428 | ∞ | 1 | -2.3766 | **0.0524** |
| Subgroup C (n = 105) | [20. 25] / < 20 | < 90 | ≤ 36 | 0.7473 | 0.5073 | ∞ | 1 | -0.4291 | 1.0000 |
| | > 25 / < 20 | < 90 | ≤ 36 | 3.9907 | 2.4813 | ∞ | 1 | 2.2258 | **0.0781** |
| | > 25 / [20. 25] | < 90 | ≤ 36 | 0.5304 | 3.6715 | ∞ | 1 | 2.4366 | **0.0445** |
| Subgroup D (n = 369) | [20. 25] / < 20 | ≥ 90 | ≤ 36 | 0.6570 | 0.2721 | ∞ | 1 | -1.0142 | 0.9315 |
| | > 25 / < 20 | ≥ 90 | ≤ 36 | 0.6028 | 0.3374 | ∞ | 1 | -0.9043 | 1.0000 |
| | > 25 / [20. 25] | ≥ 90 | ≤ 36 | 0.9175 | 0.5568 | ∞ | 1 | -0.1420 | 1.0000 |

Abbreviations: C72mean, mean vancomycin serum concentration during the first three days of continuous infusion of vancomycin [mg/L]; df, degree of freedom; $eGFR_{sCIV}$, estimated glomerular filtration rate at the start of continuous infusion of vancomycin [mL/min/1.73 m$^2$]; OR, odds ratio; $SAPS_{sCIV}$, Simplified Acute Physiology Score II at the start of continuous infusion of vancomycin; SE, standard error.

[1] p-value <0.05 was considered significant.

**Table S 21** Post-hoc comparison of the classes of mean vancomycin serum concentration during the first three days of continuous infusion of vancomycin (C72mean) within the subgroups for estimated glomerular filtration rate at the start of continuous infusion of vancomycin ($eGFR_{sCIV}$) and Simplified Acute Physiology Score II at the start of continuous infusion of vancomycin ($SAPS_{sCIV}$) for 30-day mortality with Bonferroni correction based on multivariate Cox regression.

| | C72mean contrast | $eGFR_{sCIV}$ | $SAPS_{sCIV}$ | HR | SE | df | null | z ratio | p-value[1] |
|---|---|---|---|---|---|---|---|---|---|
| Subgroup A (n = 262) | [20. 25] / < 20 | < 90 | > 36 | 1.1330 | 0.4537 | ∞ | 1 | 0.3119 | 1.0000 |
| | > 25 / < 20 | < 90 | > 36 | 1.2654 | 0.4480 | ∞ | 1 | 0.6649 | 1.0000 |
| | > 25 / [20. 25] | < 90 | > 36 | 1.1168 | 0.3154 | ∞ | 1 | 0.3913 | 1.0000 |
| Subgroup B (n = 186) | [20. 25] / < 20 | ≥ 90 | > 36 | 1.0241 | 0.4296 | ∞ | 1 | 0.0568 | 1.0000 |
| | > 25 / < 20 | ≥ 90 | > 36 | 0.2976 | 0.1511 | ∞ | 1 | -2.3870 | **0.0510** |
| | > 25 / [20. 25] | ≥ 90 | > 36 | 0.2906 | 0.1592 | ∞ | 1 | -2.2560 | **0.0722** |
| Subgroup C (n = 105) | [20. 25] / < 20 | < 90 | ≤ 36 | 0.7068 | 0.4296 | ∞ | 1 | -0.5709 | 1.0000 |
| | > 25 / < 20 | < 90 | ≤ 36 | 2.9808 | 1.6716 | ∞ | 1 | 1.9475 | 0.1544 |
| | > 25 / [20. 25] | < 90 | ≤ 36 | 4.2171 | 2.6295 | ∞ | 1 | 2.3080 | **0.0630** |
| Subgroup D (n = 369) | [20. 25] / < 20 | ≥ 90 | ≤ 36 | 0.6389 | 0.2421 | ∞ | 1 | -1.1825 | 0.7111 |
| | > 25 / < 20 | ≥ 90 | ≤ 36 | 0.7009 | 0.3497 | ∞ | 1 | -0.7124 | 1.0000 |
| | > 25 / [20. 25] | ≥ 90 | ≤ 36 | 1.0971 | 0.6025 | ∞ | 1 | 0.1688 | 1.0000 |

Abbreviations: C72mean, mean vancomycin serum concentration during the first three days of continuous infusion of vancomycin [mg/L]; df, degree of freedom; $eGFR_{sCIV}$, estimated glomerular filtration rate at the start of continuous infusion of vancomycin; HR, hazard ratio; $SAPS_{sCIV}$, Simplified Acute Physiology Score II at the start of continuous infusion of vancomycin; SE, standard error.

[1] p-value <0.05 was considered significant.

#### *1.10.4 Total AKI*

**Table S 22** Post-hoc comparison of the classes of mean vancomycin serum concentration during the first three days of continuous infusion of vancomycin (C72mean) within the subgroups for estimated glomerular filtration rate at the start of continuous infusion of vancomycin (eGFR$_{sCIV}$) and Simplified Acute Physiology Score II at the start of continuous infusion of vancomycin (SAPS$_{sCIV}$) for total acute kidney injury (AKI) with Bonferroni correction based on multivariate logistic regression.

| | C72mean contrast | eGFR$_{sCIV}$ | SAPS$_{sCIV}$ | OR | SE | df | null | z ratio | p-value[1] |
|---|---|---|---|---|---|---|---|---|---|
| Subgroup A (n = 262) | [20. 25] / < 20 | < 90 | > 36 | 1.7342 | 0.8566 | ∞ | 1 | 1.1147 | 0.7950 |
| | > 25 / < 20 | < 90 | > 36 | 2.8378 | 1.2390 | ∞ | 1 | 2.3889 | **0.0507** |
| | > 25 / [20. 25] | < 90 | > 36 | 1.6363 | 0.5657 | ∞ | 1 | 1.4245 | 0.4629 |
| Subgroup B (n = 186) | [20. 25] / < 20 | ≥ 90 | > 36 | 1.1176 | 0.5269 | ∞ | 1 | 0.2358 | 1.0000 |
| | > 25 / < 20 | ≥ 90 | > 36 | 2.3424 | 1.0333 | ∞ | 1 | 1.9296 | 0.1610 |
| | > 25 / [20. 25] | ≥ 90 | > 36 | 2.0959 | 1.0078 | ∞ | 1 | 1.5389 | 0.3715 |
| Subgroup C (n = 105) | [20. 25] / < 20 | < 90 | ≤ 36 | 3.8919 | 2.6044 | ∞ | 1 | 2.0307 | 0.1269 |
| | > 25 / < 20 | < 90 | ≤ 36 | 12.3570 | 7.6626 | ∞ | 1 | 4.0545 | **0.0002** |
| | > 25 / [20. 25] | < 90 | ≤ 36 | 3.1751 | 1.7777 | ∞ | 1 | 2.0634 | 0.1172 |
| Subgroup D (n = 369) | [20. 25] / < 20 | ≥ 90 | ≤ 36 | 2.5080 | 0.9460 | ∞ | 1 | 2.4379 | **0.0443** |
| | > 25 / < 20 | ≥ 90 | ≤ 36 | 10.1998 | 4.3964 | ∞ | 1 | 5.3880 | **<0.0001** |
| | > 25 / [20. 25] | ≥ 90 | ≤ 36 | 4.0669 | 1.7781 | ∞ | 1 | 3.2086 | **0.0040** |

Abbreviations: C72mean, mean vancomycin serum concentration during the first three days of continuous infusion of vancomycin [mg/L]; df, degree of freedom; eGFR$_{sCIV}$, estimated glomerular filtration rate at the start of continuous infusion of vancomycin [mL/min/1.73 m$^2$]; OR, odds ratio; SAPS$_{sCIV}$, Simplified Acute Physiology Score II at the start of continuous infusion of vancomycin; SE, standard error.

[1] p-value <0.05 was considered significant.

**Table S 23** Post-hoc comparison of the classes of mean vancomycin serum concentration during the first three days of continuous infusion of vancomycin (C72mean) within the subgroups for estimated glomerular filtration rate at the start of continuous infusion of vancomycin (eGFR$_{sCIV}$) and Simplified Acute Physiology Score II at the start of continuous infusion of vancomycin (SAPS$_{sCIV}$) for total acute kidney injury (AKI) with Bonferroni correction based on multivariate Cox regression.

| | C72mean contrast | eGFR$_{sCIV}$ | SAPS$_{sCIV}$ | HR | SE | df | null | z ratio | p-value[1] |
|---|---|---|---|---|---|---|---|---|---|
| Subgroup A (n = 262) | [20. 25] / < 20 | < 90 | > 36 | 1.4684 | 0.6259 | ∞ | 1 | 0.9013 | 1.0000 |
| | > 25 / < 20 | < 90 | > 36 | 2.3764 | 0.8908 | ∞ | 1 | 2.3091 | **0.0628** |
| | > 25 / [20. 25] | < 90 | > 36 | 1.6184 | 0.4627 | ∞ | 1 | 1.6838 | 0.2766 |
| Subgroup B (n = 186) | [20. 25] / < 20 | ≥ 90 | > 36 | 1.0724 | 0.4545 | ∞ | 1 | 0.1649 | 1.0000 |
| | > 25 / < 20 | ≥ 90 | > 36 | 1.9398 | 0.7297 | ∞ | 1 | 1.7612 | 0.2346 |
| | > 25 / [20. 25] | ≥ 90 | > 36 | 1.8088 | 0.7494 | ∞ | 1 | 1.4304 | 0.4578 |
| Subgroup C (n = 105) | [20. 25] / < 20 | < 90 | ≤ 36 | 3.2475 | 1.9646 | ∞ | 1 | 1.9470 | 0.1546 |
| | > 25 / < 20 | < 90 | ≤ 36 | 10.1213 | 5.5076 | ∞ | 1 | 4.2536 | **<0.0001** |
| | > 25 / [20. 25] | < 90 | ≤ 36 | 3.1167 | 1.5181 | ∞ | 1 | 2.3338 | 0.0588 |
| Subgroup D (n = 369) | [20. 25] / < 20 | ≥ 90 | ≤ 36 | 2.3717 | 0.8067 | ∞ | 1 | 2.5390 | **0.0333** |
| | > 25 / < 20 | ≥ 90 | ≤ 36 | 8.2615 | 2.9566 | ∞ | 1 | 5.9004 | **<0.0001** |
| | > 25 / [20. 25] | ≥ 90 | ≤ 36 | 3.4834 | 1.2387 | ∞ | 1 | 3.5096 | **0.0013** |

Abbreviations: C72mean, mean vancomycin serum concentration during the first three days of continuous infusion of vancomycin [mg/L]; df, degree of freedom; eGFR$_{sCIV}$, estimated glomerular filtration rate at the start of continuous infusion of vancomycin; HR, hazard ratio; SAPS$_{sCIV}$, Simplified Acute Physiology Score II at the start of continuous infusion of vancomycin; SE, standard error.

[1] p-value <0.05 was considered significant.

#### *1.10.5 Early AKI*

**Table S 24** Post-hoc comparison of the classes of mean vancomycin serum concentration during the first three days of continuous infusion of vancomycin (C72mean) within the subgroups for estimated glomerular filtration rate at the start of continuous infusion of vancomycin ($eGFR_{sCIV}$) and Simplified Acute Physiology Score II at the start of continuous infusion of vancomycin ($SAPS_{sCIV}$) for early acute kidney injury (AKI) with Bonferroni correction based on multivariate logistic regression.

| | C72mean contrast | $eGFR_{sCIV}$ | $SAPS_{sCIV}$ | OR | SE | df | null | z ratio | p-value[1] |
|---|---|---|---|---|---|---|---|---|---|
| Subgroup A (n = 262) | [20. 25] / < 20 | < 90 | > 36 | 0.4852 | 0.4028 | ∞ | 1 | -0.8712 | 1.0000 |
| | > 25 / < 20 | < 90 | > 36 | 2.4318 | 1.4784 | ∞ | 1 | 1.4617 | 0.4315 |
| | > 25 / [20. 25] | < 90 | > 36 | 5.0119 | 3.2875 | ∞ | 1 | 2.4573 | **0.0420** |
| Subgroup B (n = 186) | [20. 25] / < 20 | ≥ 90 | > 36 | 0.8706 | 0.6196 | ∞ | 1 | -0.1947 | 1.0000 |
| | > 25 / < 20 | ≥ 90 | > 36 | 1.7537 | 1.0902 | ∞ | 1 | 0.9035 | 1.0000 |
| | > 25 / [20. 25] | ≥ 90 | > 36 | 2.0143 | 1.4928 | ∞ | 1 | 0.9450 | 1.0000 |
| Subgroup C (n = 105) | [20. 25] / < 20 | < 90 | ≤ 36 | 3.3311 | 4.2104 | ∞ | 1 | 0.9520 | 1.0000 |
| | > 25 / < 20 | < 90 | ≤ 36 | 43.9635 | 47.6184 | ∞ | 1 | 3.4930 | **0.0014** |
| | > 25 / [20. 25] | < 90 | ≤ 36 | 13.1980 | 14.4535 | ∞ | 1 | 2.3560 | **0.0554** |
| Subgroup D (n = 369) | [20. 25] / < 20 | ≥ 90 | ≤ 36 | 5.9769 | 4.3789 | ∞ | 1 | 2.4404 | **0.0440** |
| | > 25 / < 20 | ≥ 90 | ≤ 36 | 31.7039 | 23.6433 | ∞ | 1 | 4.6348 | **<0.0001** |
| | > 25 / [20. 25] | ≥ 90 | ≤ 36 | 5.3044 | 3.3100 | ∞ | 1 | 2.6738 | **0.0225** |

Abbreviations: C72mean, mean vancomycin serum concentration during the first three days of continuous infusion of vancomycin [mg/L]; df, degree of freedom; $eGFR_{sCIV}$, estimated glomerular filtration rate at the start of continuous infusion of vancomycin [mL/min/1.73 m²]; OR, odds ratio; $SAPS_{sCIV}$, Simplified Acute Physiology Score II at the start of continuous infusion of vancomycin; SE, standard error.

[1] p-value <0.05 was considered significant.

**Table S 25** Post-hoc comparison of the classes of mean vancomycin serum concentration during the first three days of continuous infusion of vancomycin (C72mean) within the subgroups for estimated glomerular filtration rate at the start of continuous infusion of vancomycin ($eGFR_{sCIV}$) and Simplified Acute Physiology Score II at the start of continuous infusion of vancomycin ($SAPS_{sCIV}$) for early acute kidney injury (AKI) with Bonferroni correction based on multivariate Cox regression.

| | C72mean contrast | $eGFR_{sCIV}$ | $SAPS_{sCIV}$ | HR | SE | df | null | z ratio | p-value[1] |
|---|---|---|---|---|---|---|---|---|---|
| Subgroup A (n = 262) | [20. 25] / < 20 | < 90 | > 36 | 0.5157 | 0.4052 | ∞ | 1 | -0.8427 | 1.0000 |
| | > 25 / < 20 | < 90 | > 36 | 2.2157 | 1.2404 | ∞ | 1 | 1.4211 | 0.4658 |
| | > 25 / [20. 25] | < 90 | > 36 | 4.2966 | 2.6835 | ∞ | 1 | 2.3341 | 0.0588 |
| Subgroup B (n = 186) | [20. 25] / < 20 | ≥ 90 | > 36 | 0.8184 | 0.5505 | ∞ | 1 | -0.2979 | 1.0000 |
| | > 25 / < 20 | ≥ 90 | > 36 | 1.4526 | 0.8346 | ∞ | 1 | 0.6498 | 1.0000 |
| | > 25 / [20. 25] | ≥ 90 | > 36 | 1.7748 | 1.2251 | ∞ | 1 | 0.8311 | 1.0000 |
| Subgroup C (n = 105) | [20. 25] / < 20 | < 90 | ≤ 36 | 3.4530 | 4.1723 | ∞ | 1 | 1.0256 | 0.9152 |
| | > 25 / < 20 | < 90 | ≤ 36 | 40.2197 | 40.6900 | ∞ | 1 | 3.6517 | **0.0008** |
| | > 25 / [20. 25] | < 90 | ≤ 36 | 11.6477 | 11.8701 | ∞ | 1 | 2.4091 | **0.0480** |
| Subgroup D (n = 369) | [20. 25] / < 20 | ≥ 90 | ≤ 36 | 5.4800 | 3.9050 | ∞ | 1 | 2.3872 | **0.0509** |
| | > 25 / < 20 | ≥ 90 | ≤ 36 | 26.3664 | 18.4129 | ∞ | 1 | 4.6855 | **<0.0001** |
| | > 25 / [20. 25] | ≥ 90 | ≤ 36 | 4.8114 | 2.6844 | ∞ | 1 | 2.8158 | **0.0146** |

Abbreviations: C72mean, mean vancomycin serum concentration during the first three days of continuous infusion of vancomycin [mg/L]; df, degree of freedom; $eGFR_{sCIV}$, estimated glomerular filtration rate at the start of continuous infusion of vancomycin; HR, hazard ratio; $SAPS_{sCIV}$, Simplified Acute Physiology Score II at the start of continuous infusion of vancomycin; SE, standard error.

[1] p-value <0.05 was considered significant.

#### *1.10.6 Late AKI*

**Table S 26** Post-hoc comparison of the classes of mean vancomycin serum concentration during the first three days of continuous infusion of vancomycin (C72mean) within the subgroups for estimated glomerular filtration rate at the start of continuous infusion of vancomycin ($eGFR_{sCIV}$) and Simplified Acute Physiology Score II at the start of continuous infusion of vancomycin ($SAPS_{sCIV}$) for late acute kidney injury (AKI) with Bonferroni correction based on multivariate logistic regression.

| | C72mean contrast | $eGFR_{sCIV}$ | $SAPS_{sCIV}$ | OR | SE | df | null | z ratio | p-value[1] |
|---|---|---|---|---|---|---|---|---|---|
| Subgroup A (n = 262) | [20. 25] / < 20 | < 90 | > 36 | 3.0064 | 1.7925 | ∞ | 1 | 1.8462 | 0.1946 |
| | > 25 / < 20 | < 90 | > 36 | 2.5513 | 1.4138 | ∞ | 1 | 1.6901 | 0.2730 |
| | > 25 / [20. 25] | < 90 | > 36 | 0.8486 | 0.3305 | ∞ | 1 | -0.4215 | 1.0000 |
| Subgroup B (n = 186) | [20. 25] / < 20 | ≥ 90 | > 36 | 1.4596 | 0.8413 | ∞ | 1 | 0.6561 | 1.0000 |
| | > 25 / < 20 | ≥ 90 | > 36 | 1.9678 | 1.0975 | ∞ | 1 | 1.2136 | 0.6747 |
| | > 25 / [20. 25] | ≥ 90 | > 36 | 1.3481 | 0.7830 | ∞ | 1 | 0.5143 | 1.0000 |
| Subgroup C (n = 105) | [20. 25] / < 20 | < 90 | ≤ 36 | 3.1557 | 2.3902 | ∞ | 1 | 1.5173 | 0.3876 |
| | > 25 / < 20 | < 90 | ≤ 36 | 4.9320 | 3.5276 | ∞ | 1 | 2.2311 | **0.0770** |
| | > 25 / [20. 25] | < 90 | ≤ 36 | 1.5629 | 0.9730 | ∞ | 1 | 0.7172 | 1.0000 |
| Subgroup D (n = 369) | [20. 25] / < 20 | ≥ 90 | ≤ 36 | 1.5322 | 0.6791 | ∞ | 1 | 0.9627 | 1.0000 |
| | > 25 / < 20 | ≥ 90 | ≤ 36 | 3.8040 | 1.9343 | ∞ | 1 | 2.6274 | **0.0258** |
| | > 25 / [20. 25] | ≥ 90 | ≤ 36 | 2.4828 | 1.3280 | ∞ | 1 | 1.7001 | 0.2674 |

Abbreviations: C72mean, mean vancomycin serum concentration during the first three days of continuous infusion of vancomycin [mg/L]; df, degree of freedom; $eGFR_{sCIV}$, estimated glomerular filtration rate at the start of continuous infusion of vancomycin [mL/min/1.73 $m^2$]; OR, odds ratio; $SAPS_{sCIV}$, Simplified Acute Physiology Score II at the start of continuous infusion of vancomycin; SE, standard error.

[1] p-value <0.05 was considered significant.

**Table S 27** Post-hoc comparison of the classes of mean vancomycin serum concentration during the first three days of continuous infusion of vancomycin (C72mean) within the subgroups for estimated glomerular filtration rate at the start of continuous infusion of vancomycin ($eGFR_{sCIV}$) and Simplified Acute Physiology Score II at the start of continuous infusion of vancomycin ($SAPS_{sCIV}$) for late acute kidney injury (AKI) with Bonferroni correction based on multivariate Cox regression.

| | C72mean contrast | $eGFR_{sCIV}$ | $SAPS_{sCIV}$ | HR | SE | df | null | z ratio | p-value[1] |
|---|---|---|---|---|---|---|---|---|---|
| Subgroup A (n = 262) | [20. 25] / < 20 | < 90 | > 36 | 2.4756 | 1.3626 | ∞ | 1 | 1.6469 | 0.2987 |
| | > 25 / < 20 | < 90 | > 36 | 2.6322 | 1.3662 | ∞ | 1 | 1.8705 | 0.1842 |
| | > 25 / [20. 25] | < 90 | > 36 | 1.0649 | 0.3636 | ∞ | 1 | 0.1841 | 1.0000 |
| Subgroup B (n = 186) | [20. 25] / < 20 | ≥ 90 | > 36 | 1.4595 | 0.7923 | ∞ | 1 | 0.6965 | 1.0000 |
| | > 25 / < 20 | ≥ 90 | > 36 | 2.0869 | 1.0695 | ∞ | 1 | 1.4355 | 0.4534 |
| | > 25 / [20. 25] | ≥ 90 | > 36 | 1.4299 | 0.7617 | ∞ | 1 | 0.6713 | 1.0000 |
| Subgroup C (n = 105) | [20. 25] / < 20 | < 90 | ≤ 36 | 2.7489 | 1.9658 | ∞ | 1 | 1.4140 | 0.4721 |
| | > 25 / < 20 | < 90 | ≤ 36 | 5.8917 | 3.9488 | ∞ | 1 | 2.6461 | **0.0244** |
| | > 25 / [20. 25] | < 90 | ≤ 36 | 2.1433 | 1.2237 | ∞ | 1 | 1.3352 | 0.5454 |
| Subgroup D (n = 369) | [20. 25] / < 20 | ≥ 90 | ≤ 36 | 1.6207 | 0.6629 | ∞ | 1 | 1.1805 | 0.7134 |
| | > 25 / < 20 | ≥ 90 | ≤ 36 | 4.6641 | 2.1563 | ∞ | 1 | 3.3307 | **0.0026** |
| | > 25 / [20. 25] | ≥ 90 | ≤ 36 | 2.8778 | 1.3829 | ∞ | 1 | 2.1996 | **0.0835** |

Abbreviations: C72mean, mean vancomycin serum concentration during the first three days of continuous infusion of vancomycin [mg/L]; df, degree of freedom; $eGFR_{sCIV}$, estimated glomerular filtration rate at the start of continuous infusion of vancomycin; HR, hazard ratio; $SAPS_{sCIV}$, Simplified Acute Physiology Score II at the start of continuous infusion of vancomycin; SE, standard error.

[1] p-value <0.05 was considered significant.

#### *1.10.7 Clinical failure with survival*

**Table S 28** Post-hoc comparison of the classes of mean vancomycin serum concentration during the first three days of continuous infusion of vancomycin (C72mean) within the subgroups for estimated glomerular filtration rate at the start of continuous infusion of vancomycin ($eGFR_{sCIV}$) and Simplified Acute Physiology Score II at the start of continuous infusion of vancomycin ($SAPS_{sCIV}$) for clinical failure with survival with Bonferroni correction based on multivariate logistic regression.

| | C72mean contrast | $eGFR_{sCIV}$ | $SAPS_{sCIV}$ | OR | SE | df | null | z ratio | p-value[1] |
|---|---|---|---|---|---|---|---|---|---|
| Subgroup A (n = 262) | [20. 25] / < 20 | < 90 | > 36 | 1.1684 | 0.6198 | ∞ | 1 | 0.2934 | 1.0000 |
| | > 25 / < 20 | < 90 | > 36 | 1.5432 | 0.7317 | ∞ | 1 | 0.9151 | 1.0000 |
| | > 25 / [20. 25] | < 90 | > 36 | 1.3208 | 0.5678 | ∞ | 1 | 0.6472 | 1.0000 |
| Subgroup B (n = 186) | [20. 25] / < 20 | ≥ 90 | > 36 | 1.4654 | 0.7266 | ∞ | 1 | 0.7706 | 1.0000 |
| | > 25 / < 20 | ≥ 90 | > 36 | 3.2042 | 1.4927 | ∞ | 1 | 2.4997 | **0.0373** |
| | > 25 / [20. 25] | ≥ 90 | > 36 | 2.1866 | 1.1029 | ∞ | 1 | 1.5511 | 0.3627 |
| Subgroup C (n = 105) | [20. 25] / < 20 | < 90 | ≤ 36 | 0.9522 | 0.4899 | ∞ | 1 | -0.0951 | 1.0000 |
| | > 25 / < 20 | < 90 | ≤ 36 | 0.6356 | 0.3023 | ∞ | 1 | -0.9527 | 1.0000 |
| | > 25 / [20. 25] | < 90 | ≤ 36 | 0.6675 | 0.3345 | ∞ | 1 | -0.8068 | 1.0000 |
| Subgroup D (n = 369) | [20. 25] / < 20 | ≥ 90 | ≤ 36 | 1.1943 | 0.3454 | ∞ | 1 | 0.6138 | 1.0000 |
| | > 25 / < 20 | ≥ 90 | ≤ 36 | 1.3197 | 0.5289 | ∞ | 1 | 0.6923 | 1.0000 |
| | > 25 / [20. 25] | ≥ 90 | ≤ 36 | 1.1051 | 0.4744 | ∞ | 1 | 0.2327 | 1.0000 |

Abbreviations: C72mean, mean vancomycin serum concentration during the first three days of continuous infusion of vancomycin [mg/L]; df, degree of freedom; $eGFR_{sCIV}$, estimated glomerular filtration rate at the start of continuous infusion of vancomycin [mL/min/1.73 $m^2$]; OR, odds ratio; $SAPS_{sCIV}$, Simplified Acute Physiology Score II at the start of continuous infusion of vancomycin; SE, standard error.

[1] p-value <0.05 was considered significant.

##### *1.10.8 Microbiological failure*

**Table S 29** Post-hoc comparison of the classes of mean vancomycin serum concentration during the first three days of continuous infusion of vancomycin (C72mean) within the subgroups for estimated glomerular filtration rate at the start of continuous infusion of vancomycin ($eGFR_{sCIV}$) and Simplified Acute Physiology Score II at the start of continuous infusion of vancomycin ($SAPS_{sCIV}$) for microbiological failure with Bonferroni correction based on multivariate logistic regression.

| | C72mean contrast | $eGFR_{sCIV}$ | $SAPS_{sCIV}$ | OR | SE | df | null | z ratio | p-value[1] |
|---|---|---|---|---|---|---|---|---|---|
| Subgroup A (n = 262) | [20, 25] / < 20 | < 90 | > 36 | 1.6254 | 0.9129 | ∞ | 1 | 0.8649 | 1.0000 |
| | > 25 / < 20 | < 90 | > 36 | 1.8078 | 0.8852 | ∞ | 1 | 1.2093 | 0.6796 |
| | > 25 / [20, 25] | < 90 | > 36 | 1.1122 | 0.5055 | ∞ | 1 | 0.2340 | 1.0000 |
| Subgroup B (n = 186) | [20, 25] / < 20 | ≥ 90 | > 36 | 0.4986 | 0.2360 | ∞ | 1 | -1.4702 | 0.4245 |
| | > 25 / < 20 | ≥ 90 | > 36 | 0.5000 | 0.2428 | ∞ | 1 | -1.4272 | 0.4605 |
| | > 25 / [20, 25] | ≥ 90 | > 36 | 1.0028 | 0.5439 | ∞ | 1 | 0.0052 | 1.0000 |
| Subgroup C (n = 105) | [20, 25] / < 20 | < 90 | ≤ 36 | 1.5205 | 0.9580 | ∞ | 1 | 0.6650 | 1.0000 |
| | > 25 / < 20 | < 90 | ≤ 36 | 2.5839 | 1.4834 | ∞ | 1 | 1.6535 | 0.2947 |
| | > 25 / [20, 25] | < 90 | ≤ 36 | 1.6994 | 1.0277 | ∞ | 1 | 0.8768 | 1.0000 |
| Subgroup D (n = 369) | [20, 25] / < 20 | ≥ 90 | ≤ 36 | 0.4664 | 0.1702 | ∞ | 1 | -2.0906 | 0.1097 |
| | > 25 / < 20 | ≥ 90 | ≤ 36 | 0.7146 | 0.3212 | ∞ | 1 | -0.7477 | 1.0000 |
| | > 25 / [20, 25] | ≥ 90 | ≤ 36 | 1.5322 | 0.7766 | ∞ | 1 | 0.8419 | 1.0000 |

Abbreviations: C72mean, mean vancomycin serum concentration during the first three days of continuous infusion of vancomycin [mg/L]; df, degree of freedom; $eGFR_{sCIV}$, estimated glomerular filtration rate at the start of continuous infusion of vancomycin [mL/min/1.73 m²]; OR, odds ratio; $SAPS_{sCIV}$, Simplified Acute Physiology Score II at the start of continuous infusion of vancomycin; SE, standard error.

[1] p-value <0.05 was considered significant.

### 1.11 ROC Analysis

**Table S 30** Cut-off values of mean vancomycin serum concentration during the first three days of continuous infusion of vancomycin (C72mean) during continuous infusion of vancomycin for therapeutic efficacy and safety determined by receiver-operating characteristic (ROC) and associated performance measures.

| Group | Outcome parameter | C72mean cut-off [mg/L] | Youden Index | Error rate | Sensitivity | Specificity | PPV | NPV | AUC |
|---|---|---|---|---|---|---|---|---|---|
| Overall (n= 922) | ICU mortality | **23.532** | 0.260 | 0.333 | 0.573 | 0.686 | 0.273 | 0.887 | 0.655 |
| | In-hospital mortality | **22.859** | 0.255 | 0.347 | 0.578 | 0.677 | 0.366 | 0.833 | 0.662 |
| | 30-day mortality | **26.394** | 0.206 | 0.289 | 0.434 | 0.772 | 0.295 | 0.861 | 0.626 |
| | AKI (total) | **24.132** | 0.309 | 0.305 | 0.590 | 0.720 | 0.327 | 0.884 | 0.687 |
| | AKI (early) | **24.333** | 0.390 | 0.297 | 0.686 | 0.704 | 0.160 | 0.965 | 0.721 |
| | AKI (late) | **23.035** | 0.227 | 0.362 | 0.583 | 0.645 | 0.171 | 0.925 | 0.631 |
| | Clinical failure (survival) | **21.872** | 0.069 | 0.525 | 0.626 | 0.443 | 0.194 | 0.846 | 0.529 |
| | Microbiological failure | **19.433** | 0.092 | 0.431 | 0.486 | 0.606 | 0.352 | 0.728 | 0.541 |
| Subgroup A (n= 262) | ICU mortality | **28.999** | 0.168 | 0.386 | 0.506 | 0.661 | 0.392 | 0.756 | 0.563 |
| | In-hospital mortality | **28.999** | 0.159 | 0.405 | 0.482 | 0.678 | 0.520 | 0.644 | 0.570 |
| | 30-day mortality | **28.999** | 0.206 | 0.370 | 0.530 | 0.676 | 0.431 | 0.756 | 0.575 |
| | AKI (total) | **27.573** | 0.163 | 0.420 | 0.584 | 0.578 | 0.366 | 0.770 | 0.551 |
| | AKI (early) | **26.455** | 0.238 | 0.485 | 0.758 | 0.480 | 0.174 | 0.932 | 0.597 |
| | AKI (late) | **27.407** | 0.073 | 0.469 | 0.545 | 0.528 | 0.189 | 0.852 | 0.500 |
| | Clinical failure (survival) | **32.291** | 0.087 | 0.683 | 0.844 | 0.243 | 0.134 | 0.918 | 0.512 |
| | Microbiological failure | **27.784** | 0.092 | 0.481 | 0.608 | 0.484 | 0.317 | 0.758 | 0.522 |
| Subgroup B (n= 186) | ICU mortality | **22.135** | 0.187 | 0.333 | 0.484 | 0.703 | 0.246 | 0.872 | 0.581 |
| | In-hospital mortality | **17.344** | 0.210 | 0.532 | 0.860 | 0.350 | 0.285 | 0.893 | 0.606 |
| | 30-day mortality | **25.312** | 0.165 | 0.661 | 0.926 | 0.239 | 0.171 | 0.950 | 0.529 |
| | AKI (total) | **24.047** | 0.332 | 0.247 | 0.529 | 0.803 | 0.375 | 0.884 | 0.681 |
| | AKI (early) | **29.991** | 0.293 | 0.129 | 0.375 | 0.918 | 0.300 | 0.940 | 0.671 |
| | AKI (late) | **24.047** | 0.329 | 0.247 | 0.556 | 0.774 | 0.208 | 0.942 | 0.655 |
| | Clinical failure (survival) | **24.047** | 0.301 | 0.253 | 0.522 | 0.079 | 0.250 | 0.920 | 0.647 |
| | Microbiological failure | **19.285**[1] | 0.067 | 0.484 | 0.583 | 0.484 | 0.350 | 0.709 | 0.496[1] |
| Subgroup C (n= 105) | ICU mortality | **24.301** | 0.334 | 0.410 | 0.769 | 0.565 | 0.200 | 0.945 | 0.654 |
| | In-hospital mortality | **24.301** | 0.298 | 0.390 | 0.714 | 0.583 | 0.300 | 0.891 | 0.645 |
| | 30-day mortality | **25.885** | 0.042 | 0.333 | 0.750 | 0.652 | 0.279 | 0.935 | 0.683 |
| | AKI (total) | **25.885** | 0.549 | 0.295 | 0.875 | 0.674 | 0.326 | 0.968 | 0.759 |
| | AKI (early) | **26.195** | 0.640 | 0.343 | 1.000 | 0.640 | 0.122 | 1.000 | 0.780 |
| | AKI (late) | **24.301** | 0.484 | 0.390 | 0.909 | 0.574 | 0.200 | 0.982 | 0.721 |
| | Clinical failure (survival) | **21.807** | 0.184 | 0.381 | 0.542 | 0.642 | 0.310 | 0.825 | 0.573 |
| | Microbiological failure | **31.592** | 0.150 | 0.533 | 0.839 | 0.311 | 0.338 | 0.821 | 0.561 |
| Subgroup D (n= 369) | ICU mortality | **16.257** | 0.200 | 0.558 | 0.794 | 0.406 | 0.119 | 0.951 | 0.593 |
| | In-hospital mortality | **16.554** | 0.228 | 0.509 | 0.784 | 0.443 | 0.184 | 0.928 | 0.592 |
| | 30-day mortality | **20.423**[1] | 0.066 | 0.637 | 0.750 | 0.316 | 0.118 | 0.912 | 0.460[1] |
| | AKI (total) | **19.158** | 0.346 | 0.344 | 0.696 | 0.650 | 0.221 | 0.938 | 0.699 |
| | AKI (early) | **19.158** | 0.569 | 0.355 | 0.938 | 0.632 | 0.103 | 0.996 | 0.828 |
| | AKI (late) | **19.712** | 0.219 | 0.355 | 0.567 | 0.652 | 0.126 | 0.944 | 0.608 |
| | Clinical failure (survival) | **19.786**[1] | 0.045 | 0.407 | 0.393 | 0.653 | 0.250 | 0.785 | 0.492[1] |
| | Microbiological failure | **19.429** | 0.155 | 0.477 | 0.727 | 0.429 | 0.371 | 0.771 | 0.557 |

Subgroup A: eGFR at sCIV <90 mL/min/1.73 m² and SAPS II at sCIV >36; subgroup B: eGFR at sCIV ≥90 mL/min/1.73 m² and SAPS II at sCIV >36; subgroup C: eGFR at sCIV <90 mL/min/1.73 m² and SAPS II at sCIV ≤36; subgroup D: eGFR at sCIV ≥90 mL/min/1.73 m² and SAPS II at sCIV ≤36.

Abbreviations: AKI, acute kidney injury; AUC, area under the curve; C72mean, mean vancomycin serum concentration during the first three days of continuous infusion of vancomycin; eGFR, estimated glomerular filtration rate at the start of continuous infusion of vancomycin; ICU, intensive care unit; NPV, negative predicted value; PPV, positive predicted value; SAPS, Simplified Acute Physiology Score II at the start of continuous infusion of vancomycin; sCIV, start of continuous infusion of vancomycin.

[1] ≥ cut-off result was worse classifier than guessing, i.e. area under the curve (AUC) <0.5

### 1.12 CART Analysis

**Table S 31** Cut-off values of mean vancomycin serum concentration during the first three days of continuous infusion of vancomycin (C72mean) during continuous infusion of vancomycin for therapeutic efficacy and safety determined by classification and regression tree (CART) and associated performance measures

| | Outcome parameter | **C72mean cut-off** [mg/L] | Youden Index | Error rate | Sensitivity | Specificity | PPV | NPV | AUC |
|---|---|---|---|---|---|---|---|---|---|
| **Overall (n= 922)** | ICU mortality | **27.812** | 0.231 | 0.247 | 0.408 | 0.824 | 0.322 | 0.871 | 0.655 |
| | In-hospital mortality | **26.195** | 0.238 | 0.294 | 0.449 | 0.789 | 0.407 | 0.816 | 0.662 |
| | 30-day mortality | **29.039** | 0.197 | 0.242 | 0.349 | 0.848 | 0.335 | 0.856 | 0.626 |
| | AKI (total) | **24.132** | 0.309 | 0.305 | 0.590 | 0.720 | 0.327 | 0.884 | 0.687 |
| | AKI (early) | **27.673** | 0.349 | 0.214 | 0.543 | 0.806 | 0.187 | 0.955 | 0.721 |
| | AKI (late) | **23.035** | 0.227 | 0.362 | 0.583 | 0.645 | 0.171 | 0.925 | 0.631 |
| | Clinical failure (survival) | **13.352** | 0.064 | 0.223 | 0.153 | 0.910 | 0.269 | 0.834 | 0.529 |
| | Microbiological failure | **19.433** | 0.092 | 0.431 | 0.486 | 0.606 | 0.352 | 0.730 | 0.541 |
| **Subgroup A (n= 262)** | ICU mortality | **20.467** | 0.141 | 0.565 | 0.911 | 0.230 | 0.338 | 0.857 | 0.563 |
| | In-hospital mortality | **20.856** | 0.154 | 0.473 | 0.891 | 0.263 | 0.467 | 0.769 | 0.570 |
| | 30-day mortality | **28.999** | 0.206 | 0.370 | 0.530 | 0.676 | 0.431 | 0.756 | 0.575 |
| | AKI (total) | **27.573** | 0.163 | 0.420 | 0.584 | 0.578 | 0.366 | 0.770 | 0.551 |
| | AKI (early) | **26.455** | 0.238 | 0.485 | 0.758 | 0.480 | 0.174 | 0.932 | 0.597 |
| | AKI (late) | **33.325** | -0.102 | 0.328 | 0.114 | 0.784 | 0.096 | 0.614 | 0.500 |
| | Clinical failure (survival) | **15.875** | 0.055 | 0.145 | 0.094 | 0.961 | 0.250 | 0.884 | 0.512 |
| | Microbiological failure | **46.105** | 0.037 | 0.691 | 1.000 | 0.037 | 0.290 | 1.000 | 0.522 |
| **Subgroup B (n= 186)** | ICU mortality | **22.135** | 0.187 | 0.333 | 0.484 | 0.703 | 0.246 | 0.872 | 0.581 |
| | In-hospital mortality | **17.344** | 0.210 | 0.532 | 0.860 | 0.350 | 0.285 | 0.893 | 0.606 |
| | 30-day mortality | **25.312** | 0.165 | 0.661 | 0.926 | 0.239 | 0.071 | 0.950 | 0.529 |
| | AKI (total) | **24.047** | 0.332 | 0.247 | 0.529 | 0.803 | 0.375 | 0.884 | 0.681 |
| | AKI (early) | **29.991** | 0.293 | 0.129 | 0.375 | 0.918 | 0.300 | 0.940 | 0.671 |
| | AKI (late) | **24.047** | 0.329 | 0.247 | 0.556 | 0.774 | 0.208 | 0.942 | 0.655 |
| | Clinical failure (survival) | **24.047** | 0.301 | 0.253 | 0.522 | 0.779 | 0.250 | 0.920 | 0.647 |
| | Microbiological failure | **23.588**[1] | -0.085 | 0.457 | 0.217 | 0.698 | 0.255 | 0.652 | 0.496[1] |
| **Subgroup C (n= 105)** | ICU mortality | **27.509** | 0.322 | 0.305 | 0.615 | 0.707 | 0.229 | 0.929 | 0.654 |
| | In-hospital mortality | **24.301** | 0.298 | 0.390 | 0.714 | 0.583 | 0.300 | 0.891 | 0.645 |
| | 30-day mortality | **26.959** | 0.395 | 0.295 | 0.688 | 0.708 | 0.297 | 0.926 | 0.683 |
| | AKI (total) | **25.885** | 0.549 | 0.295 | 0.875 | 0.674 | 0.326 | 0.968 | 0.759 |
| | AKI (early) | **26.195** | 0.640 | 0.343 | 1.000 | 0.640 | 0.122 | 1.000 | 0.780 |
| | AKI (late) | **24.301** | 0.484 | 0.390 | 0.909 | 0.574 | 0.200 | 0.982 | 0.721 |
| | Clinical failure (survival) | **36.350** | 0.136 | 0.667 | 1.000 | 0.136 | 0.255 | 1.000 | 0.573 |
| | Microbiological failure | **31.592** | 0.150 | 0.533 | 0.839 | 0.311 | 0.338 | 0.821 | 0.561 |
| **Subgroup D (n= 369)** | ICU mortality | **27.555** | 0.144 | 0.106 | 0.176 | 0.967 | 0.353 | 0.920 | 0.593 |
| | In-hospital mortality | **27.555** | 0.106 | 0.146 | 0.137 | 0.969 | 0.412 | 0.875 | 0.592 |
| | 30-day mortality | **14.025**[1] | -0.149 | 0.320 | 0.100 | 0.751 | 0.047 | 0.873 | 0.460[1] |
| | AKI (total) | **30.154** | 0.177 | 0.117 | 0.196 | 0.981 | 0.600 | 0.895 | 0.699 |
| | AKI (early) | **33.856** | 0.242 | 0.041 | 0.250 | 0.992 | 0.571 | 0.967 | 0.828 |
| | AKI (late) | **19.712** | 0.219 | 0.355 | 0.567 | 0.652 | 0.126 | 0.944 | 0.608 |
| | Clinical failure (survival) | **13.379**[1] | -0.093 | 0.702 | 0.738 | 0.168 | 0.207 | 0.686 | 0.492[1] |
| | Microbiological failure | **19.429** | 0.155 | 0.477 | 0.727 | 0.429 | 0.371 | 0.771 | 0.557 |

Subgroup A: eGFR at sCIV <90 mL/min/1.73 m² and SAPS II at sCIV >36; subgroup B: eGFR at sCIV ≥90 mL/min/1.73 m² and SAPS II at sCIV >36; subgroup C: eGFR at sCIV <90 mL/min/1.73 m² and SAPS II at sCIV ≤36; subgroup D: eGFR at sCIV ≥90 mL/min/1.73 m² and SAPS II at sCIV ≤36.

Abbreviations: AKI, acute kidney injury; AUC, area under the curve; C72mean, mean vancomycin serum concentration during the first three days of continuous infusion of vancomycin; eGFR, estimated glomerular filtration rate at the start of continuous infusion of vancomycin; ICU, intensive care unit; NPV, negative predicted value; PPV, positive predicted value; SAPS, Simplified Acute Physiology Score II at the start of continuous infusion of vancomycin; sCIV, start of continuous infusion of vancomycin.

[1] ≥ cut-off result was worse classifier than guessing, i.e. area under the curve (AUC) <0.5

### 1.13 Concentration groups

**Table S 32** Characteristics of the study population, with regard to concentration groups of mean vancomycin serum concentration during the first three days of continuous infusion of vancomycin (C72mean [mg/L]).

| | **Characteristics** | **N** | **Overall** (n=922[1]) | **<15** (n=161[1]) | **15-20** (n=258[1]) | **20-25** (n=221[1]) | **25-30** (n=126[1]) | **>30** (n=156[1]) | **p-value**[2] |
|---|---|---|---|---|---|---|---|---|---|
| **Overall** (n = 922) | **eGFR** | 922 | 97 (72, 113) | 113 (102, 127) | 106 (93, 118) | 97 (77, 109) | 72 (46, 92) | 66 (47, 87) | **<0.001** |
| | **SAPS** | 922 | 36 (28, 45) | 30 (25, 37) | 33 (26, 40) | 36 (30, 43) | 43 (36, 49) | 42 (35, 49) | **<0.001** |
| | **CL$_{vancomycin}$** day 1-3 | 922 | 4.9 (3.5, 6.6) | 8.6 (7.1, 10.5) | 6.1 (5.3, 6.9) | 4.6 (4.0, 5.3) | 3.5 (3.2, 3.8) | 2.7 (2.2, 3.0) | **<0.001** |
| | **Dose day 1-3** [mg/kg TBW/day] | 922 | 31 (26, 37) | 30 (25, 36) | 33 (28, 39) | 32 (27, 38) | 29 (24, 35) | 29 (24, 36) | **<0.001** |
| | **ICU mortality** | 922 | 157 (17%) | 10 (6.2%) | 34 (13%) | 35 (16%) | 31 (25%) | 47 (30%) | **<0.001** |
| | **In-hospital mortality** | 922 | 225 (24%) | 14 (8.7%) | 50 (19%) | 52 (24%) | 43 (34%) | 66 (42%) | **<0.001** |
| | **30-day mortality** | 922 | 166 (18%) | 13 (8.1%) | 41 (16%) | 35 (16%) | 29 (23%) | 48 (31%) | **<0.001** |
| | **Clinical failure with survival** | 922 | 163 (18%) | 34 (21%) | 41 (16%) | 42 (19%) | 21 (17%) | 25 (16%) | 0.6 |

| | **Characteristics** | **N** | **Overall** (n=262[1]) | **<15** (n=10[1]) | **15-20** (n=34[1]) | **20-25** (n=56[1]) | **25-30** (n=73[1]) | **>30** (n=89[1]) | **p-value**[2] |
|---|---|---|---|---|---|---|---|---|---|
| **Subgroup A** (n = 262) | **eGFR** | 262 | 61 (43, 76) | 68 (56, 86) | 64 (54, 80) | 66 (44, 77) | 57 (38, 73) | 59 (42, 70) | 0.11 |
| | **SAPS** | 262 | 46 (41, 53) | 41 (38, 48) | 46 (40, 52) | 47 (43, 54) | 47 (42, 53) | 46 (42, 57) | 0.3 |
| | **CL$_{vancomycin}$** day 1-3 | 262 | 3.4 (2.6, 4.4) | 6.9 (6.5, 7.8) | 5.5 (4.7, 6.5) | 4.1 (3.5, 4.7) | 3.4 (3.0, 3.7) | 2.6 (2.2, 3.0) | **<0.001** |
| | **Dose day 1-3** [mg/kg TBW/day] | 262 | 28 (23, 32) | 26 (17, 29) | 30 (22, 33) | 27 (23, 30) | 27 (23, 31) | 29 (23, 35) | 0.2 |
| | **ICU mortality** | 262 | 79 (30%) | 2 (20%) | 5 (15%) | 17 (30%) | 25 (34%) | 30 (34%) | 0.2 |
| | **In-hospital mortality** | 262 | 110 (42%) | 3 (30%) | 8 (24%) | 24 (43%) | 32 (44%) | 43 (48%) | 0.13 |
| | **30-day mortality** | 262 | 83 (32%) | 3 (30%) | 5 (15%) | 17 (30%) | 24 (33%) | 34 (38%) | 0.2 |
| | **Clinical failure with survival** | 262 | 32 (12%) | 2 (20%) | 3 (8.8%) | 8 (14%) | 9 (12%) | 10 (11%) | 0.8 |

| | **Characteristics** | **N** | **Overall** (n=186[1]) | **<15** (n=33[1]) | **15-20** (n=64[1]) | **20-25** (n=48[1]) | **25-30** (n=21[1]) | **>30** (n=20[1]) | **p-value**[2] |
|---|---|---|---|---|---|---|---|---|---|
| **Subgroup B** (n = 186) | **eGFR** | 186 | 102 (96, 111) | 105 (101, 115) | 103 (97, 114) | 105 (96, 112) | 100 (92, 103) | 97 (95, 101) | **0.003** |
| | **SAPS** | 186 | 42 (39, 47) | 40 (39, 43) | 43 (38, 47) | 42 (40, 45) | 42 (39, 45) | 48 (42, 52) | 0.058 |
| | **CL$_{vancomycin}$** day 1-3 | 186 | 5.4 (4.2, 6.5) | 8.3 (6.5, 10.5) | 6.0 (5.4, 6.8) | 4.8 (4.4, 5.5) | 3.7 (3.3, 3.8) | 2.8 (2.4, 3.0) | **<0.001** |
| | **Dose day 1-3** [mg/kg TBW/day] | 186 | 33 (28, 40) | 30 (23, 35) | 33 (29, 40) | 34 (30, 40) | 37 (29, 46) | 32 (28, 36) | **0.033** |
| | **ICU mortality** | 186 | 31 (17%) | 3 (9.1%) | 10 (16%) | 10 (21%) | 2 (9.5%) | 6 (30%) | 0.3 |
| | **In-hospital mortality** | 186 | 43 (23%) | 4 (12%) | 14 (22%) | 12 (25%) | 5 (24%) | 8 (40%) | 0.2 |
| | **30-day mortality** | 186 | 27 (15%) | 3 (9.1%) | 13 (20%) | 8 (17%) | 1 (4.8%) | 2 (10%) | 0.4 |
| | **Clinical failure with survival** | 186 | 23 (12%) | 3 (9.1%) | 5 (7.8%) | 5 (10%) | 5 (24%) | 5 (25%) | 0.13 |

| | **Characteristics** | **N** | **Overall** (n=105[1]) | **<15** (n=10[1]) | **15-20** (n=23[1]) | **20-25** (n=25[1]) | **25-30** (n=16[1]) | **>30** (n=31[1]) | **p-value**[2] |
|---|---|---|---|---|---|---|---|---|---|
| **Subgroup C** (n = 105) | **eGFR** | 105 | 75 (53, 84) | 82 (76, 87) | 84 (76, 88) | 76 (64, 86) | 67 (44, 80) | 56 (38, 73) | **<0.001** |
| | **SAPS** | 105 | 32 (28, 34) | 28 (26, 31) | 32 (28, 34) | 33 (29, 34) | 31 (30, 34) | 32 (29, 35) | 0.13 |
| | **CL$_{vancomycin}$** day 1-3 | 105 | 4.3 (3.1, 5.8) | 9.1 (7.5, 9.5) | 5.9 (5.2, 6.5) | 4.5 (4.0, 4.7) | 3.6 (3.3, 3.8) | 2.7 (2.1, 3.1) | **<0.001** |
| | **Dose day 1-3** [mg/kg TBW/day] | 105 | 28 (24, 34) | 26 (23, 29) | 26 (24, 30) | 30 (25, 37) | 27 (24, 33) | 30 (23, 34) | 0.4 |
| | **ICU mortality** | 105 | 13 (12%) | 0 (0%) | 3 (13%) | 1 (4.0%) | 3 (19%) | 6 (19%) | 0.3 |
| | **In-hospital mortality** | 105 | 21 (20%) | 0 (0%) | 4 (17%) | 3 (12%) | 5 (31%) | 9 (29%) | 0.2 |
| | **30-day mortality** | 105 | 16 (15%) | 0 (0%) | 3 (13%) | 1 (4.0%) | 4 (25%) | 8 (26%) | 0.081 |
| | **Clinical failure with survival** | 105 | 24 (23%) | 3 (30%) | 5 (22%) | 6 (24%) | 3 (19%) | 7 (23%) | >0.9 |

| | **Characteristics** | **N** | **Overall** (n=369[1]) | **<15** (n=108[1]) | **15-20** (n=137[1]) | **20-25** (n=92[1]) | **25-30** (n=16[1]) | **>30** (n=16[1]) | **p-value**[2] |
|---|---|---|---|---|---|---|---|---|---|
| **Subgroup D** (n = 369) | **eGFR** | 369 | 113 (104, 124) | 119 (109, 130) | 114 (106, 123) | 107 (100, 117) | 105 (98, 113) | 113 (105, 116) | **<0.001** |
| | **SAPS** | 369 | 28 (23, 32) | 27 (23, 30) | 27 (23, 32) | 29 (23, 33) | 30 (28, 33) | 26 (22, 31) | 0.2 |
| | **CL$_{vancomycin}$** day 1-3 | 369 | 6.2 (4.7, 7.9) | 8.9 (7.8, 10.5) | 6.4 (5.5, 7.4) | 4.7 (4.1, 5.4) | 3.8 (3.7, 4.2) | 2.9 (2.6, 3.2) | **<0.001** |
| | **Dose day 1-3** [mg/kg TBW/day] | 369 | 34 (28, 40) | 32 (27, 37) | 35 (29, 40) | 34 (29, 40) | 35 (31, 40) | 34 (28, 40) | 0.064 |
| | **ICU mortality** | 369 | 34 (9.2%) | 5 (4.6%) | 16 (12%) | 7 (7.6%) | 1 (6.3%) | 5 (31%) | **0.018** |
| | **In-hospital mortality** | 369 | 51 (14%) | 7 (6.5%) | 24 (18%) | 13 (14%) | 1 (6.3%) | 6 (38%) | **0.006** |
| | **30-day mortality** | 369 | 40 (11%) | 7 (6.5%) | 20 (15%) | 9 (9.8%) | 0 (0%) | 4 (25%) | 0.052 |
| | **Clinical failure with survival** | 369 | 84 (23%) | 26 (24%) | 28 (20%) | 23 (25%) | 4 (25%) | 3 (19%) | >0.9 |

Subgroup A: eGFR at sCIV <90 mL/min/1.73 m² and SAPS II at sCIV >36; subgroup B: eGFR at sCIV ≥90 mL/min/1.73 m² and SAPS II at sCIV >36; subgroup C: eGFR at sCIV <90 mL/min/1.73 m² and SAPS II at sCIV ≤36; subgroup D: eGFR at sCIV ≥90 mL/min/1.73 m² and SAPS II at sCIV ≤36.

Abbreviations: CL, clearance [L/h]; C72mean, mean vancomycin serum concentration during the first three days of continuous infusion of vancomycin [mg/L]; eGFR, estimated glomerular filtration rate; SAPS, Simplified Acute Physiology Score II; sCIV, start of continuous infusion of vancomycin; TBW, total body weight.

[1] n (%); Median (IQR); [2] Kruskal-Wallis rank sum test; Pearson's Chi-squared test; Fisher's exact test; p-value <0.05 was considered significant.
